# Comparative safety and sedation-related outcomes of fospropofol disodium versus propofol for adult sedation outside general anaesthesia induction or maintenance (including ICU and procedural sedation): a systematic review, meta-analysis and trial sequential analysis

**DOI:** 10.64898/2026.09.22.26363627

**Authors:** Guiping Xu, Yutong Shi, Yang Wang, Alimujiang Simayi, Li Qu

## Abstract

**Background:** Fospropofol disodium is a water-soluble prodrug of propofol that avoids a lipid emulsion carrier. It may therefore reduce injection pain, cardiorespiratory depression and lipid exposure. In May 2021 it was approved in China (H20210017) for induction of general anaesthesia in adults. Randomised trials have since tested it for sedation outside the operating room, but this setting has not been systematically reviewed.

**Methods:** We followed PRISMA 2020 and PRISMA-S and searched Europe PMC, PubMed, Embase, Cochrane CENTRAL, Web of Science and the Chinese databases CNKI, Wanfang and VIP (the last accessed through the XJMU Library database proxy) from inception to 2026. Eligible patients were adults receiving sedation for purposes other than general anaesthesia induction or maintenance, including continuous sedation during ICU mechanical ventilation and procedural sedation. These uses are referred to here as sedation outside the operating room. We included randomised controlled trials with propofol as the comparator and excluded trials of general anaesthesia induction or maintenance to avoid overlap with the published induction meta-analysis. Binary outcomes were pooled with a DerSimonian-Laird random-effects model, and REML plus modified Hartung-Knapp estimates were reported alongside to improve interval coverage when few studies were available. Outcomes with a high proportion of zero cells were also analysed with the Peto method. We stratified by setting and blinding and performed leave-one-out analysis, trial sequential analysis (TSA) and GRADE assessment. Hypotension reached the prespecified threshold of at least 10 studies, so publication bias was assessed with the Harbord test, with the originally specified Egger’s test reported alongside. The protocol was prospectively registered and took effect on 16 September 2026. All searches were completed on 21 September 2026, and screening, extraction and analysis were performed after registration.

**Results:** Twelve randomised controlled trials were included. Event counts for all 12 studies were checked against the full-text original publications. For Liu 2026, a preprint, counts were taken directly from its Tables 2 and 3. Compared with propofol, fospropofol disodium reduced injection pain under the DerSimonian-Laird model (8 studies, OR 0.09, 0.03–0.21) and increased pruritus/paraesthesia (4 studies, OR 32.56, 12.89–82.26; RD +28.5%). During procedural sedation, it was associated with fewer respiratory adverse events (5 studies, OR 0.19, 0.11–0.32). This composite includes respiratory depression, hypoxaemia and related events reported by individual trials and should not be extrapolated to all adult sedation settings. For hypertriglyceridaemia, the DerSimonian-Laird estimate was OR 0.29 (0.13–0.65), but all four studies were conducted in ICU settings and the REML plus modified Hartung-Knapp estimate was 0.29 (0.08–1.08), with a confidence interval crossing 1. Certainty was very low, so this finding remains hypothesis-generating. Hypotension differed by setting: OR 0.24 (0.13–0.42) for procedural sedation and 0.90 (0.53–1.55) for continuous ICU sedation, with a significant interaction (P < 0.001). The overall estimate (OR 0.51, 0.28–0.93) is reported for summary only. Under REML plus modified Hartung-Knapp it was 0.51 (0.25–1.03), crossing 1, so it does not support a claim that the drug lowers blood pressure and should not be extrapolated to the ICU. Bradycardia did not differ significantly (9 studies, OR 0.45, 0.17–1.18), and the direction reversed in the only double-blind study. Two studies reported slower onset or loss of consciousness with fospropofol disodium (MD +2.22 and +0.92 min), but these were not pooled because heterogeneity was extreme (*I*^2^ = 99.6%). Recovery, discharge and successful extubation showed no clear differences, although heterogeneity was high (*I*^2^ = 85%–92%) and the evidence cannot establish equivalence. Sedation efficacy was not pooled because definitions differed. Patient-important outcomes (death, delirium, ICU length of stay and duration of mechanical ventilation) were each reported by only one or two small studies with inconsistent definitions and time points, so no reliable conclusion can be drawn. Setting-specific results should take priority over the overall mean. Certainty ranged from very low to moderate.

**Conclusions:** During procedural sedation, fospropofol disodium may reduce injection pain and respiratory adverse events and may lower the risk of hypotension compared with propofol. This hypotension benefit should not be extrapolated to the ICU. During continuous ICU sedation no data were available for injection pain or respiratory adverse events, so the safety evidence in that setting remains limited. Hypertriglyceridaemia was lower only under the DerSimonian-Laird model. Because the result depends on the method and certainty is very low, it cannot be treated as definitive. The main cost is a clear increase in pruritus and abnormal sensations. Onset may be slower, but the evidence comes from two bolus-dosing studies in procedural sedation, and no such disadvantage was seen during continuous ICU infusion. Evidence on sedation efficacy, recovery time, discharge time and successful extubation is limited, and equivalence between the two drugs cannot be asserted. Because most studies were small and open-label and certainty ranged from very low to moderate, these findings support investigational use only and do not justify a routine clinical recommendation.

**Research in context:** *Evidence before this study:* We searched Europe PMC, PubMed, Embase, Cochrane CENTRAL, Web of Science Core Collection and Chinese databases (CNKI, Wanfang, VIP) by crossing drug terms (fospropofol, fospropofol disodium, Lusedra, Aquavan) with sedation setting terms (ICU, mechanical ventilation, endoscopy, gastroscopy, hysteroscopy, bronchoscopy, procedural sedation). Within the retrieved literature, the only relevant meta-analysis addressed general anaesthesia induction; no synthesis of the sedation setting was identified.

*Added value of this study:* The present synthesis focuses on sedation outside the operating room, includes 12 RCTs, and stratifies by setting. It reports eight safety and efficacy outcomes (recovery time and other continuous outcomes are reported narratively), together with heterogeneity, leave-one-out sensitivity analysis, trial sequential analysis and GRADE certainty, and it quantifies the costs of fospropofol disodium (pruritus/paraesthesia; slower onset after bolus dosing).

*Clinical implications:* Fospropofol disodium may reduce injection pain during sedation. For hypertriglyceridaemia the result depends on the estimator and certainty is very low. During procedural sedation it may reduce hypotension and respiratory adverse events, whereas the evidence for a hypotension benefit during continuous ICU sedation is weak. Because the approved indication in China is adult general anaesthesia induction only, these uses remain investigational; they do not constitute a routine clinical recommendation. Applicability to specific patient groups remains an inference from pharmacological mechanism.

## 1 Introduction

### 1.1 The clinical importance of sedation

Sedation outside the operating room refers to sedation given outside the operating theatre for purposes other than general anaesthesia. It covers continuous sedation of mechanically ventilated ICU patients and sedation for procedures such as gastrointestinal endoscopy, hysteroscopy and bronchoscopy. As comfort-oriented care has expanded and the range of procedures has grown, demand for this type of sedation has risen steadily, and it is now part of routine work in anaesthesia and critical care [8]. Sedation is not a low-risk procedure. A prospective cohort study of 2132 patients undergoing gastrointestinal endoscopy sedation in nine university-affiliated hospitals reported significant adverse events in 23.0% of patients, with significant hypotension in 11.8%; risk increased with age, abnormal body mass index and higher ASA class, and 30-day mortality was 1.2% (6.0% in emergency patients) [9]. The safety of sedation is therefore closely tied to whether the procedure can be completed, and particular caution is warranted in older patients or those with poor cardiorespiratory reserve.

### 1.2 Formulation limitations of propofol

Propofol has a rapid onset, a short half-life and is easy to titrate, and it has long been a first-line sedative. Its commercial formulation is a lipid emulsion, which brings several inherent problems. First, injection pain is very common. A quantitative systematic review of 56 randomised studies and 6264 patients found that a mean of 70% of patients reported pain on injection [10]. The mechanism involves activation of the plasma kallikrein-kinin system by the lipid carrier and generation of bradykinin, which causes local venodilation and increased permeability, so that free propofol in the aqueous phase more readily reaches free nerve endings in the vessel wall (adventitia) [12, 11]. Second, the rapid rise in plasma concentration after a bolus can cause circulatory and respiratory depression [21]. Third, the lipid emulsion itself represents an exogenous lipid load. A single-centre retrospective cohort study reported that 17.4% (19/109) of critically ill adults receiving continuous infusion for at least 24 h developed triglycerides above 400 mg/dL [13]. Fourth, a small number of patients develop propofol infusion syndrome (PRIS), whose pathophysiology is thought to involve impaired mitochondrial fatty acid *β*-oxidation, damage to the electron transport chain, and blockade of *β*-adrenergic receptors and myocardial calcium channels [16, 15]. A prospective study of 1017 critically ill patients receiving infusion for at least 24 h in 11 centres reported an incidence of 1.1% [14], and the syndrome carries a poor prognosis once it occurs [16, 15]. In addition, lipid emulsions carry a risk of bacterial contamination and require additional storage precautions [26, 21].

### 1.3 Pharmacology and current clinical use of fospropofol disodium

fospropofol disodium was developed as a water-soluble prodrug to address these formulation problems. After intravenous injection it is hydrolysed by alkaline phosphatase on the surface of vascular endothelial cells, releasing active propofol and producing sedation and hypnosis [19, 21]. Because enzymatic cleavage of the phosphate ester is the rate-limiting step, the peak plasma concentration of propofol occurs about 9–15 min after dosing and rises relatively gradually [21, 18]. Because it contains no lipid emulsion, the drug may in theory reduce injection pain and, by slowing the rise in plasma concentration, reduce circulatory and respiratory depression, while avoiding exogenous lipid load and bacterial contamination. It was approved by the US Food and Drug Administration in December 2008 for sedation during monitored anaesthesia care, and was withdrawn from the US market in 2012; contributing factors included limited safety data on formaldehyde accumulation, the requirement for administration by trained anaesthesia personnel, and pharmacokinetic disadvantages such as delayed onset in the outpatient setting, which together led to poor commercial performance [21]. It was approved in China on 19 May 2021 by the National Medical Products Administration (approval number H20210017; this date is the issue date on the approval document, see the NMPA list of approval documents pending collection dated 2021-05-24), with an approved indication of induction of general anaesthesia in adults [21, 20]. Use for maintenance of anaesthesia, sedation during ICU mechanical ventilation and procedural sedation is outside the approved indication and remains investigational. Randomised trials have nevertheless been conducted in these settings and their evidence has yet to be synthesised. The pharmacological pathway is shown in Supplementary Material S6.

### 1.4 Existing evidence and the gap

A recent meta-analysis evaluated fospropofol disodium versus propofol for general anaesthesia induction and reported less injection pain and bradycardia with fospropofol disodium, but slower onset and more pruritus and numbness [23]. Induction and sedation are different clinical problems. Induction uses a single dose and aims to reach an adequate depth of anaesthesia quickly, whereas sedation requires continuous or repeated dosing, and the relevant questions are whether the target depth can be maintained, whether hypotension or respiratory depression occurs, how lipids change, and how long recovery and discharge take. Because dosing and endpoints differ, conclusions from the induction setting cannot be extrapolated directly to sedation.

After crossing drug terms with sedation-setting terms across five databases, no published systematic review or meta-analysis of fospropofol disodium for sedation outside the operating room was identified. Because the search strategy also required a co-hit on propofol, this finding means only that no such synthesis with propofol as the comparator was retrieved; it does not exclude studies that used another sedative as the comparator while also evaluating fospropofol disodium. Two related protocols are registered in PROSPERO. The provenance of those two registration numbers should be stated: PROSPERO does not provide a stable, reproducible bulk-search export, so it was used by checking the specific registration records directly related to this question (query terms fospropofol, ciprofol and the Chinese drug name), rather than as a systematic search source contributing records to screening. This is flagged in Appendix 1 and in the search records. The two records are: a meta-analysis of ciprofol and fospropofol disodium for sedation anaesthesia outside the operating room (CRD42024618153, registered December 2024) and a systematic review of the safety and efficacy of fospropofol disodium in anaesthesia and sedation (CRD42023477740, registered November 2023). As of the search date (21 September 2026), neither had a corresponding published paper. Compared with these protocols, the present review differs in several respects: (i) the comparator is restricted to propofol in a head-to-head comparison, whereas CRD42024618153 used an undefined “usual care” comparator and included ciprofol; (ii) the population covers both continuous sedation during ICU mechanical ventilation and procedural sedation, whereas that protocol covers procedural sedation only; (iii) eight efficacy and safety outcomes are assessed (hypotension, injection pain, bradycardia, respiratory depression/hypoxaemia, hypertriglyceridaemia, pruritus/paraesthesia, sedation efficacy and successful extubation; recovery time and other continuous outcomes are reported narratively), more than the four in that protocol; (iv) the search was updated to September 2026 and most included studies were published after that protocol’s search cut-off; and (v) GRADE assessment, trial sequential analysis, leave-one-out sensitivity analysis and assessment of publication bias were added, none of which formed part of those protocols. The present protocol is registered in PROSPERO (CRD420261507586).

### 1.5 Objectives and expectations

Against this background, we assessed the comparative safety and sedation-related outcomes of fospropofol disodium versus propofol in adults receiving sedation outside the operating room (ICU mechanical ventilation and procedural sedation), restricting the scope to that setting to distinguish it from the existing induction meta-analysis. We expected that the two drugs would have similar sedation efficacy, that fospropofol disodium might reduce injection pain and some cardiopulmonary adverse events, and that it might have a slower onset and cause more pruritus or abnormal sensations. These expectations do not constitute a non-inferiority hypothesis.

## 2 Methods

### 2.1 Protocol and registration

This systematic review followed PRISMA 2020 [1], the PRISMA-S reporting standard for searches, and the Cochrane Handbook [2]. The review was prospectively registered: the protocol was submitted on 15 September 2026 and the PROSPERO record became effective on 16 September 2026 (CRD420261507586). All database and registry searches were run on 21 September 2026, and data extraction, risk-of-bias assessment and all statistical analyses were undertaken after registration. Analyses carried out after the protocol was finalised are labelled as exploratory in the Results.

#### Deviations from the protocol

So that readers can distinguish pre-specified from post-hoc decisions, all deviations are listed together. (i) *Publication-bias test*: the original protocol specified Egger’s test; because the primary outcome, hypotension, is binary and the correlation between the log OR and its standard error inflates the false-positive rate of Egger’s test, this review uses the Harbord test, which is appropriate for binary outcomes, as the primary test and also reports the originally specified Egger’s test to preserve continuity with the registered protocol (see section 3.14 and Supplementary Material S6-E). (ii) *Setting strata*: the protocol planned four strata (ICU, gastroscopy, bronchoscopy, hysteroscopy); as no eligible bronchoscopy trial and only two hysteroscopy trials were available at the search date, that granularity was not feasible and the three procedural settings were combined into a single procedural stratum (see section 2.6). (iii) *Trial sequential analysis*: the assumed control risk came from this review’s own pooled data rather than an external pre-specified value, so the TSA is positioned as exploratory supporting evidence, not as a confirmatory result. (iv) *Post-hoc exploratory analyses*: stratification by blinding, exclusion of the Gao cohorts, perturbation of the preprint event counts and the estimator sensitivity analysis are each flagged in section 2.6 and in the Results. These deviations have been registered as an amendment to the PROSPERO record, and the amendment was made without access to any outcome data.

### 2.2 Scope and definitions

In this article, “sedation outside the operating room” and “sedation for purposes other than general anaesthesia induction or maintenance” are used interchangeably and refer to sedation given for purposes other than induction or maintenance of general anaesthesia, covering (i) continuous sedation of mechanically ventilated ICU patients and (ii) sedation for procedures such as gastrointestinal endoscopy, hysteroscopy and bronchoscopy. This definition was chosen so that the scope does not overlap with the 2026 meta-analysis of general anaesthesia induction [23]. ICU sedation is not classified as out-of-operating-room anaesthesia (NORA) in the narrow sense under some classification systems; we adopted the broader operational definition and discuss this limitation below.

### Outcome definitions and dosing regimens

The original studies used different thresholds, recording times and decision rules for the same outcome. For example, thresholds for hypertriglyceridaemia ranged from 1.7 to 2.3 mmol/L, some studies did not report a bradycardia threshold, and some respiratory definitions used a composite that included cough. We did not re-harmonise outcome definitions; event counts were extracted according to the definitions reported in each article. The dosing regimen of each study (dose, route, whether infusion was continuous, concomitant analgesics and the propofol comparator dose) and the operational definition of each outcome are provided as supplementary material (Supplementary Table S3, which includes the page or table of origin for every data point). This variation in definitions is itself a source of heterogeneity and underlies the inconsistency seen for hypertriglyceridaemia and bradycardia; the consequences are addressed in the GRADE downgrading reasons and in the sensitivity analyses (sections 3.12, 3.17 and Appendix 6). “Respiratory depression/hypoxaemia” is a composite respiratory adverse-event outcome defined a priori for this review rather than a single endpoint identical across trials: the included studies reported respiratory depression, hypoxaemia or closely related events, and the specific definitions and measurement windows are given in Supplementary Table S3. The composite is used for exploratory pooling only and the original definitions should be kept in view when it is interpreted.

### Choice and interpretation of the random-effects estimator

The DerSimonian-Laird method was used as the primary estimator to remain consistent with the PROSPERO protocol and the pre-specified analysis plan, not because it is optimal when the number of studies is small. Because most outcomes in this review are based on only 4 to 10 studies and zero events and large effects are not uncommon, all outcomes are also reported with REML estimation of *τ* ^2^ plus modified Hartung-Knapp confidence intervals: REML estimates *τ* ^2^, and the Hartung-Knapp adjustment provides a more conservative interval when few studies are available; following common practice for small *k*, the fixed-effect variance is retained when the Hartung-Knapp standard error falls below it (truncation). Where the two estimators change the conclusion or the significance, the result is reported as sensitive to the estimator and the evidence as uncertain, rather than selecting the more favourable estimate as definitive; hypertriglyceridaemia is such a case.

### 2.3 Eligibility criteria

**Population (P):** adults receiving sedation outside the operating room (ICU mechanical ventilation or procedural sedation/endoscopy). **Intervention (I):** fospropofol disodium. **Comparator (C):** propofol. **Outcomes (O):** the eight pre-specified outcomes were sedation efficacy (success rate or target attainment), hypotension, injection pain, bradycardia, respiratory depression/hypoxaemia, hypertriglyceridaemia, pruritus/paraesthesia and successful extubation; “respiratory depression/hypoxaemia” is a composite of respiratory depression, hypoxaemia and closely related events reported by individual trials and is also referred to in the narrative as “respiratory adverse events”; recovery times (onset, recovery and discharge) and adverse events are additionally reported narratively and were not pooled (see sections 1.4 and 3.9). **Design (S):** randomised controlled trials, including pilot trials. **Study type:** both formally published articles and preprints not yet peer reviewed were eligible; preprints (for example Liu 2026, Research Square) are labelled as not peer reviewed in the figures and tables, and the robustness of conclusions to their exclusion was assessed in sensitivity analysis. **Exclusions:** trials of general anaesthesia induction or maintenance (which belong to the existing induction meta-analysis), studies without a propofol comparator, non-randomised studies, and dose-finding studies without extractable data. **On restricting the comparator:** the comparator was required to be propofol rather than “usual care” or another sedative. This restriction was made for two reasons. First, the question is a head-to-head comparison of alternative strategies, and only a uniform comparator provides effect sizes (OR, RD and the derived NNT/NNH) that can be used directly in clinical decisions. Second, relaxing the restriction would include many dose-finding studies that use dose-to-dose or placebo comparisons, whose differences are not between-group comparisons and cannot be interpreted when pooled. This restriction also distinguishes the present review from the registered protocols on the same topic (CRD42023477740, CRD42024618153).

### 2.4 Search strategy

We searched Europe PMC, PubMed, CNKI, Wanfang and VIP (inception to 2026, no language restriction). Search strings combined drug terms with sedation-setting terms, and the full strategies are given in Appendix 1. Searching was done in two steps, with 21 September 2026 as the final search date for all databases. A narrower “drug term *×* propofol *×* setting term” strategy was used first to build the screening pool, giving Europe PMC 144, PubMed 55, CNKI 27, Wanfang 43 and VIP 69, or 338 records in total. A broadened strategy was then used to re-search those five databases and to extend the search to Embase, Cochrane CENTRAL and Web of Science Core Collection; the per-database results are given in item (vii) below and in Appendix 1. The two steps differ only in the breadth of the Chinese-language strategies: CNKI rose from 27 records (professional search with exact crossing of subject fields) to 69 (68 unique) and Wanfang from 43 records (32 retrieved) to 44 (42 unique), whereas Europe PMC (144), PubMed (55) and VIP (69) returned the same numbers in both steps. After deduplication by title, 214 records remained (327 records were retrievable and 113 were duplicates). Two reviewers screened titles and abstracts independently, excluding 200 records and leaving 14 for full-text assessment; two were excluded at full text, one because it studied general anaesthesia induction with laryngeal mask ventilation and one because it was a dose-finding study without a propofol comparator, leaving 12 RCTs (Figure 1). All event counts in the included studies were verified against the full-text original publications. Of the 43 Wanfang records, 32 were retrievable and the remaining 11 did not enter the original deduplication and screening pool; a subsequent broad Wanfang search identified no further eligible study. The reproducibility limitation of the Chinese database counts and the supplementary material inventory are described in Appendix 1.

**Figure 1:**
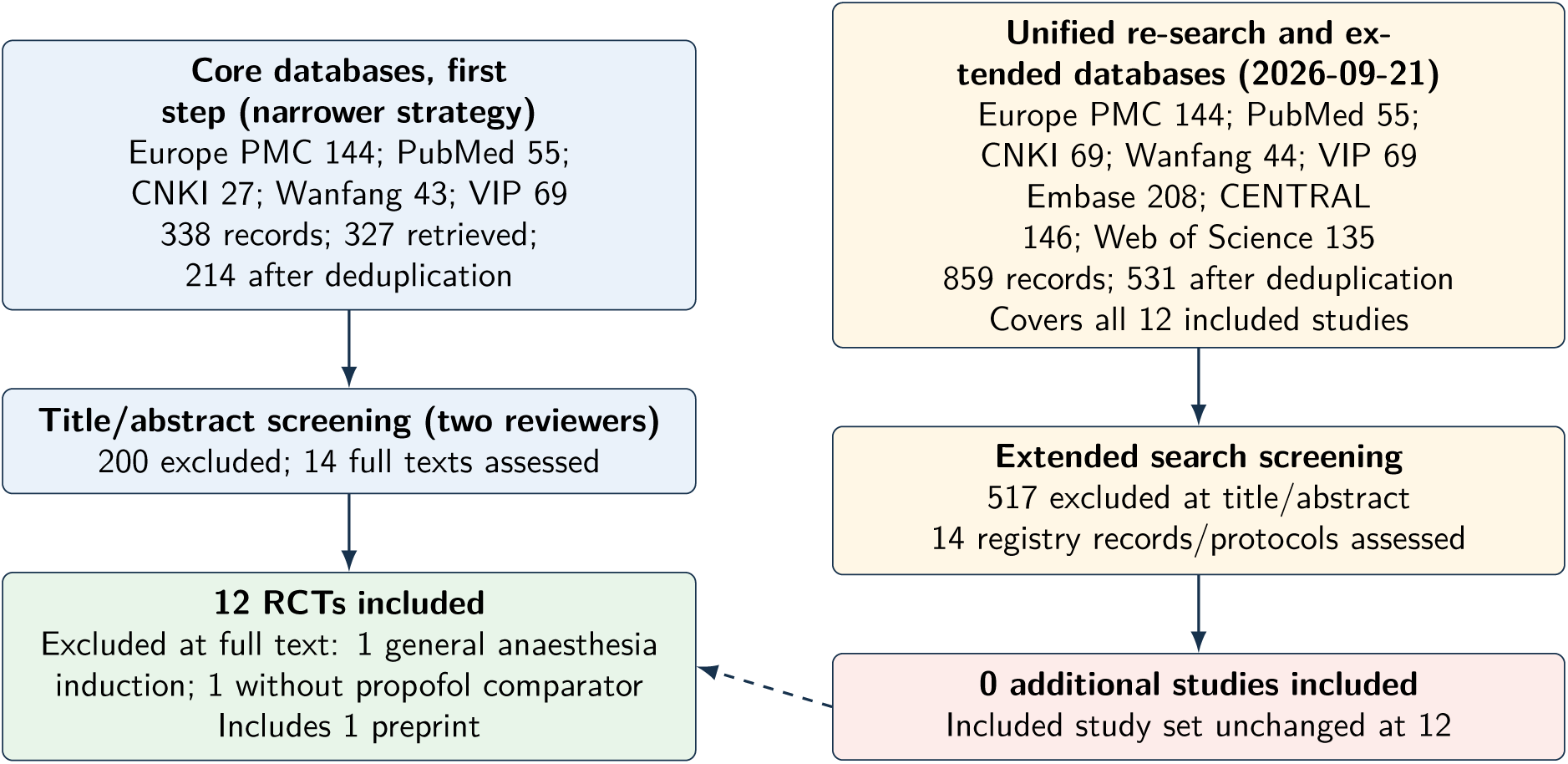
PRISMA 2020 flow diagram. Searching was done in two steps, with 21 September 2026 as the final search date. First step, core databases (narrower strategy): Europe PMC 144, PubMed 55, CNKI 27, Wanfang 43, VIP 69, with 327 records retrieved and 214 after deduplication; two reviewers excluded 200 records at title and abstract screening, assessed 14 full texts, and excluded one general anaesthesia trial and one dose-finding study without a propofol comparator at the full-text stage, leaving 12 studies. Unified re-search and extended databases (21 September 2026): Europe PMC 144, PubMed 55, CNKI 69, Wanfang 44, VIP 69, Embase 208, Cochrane CENTRAL 146, Web of Science Core Collection 135; 859 records were merged, 531 remained after deduplication, and that record set covers all 12 included studies; 517 records were excluded at title and abstract stage, and 14 registry or protocol records were assessed and all excluded, so no additional study was included and the included set remained at 12.

### Search (21 September 2026): standard databases, trial registries and conference proceedings

The registered protocol specified that both published and unpublished studies would be sought (”Both published and unpublished studies will be sought”). Beyond the five core databases, we searched ClinicalTrials.gov as specified (search term fospropofol, 33 registered trials). All searches were run after the PROSPERO record took effect. The results were handled as follows. (i) Most records corresponded to published articles whose comparators were not propofol (midazolam, placebo or dose-to-dose comparisons) and were excluded under the pre-specified criteria. (ii) One completed randomised trial, NCT01401049 (USA, *n* = 116, single-blind, primary outcome injection pain), compared fospropofol disodium with propofol head to head but is not included in this review: the comparator arm was a mixture of propofol and lidocaine, and the registry record states that the original files and patient data were lost because of a hurricane, so no extractable outcome data were provided. The reasons are recorded in Appendix 4. (iii) One ongoing trial, NCT07659132 (fospropofol disodium versus propofol for ureteroscopic anaesthesia, *n* = 190), may be eligible for a future update. (iv) We also searched the WHO International Clinical Trials Registry Platform (ICTRP) (search term fospropofol, 140 registered trials; this platform aggregates national registries and includes records from the Chinese Clinical Trial Registry (ChiCTR) that are not indexed in ClinicalTrials.gov). Most of the 140 records concerned general anaesthesia induction or maintenance and were outside the scope of this review; among 41 records related to sedation, no additional extractable outcome data were found beyond the included studies, so the included set was unchanged. (v) We additionally searched two Chinese sources: SinoMed (search term “fospropofol”, 35 records) and Wanfang (614 records, of which about 98 were relevant); no additional eligible study was identified. Most records concerned general anaesthesia induction or maintenance, dose-finding studies, animal experiments, analytical methodology or reviews and were excluded under the pre-specified criteria (see supplementary material). (vi) We also searched the CNKI and Wanfang conference proceedings (terms included fospropofol, fospropofol sodium, Lusedra, Aquavan and GPI-15715; CNKI was searched by subject and by title, Wanfang was restricted to conference proceedings). After deduplication, only three conference records genuinely concerned this drug, covering an assay methodology, a phase I tolerability study in healthy volunteers and a mouse experiment on analgesic interaction; none met the criteria of adult sedation outside the operating room with a propofol comparator, so no additional eligible RCT was identified and the included set was unchanged. SinoMed did not index the three Chinese studies included here (Xiong 2026, Zhan 2025 and Fan 2024), which indicates incomplete coverage of Chinese literature on this topic and shows that a single Chinese database is not sufficient for a complete search. (vii) Extended standard-database search. On 21 September 2026 we reran Europe PMC (144), PubMed (55), CNKI (69, 68 unique), Wanfang (44, 42 unique), Embase (208, 200 unique), Cochrane CENTRAL (146) and Web of Science Core Collection (135). After merging and deduplication using normalised titles with DOI/PMID priority, 531 unique records remained; 517 were excluded at title and abstract stage, and the remaining 14 were registry records, protocols or registration-only reports that were assessed further and then excluded because no published results were available, the setting did not match, or there was no propofol comparator. No additional published RCT entered full-text extraction. The additional records mainly concerned general anaesthesia induction or maintenance, dose-finding studies without a propofol comparator, reviews and conference abstracts, and ongoing or unpublished registered trials. VIP was searched on 21 September 2026 and returned 69 records that entered the merged deduplicated set. The merged record set from this unified re-search covers all 12 studies included in this review (record-by-record coverage check in the “search records” package of the supplementary material), so the final included set does not depend on the narrower first-step strategy; the extended search did not change the set of 12 included studies, although several potentially eligible trials are recruiting or registered without results. These coverage limitations and the need for updates are discussed below.

### 2.5 Study selection and data extraction

Two reviewers screened and extracted data independently, resolving disagreements by discussion or third-party adjudication. Extracted items were study characteristics, population, dosing regimen, outcome definitions and values (binary event counts and numbers analysed). Risk of bias was assessed with Cochrane RoB 2 across five domains [3]. In addition to the study-level overview, risk of bias was reassessed for four key outcomes (hypotension, injection pain, respiratory adverse events and pruritus/paraesthesia): the randomisation, deviations from intended intervention, missing outcome data and selection of the reported result domains followed the two-reviewer adjudication, while the measurement of the outcome domain was re-judged according to whether the outcome was subjective or objective, and the overall judgement was recalculated accordingly. Outcome-specific results are shown in Supplementary Table S4. Event counts for all included studies were verified against the full-text original publications; the verification procedure and results are described in Appendix 5.

### 2.6 Statistical synthesis

The primary outcome was hypotension; injection pain, bradycardia, respiratory depression/hypoxaemia, hypertriglyceridaemia, pruritus/paraesthesia, sedation efficacy and successful extubation were secondary outcomes. Binary outcomes are expressed as OR with 95% CI. All binary outcomes were pooled with a DerSimonian-Laird random-effects model as the primary analysis [5], with REML plus modified Hartung-Knapp results reported alongside as an estimator sensitivity analysis; where the two changed the conclusion, the positive DerSimonian-Laird result was not treated as definitive. A 0.5 continuity correction was applied only when a study’s 2 *×* 2 table genuinely contained a zero cell; a study with one event in an arm but no zero cell was not corrected. For example, Liu 2026 reported pruritus/paraesthesia as 30*/*213 versus 1*/*213, which contains no zero cell, so the study-specific OR is 34.75; 0.5 was added only for the zero-event comparator arms of Tang 2026 and Fan 2024. Consistent with the registered protocol (CRD420261507586), three outcomes with a high proportion of zero cells (injection pain, respiratory depression/hypoxaemia and pruritus/paraesthesia) are also reported with a Peto OR as a robustness analysis; the zero cells were distributed as follows: 3 of 8 studies had no events in the fospropofol disodium arm for injection pain, 1 of 5 for respiratory depression/hypoxaemia, and 2 of 4 had no events in the propofol arm for pruritus/paraesthesia. Peto was not used as the primary estimator because it assumes an effect close to the null (OR *≈* 1) and similar arm sizes, and it is biased towards the null when the effect is large or the arms are very unbalanced. This applies to pruritus/paraesthesia (all study-specific ORs above 11, with Fan 2024 at 126:34 and Tang 2026 at 52:29) and to injection pain, where the Peto estimate is clearly lower than the study-specific values and the crude pooled estimate that requires no correction. Both sets of estimates are therefore reported so that readers can judge for themselves. Heterogeneity was quantified with *I*^2^ and *τ* ^2^ and stratified into ICU and procedural sedation layers. Under the registered protocol, four setting strata were originally planned (ICU, gastroscopy, bronchoscopy and hysteroscopy). In the included studies, no eligible bronchoscopy trial was available as of the search date (one fospropofol disodium study in this setting, a master’s thesis on painless fibreoptic bronchoscopy in older patients, explored the dose-response of fospropofol disodium combined with alfentanil without a propofol comparator and was excluded under the pre-specified criteria; a randomised trial of fospropofol disodium versus propofol in this setting is recruiting (ChiCTR2600128516) and had no results at the search date), and only two hysteroscopy trials were available, so that level of granularity was not methodologically feasible.

The three procedural settings (gastroscopy, bronchoscopy and hysteroscopy) were therefore combined into a single procedural sedation layer analysed alongside the ICU layer; this decision was made on the basis of data availability and was not fully pre-specified. Within-stratum estimates and the between-stratum interaction test (*Q_bet_*) used the same model as the primary analysis, that is, random effects for all outcomes; the stratum subtotals in the forest plots follow from this. *Q_bet_* was computed by inverse-variance weighting of the stratum-specific pooled estimates *θ*^^^*g* and their standard errors *se_g_*: *Q_bet_* = ∑_g_ (*θ*^^^*_g_ − θ*^^^*·*)^2^*/se*^2^_g_, where *θ*^^^*·* = (∑_g_ *θ*^^^*_g_/se*^2^) / (∑_g_ 1/se^2^_g_) and the degrees of freedom are the number of strata minus one; when a stratum contains a single study, the within-stratum “pooled” estimate is that study itself and the test is not methodologically valid. The primary outcome is also reported as a risk difference (RD) with the derived NNT/NNH; RD used the same continuity correction as the primary OR analysis for studies with zero cells, so its values do not correspond exactly to the Peto estimates and should be interpreted with that in mind.

The following pre-specified sensitivity analyses were performed: (i) leave-one-out analysis; (ii) exclusion of the preprint; and (iii) handling of the dose arms of multi-arm trials. Four additional exploratory analyses are reported and labelled as such in the Results: (iv) trial sequential analysis (TSA; using the pooled control event rate in this review, 19.2%, as the assumed control risk, with a 25% relative risk reduction, two-sided *α* = 0.05 and 80% power; because that assumed control risk comes from this review’s own pooled data rather than an external pre-specified value, the result is interpreted as exploratory only); (v) a descriptive comparison by blinding status (double-blind studies versus the remaining open-label or undescribed studies); (vi) because Gao 2024 and Gao 2025 may come from adjacent cohorts at the same institution, sensitivity analyses for hypertriglyceridaemia and for hypotension excluding either study; and (vii) a two-arm reverse perturbation of the Liu 2026 preprint event counts.

### Publication-bias test

The original protocol specified Egger’s test; because the primary outcome, hypotension, is binary and the log OR and its standard error are calculated from the same event counts and are therefore correlated, Egger’s test (which derives from continuous outcomes) has an inflated false-positive rate in this setting. This review therefore uses the Harbord test, based on the efficient score, as the *primary* publication-bias test; the originally specified Egger’s test is still reported for continuity with the original protocol and earlier literature, but its result is presented for comparison only and is not the primary basis for interpretation (the deviation is listed in section 2.1; see section 3.14 and Supplementary Material S6-E). Continuous outcomes were pooled according to the number of studies, the clinical construct and heterogeneity; onset or loss-of-consciousness time was not pooled quantitatively because only two studies reported it and *I*^2^ *>* 90%, and recovery and discharge times were pooled as randomeffects mean differences. Certainty of evidence was assessed with GRADE [4]. Analyses used Python 3.11.2 (numpy 2.4.6, scipy 1.17.1, matplotlib 3.11.1); pooled estimates used DerSimonian-Laird and Peto implementations of the fixed formulas, and *τ* ^2^ for the random-effects model was obtained by the method of moments. The estimator sensitivity analysis was computed by a separate script (estimator_check.py) that implements REML *τ* ^2^ by numerical maximisation of the REML log-likelihood together with modified Hartung-Knapp intervals, reporting both the truncated and untruncated intervals. No random numbers were used, so no seed was set. The analysis scripts and data files are provided with the manuscript as supplementary material (see the data availability statement).

## 3 Results

### 3.1 Study selection and characteristics

Twelve RCTs were included (Figure 1), all with a propofol comparator, and all contributed to the quantitative synthesis. Zhang Yi 2026 was a dose-finding study in which all three sex subgroups received fospropofol disodium without a propofol comparator; it did not meet the requirement for a propofol comparator and was therefore excluded, with the reasons recorded in Appendix 4. Fan 2024 was a five-arm dose-finding trial (four fospropofol disodium arms plus one propofol arm), but binary safety data were extractable from the propofol arm, so the fospropofol disodium arms were combined and included in the quantitative synthesis. The studies came from China (11) and the USA (1), sample sizes ranged from 60 to 426, and settings covered ICU mechanical ventilation, painless gastroscopy or gastrointestinal endoscopy, and hysteroscopy. Liu 2026 was a five-centre RCT preprint (*n* = 426) and the largest included study. Regarding blinding, only Liu 2026 (patients and endoscopists blinded) and Fan 2024 (randomised, controlled, double-blind) reported a double-blind design; the remainder were open-label, single-blind (blinding target unspecified) or did not describe blinding (Table 1). Binary event counts extracted study by study are shown in Table 2.

**Table 1:** Characteristics of the 12 included studies.

| Study | Setting | Country | n | Blinding | Key outcomes reported |
| --- | --- | --- | --- | --- | --- |
| Feng 2026 [25] | ICU, goal-directed sedation | China | 210 | Open (no blinding) | Hypotension, bradycardia, hypertriglyceridaemia, delirium |
| Gao 2025 [26] | ICU, deep sedation | China | 60 | Open (no blinding)<sup>g</sup> | Hypotension, bradycardia, hypertriglyceridaemia, extubation |
| Gao 2024 [27] | ICU, long-term sedation | China | 60 | Open (not blinded) | Hypotension, bradycardia, hypertriglyceridaemia, target attainment time |
| Candiotti 2011 [28] | ICU sedation | USA | 60 | Open (no blinding) | Hypotension, bradycardia, adverse events |
| Xiong 2026 [29] | ICU sedation after tracheal intubation | China | 62 | Not described | Hypotension, bradycardia, hypertriglyceridaemia, extubation |
| Bai 2025 [31] | Sedation for gastrointestinal endoscopy | China | 136 | Not described | Hypotension, bradycardia, injection pain |
| Zhan 2025 [32] | Sedation for painless gastroscopy | China | 180 | Not described | Injection pain, body movement, haemodynamics |
| Xu 2025 [33] | Sedation for painless gastroscopy | China | 80 | Not described | Respiratory depression, injection pain, hypotension |
| Tang 2026 [34] | Sedation for hysteroscopy | China | 81 | Not described | Injection pain, pruritus/abnormal sensation, hypotension |
| Zhang 2025 [35] | Sedation for gynaecological hysteroscopy | China | 92 | Not described | Hypotension, bradycardia, injection pain, respiratory depression |
| Fan 2024 [37] | Sedation for gastroscopy (five arms, dose-finding) | China | 160 | Double-blind | Injection pain, pruritus, respiratory depression, onset time |
| Liu 2026 [38] | Sedation for gastrointestinal endoscopy (5 centres) | China | 426 | Double-blind (patients and endoscopists) | Hypotension, hypoxaemia, injection pain, paraesthesia |

Supplementary Material S6 shows the data coverage for each outcome: hypotension 10 studies, injection pain 8, bradycardia 9, respiratory adverse events 5, hypertriglyceridaemia 4, pruritus/paraesthesia 5 (in which both arms of Gao 2024 had zero events and were not pooled, so the pooled analysis for that outcome is based on 4 studies) and sedation efficacy 3 (a narrative outcome that was not pooled, so the figure marks it as *data reported* rather than *events counted*); successful extubation (2 studies) is shown in the extubation forest plot in S6. The figure makes the density of evidence and the gaps for each outcome visible at a glance.

### 3.2 Risk of bias

Overall, the included studies raised “some concerns” (Figure 2), most often because of open-label design or inadequate description of blinding and, in some studies, incomplete prospective registration; methodological reporting in the Chinese-language journal articles was relatively limited. For the key outcomes, injection pain and pruritus/paraesthesia are susceptible to subjective judgement and caregiver behaviour, whereas hypotension and respiratory depression are objectively measured, although randomisation, data completeness and the treatment context may still differ. Only 2 of the 12 pooled studies used a double-blind design, which is the most fundamental threat to validity in this review; its influence is quantified in the sensitivity analyses (section 3.12). The largest study, Liu 2026 (*n* = 426), was at low risk in the randomisation, deviations from intended intervention, missing outcome data and measurement of the outcome domains (it reported central randomisation, no loss to follow-up, and blinding of both patients and assessors), but the selection of the reported result domain was judged to raise some concerns because the article is a preprint without an accessible statistical analysis plan.

**Figure 2:**
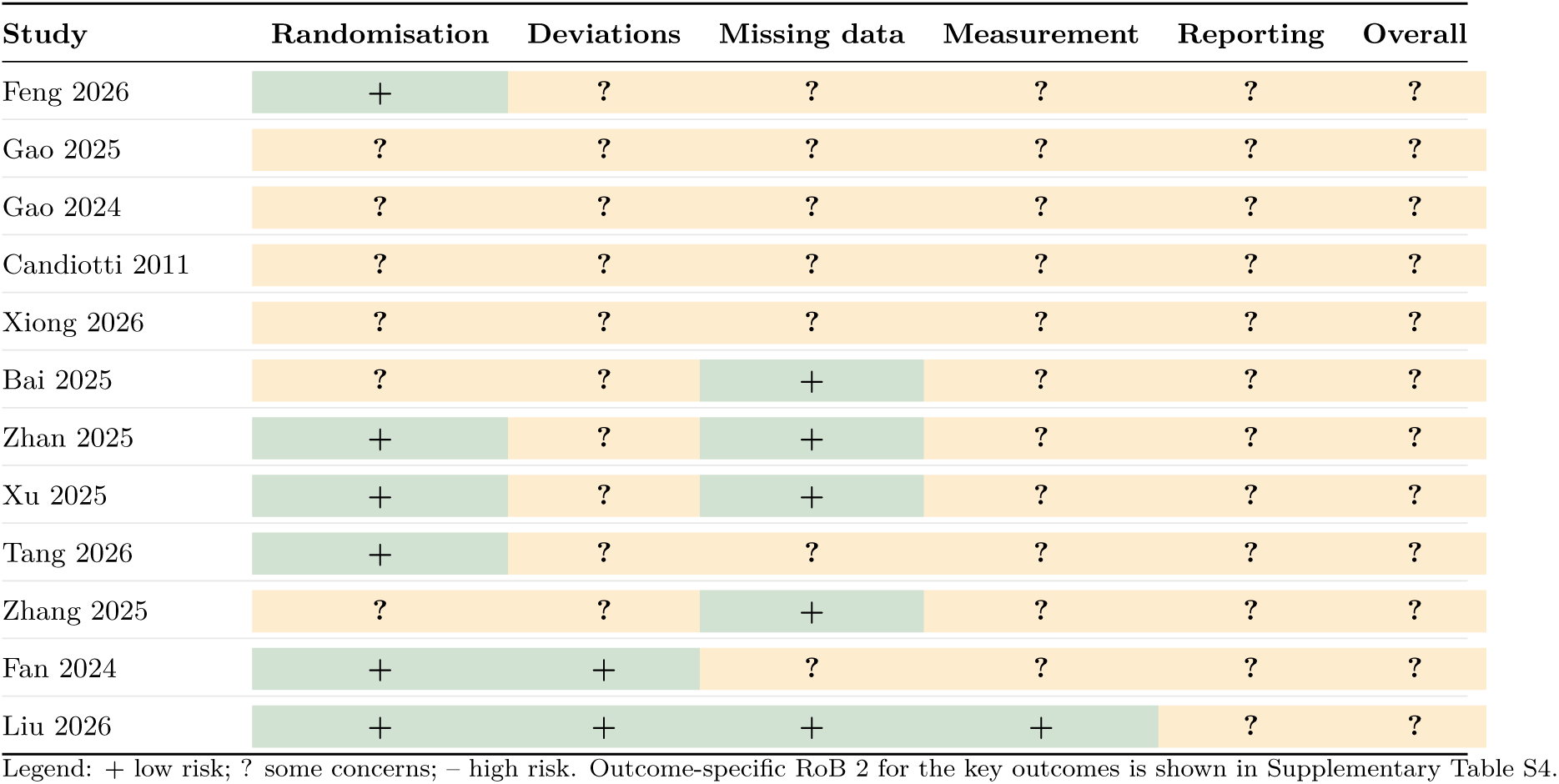
Study-level RoB 2 overview for the 12 included studies. Outcome-specific RoB 2 for the four key outcomes is shown in Supplementary Table S4. Disagreements between reviewers were resolved to the more conservative judgement; the complete domain-by-domain and study-by-study judgements with their rationale are provided in the supplementary material.

Three points about the risk-of-bias assessment should be noted. (i) Assessment approach: RoB 2 is in principle applied outcome by outcome, whereas we produced one overall judgement per study, anchored to the primary outcome (hypotension), and recorded the five domains and the signalling questions within each domain in the supplementary RoB table. Because most outcomes within a study came from the same randomisation and blinding arrangements, study-level and outcome-level judgements differed only to a limited extent, but this simplification remains a methodological limitation. (ii) Information sources: risk of bias for all 12 included studies was assessed from the full-text original publications (including the publishers’ full-text PDFs for the two English-language studies that were previously available only as abstracts); the source used for each study is flagged study by study in the scoring table. (iii) Traceability: the ratings in Figure 2 and Table 1 were not generated automatically from the raw data but were determined by two reviewers independently from the sources described above and then adjudicated; the signalling-question level scoring, the two reviewers’ original ratings and the disagreement record are provided in the supplementary RoB table. Given these three constraints, the risk-of-bias ratings should be read as a directional judgement rather than a precise outcome-specific risk estimate.

### 3.3 Hypotension

The hypotension outcome in this review uses a combined definition in which any event meeting the threshold reported by a trial during the study period is counted, and 10 studies (1197 participants) were included: Gao 2024 was added (14/30 versus 12/30, using the same definition as its companion study Gao 2025), and Candiotti 2011 was switched to the treatment-emergent adverse event (TEAE) count from its Table 3 (6/38 versus 2/22) so that it matches the threshold-based counting used elsewhere. The same article also reports a serious adverse event count of 1/38 versus 1/22 under a definition requiring a systolic blood pressure of 90 mmHg or less plus medical intervention; because that definition differs, it is reported below with the intervention-requiring and vasopressor-requiring data.

**Table 2:**
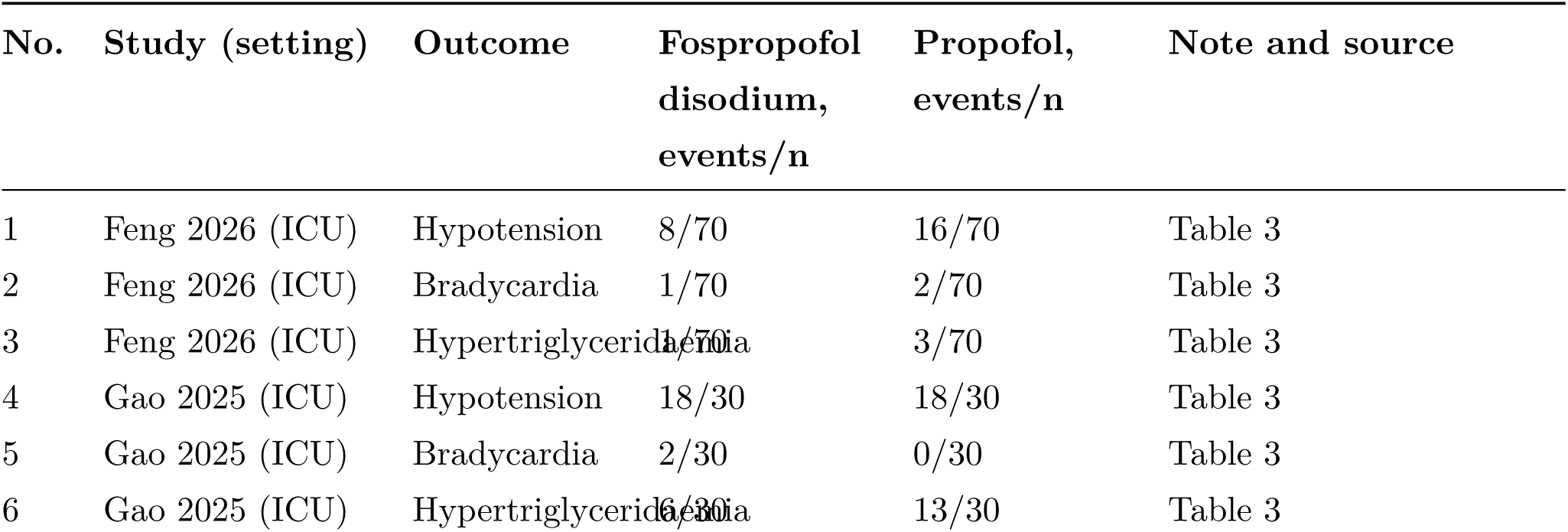
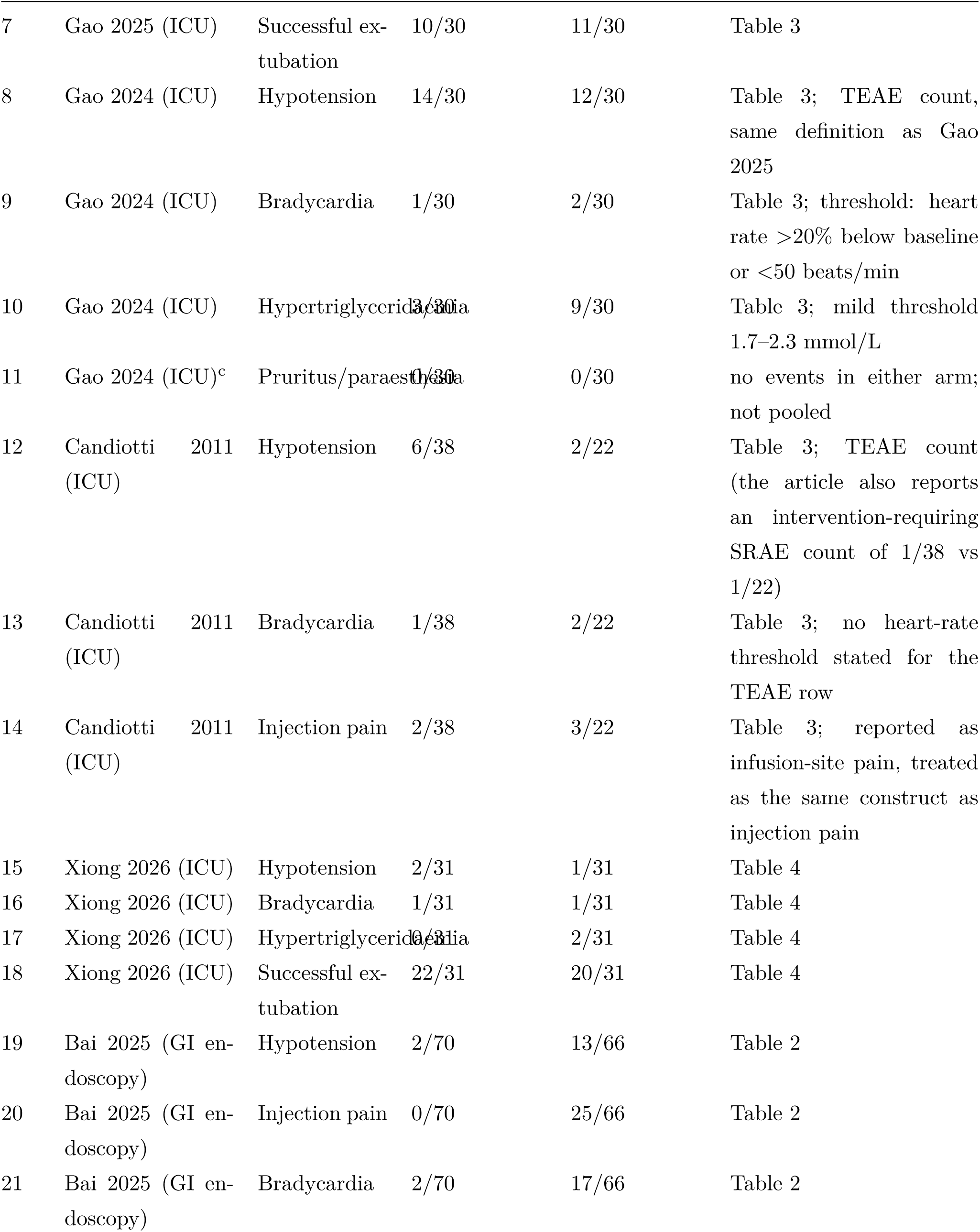
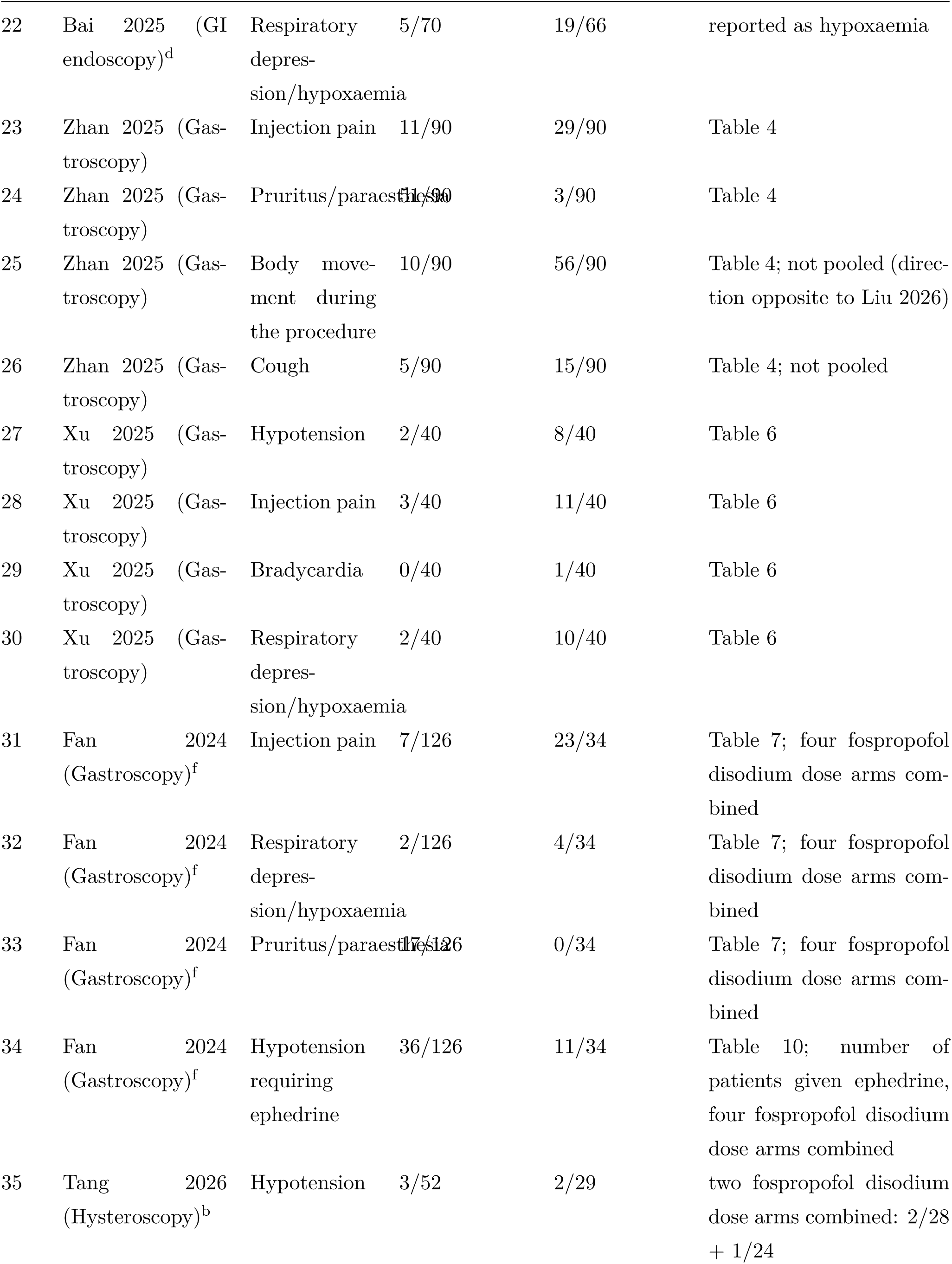
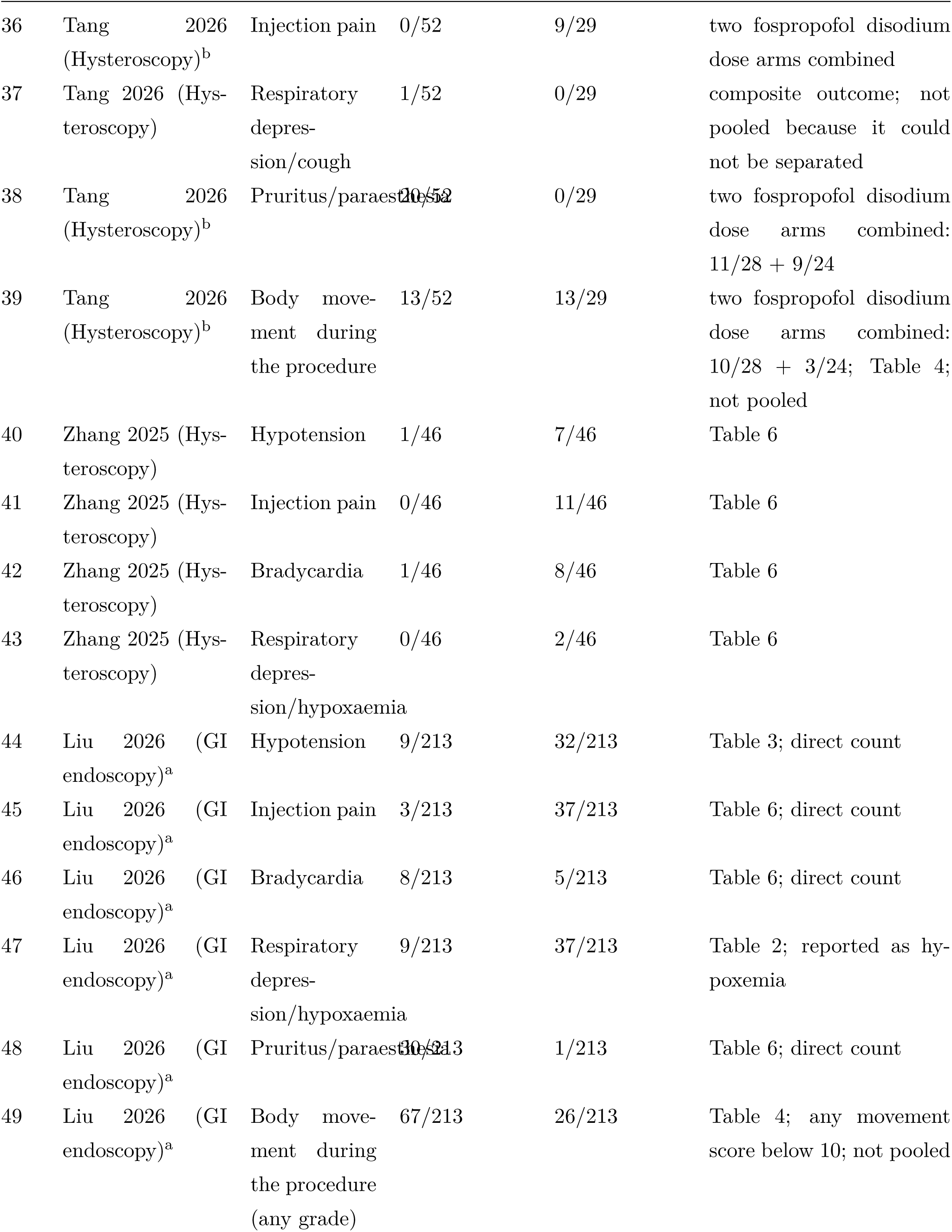
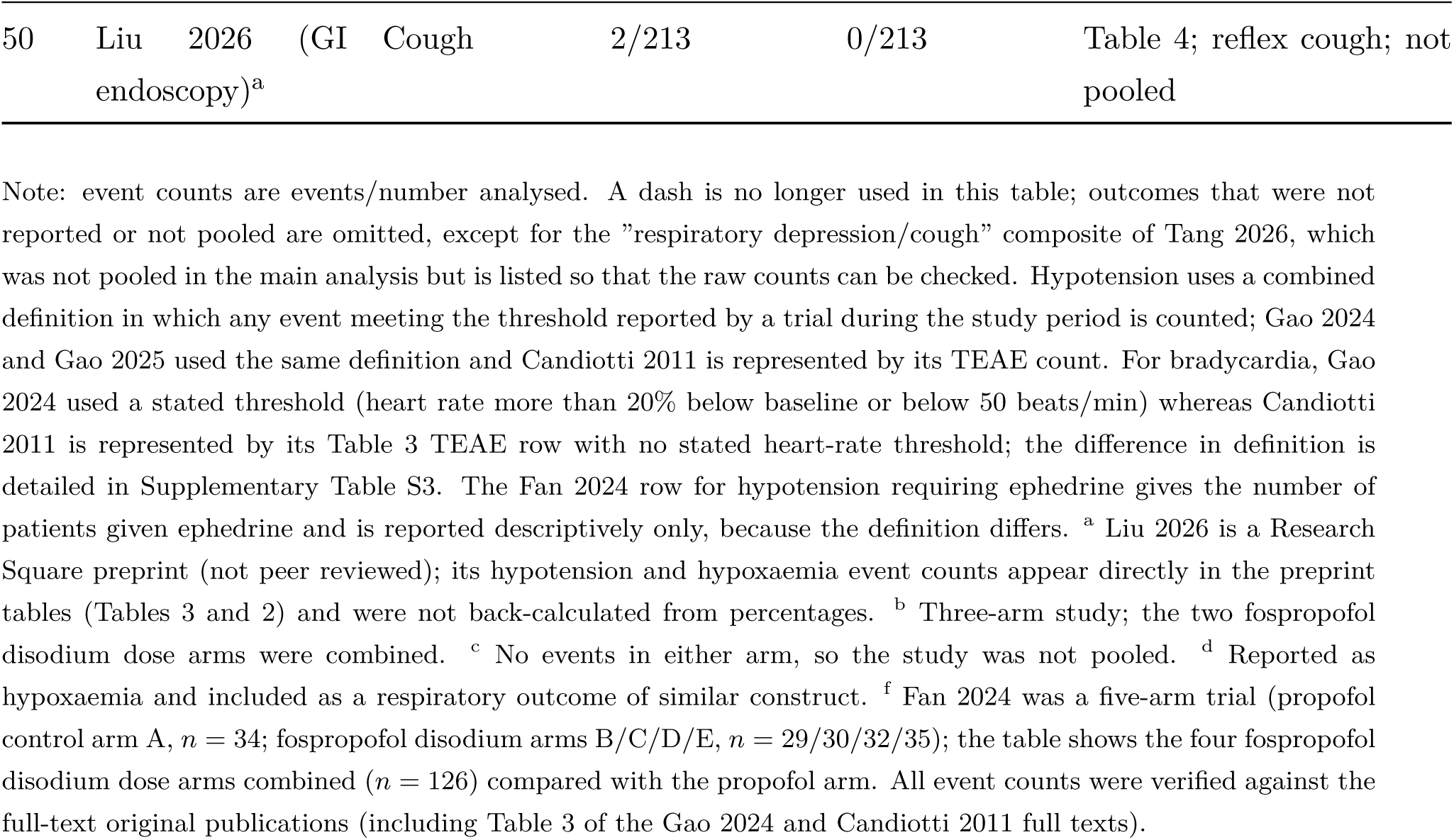
Binary event counts by study and outcome (fospropofol disodium versus propofol).

**Table 3:** Threshold, recording window and intervention requirement for hypotension in each study (10 studies).

| Study | Threshold | Recording window | Intervention required | Events (fospropofol/propofol) |
| --- | --- | --- | --- | --- |
| Feng 2026 | BP <90/60 mmHg | during the study (specific window not reported) | No | 8/70 vs 16/70 |
| Gao 2025 | MAP fall >20% from baseline or MAP <60 mmHg | during study drug infusion, up to 48 h | No | 18/30 vs 18/30 |
| Gao 2024 | MAP fall >20% from baseline or MAP <60 mmHg | during study drug infusion | No | 14/30 vs 12/30 |
| Candiotti 2011 | TEAE count in Table 3 (no BP threshold given) | during the study | No | 6/38 vs 2/22 |

| Study | Threshold | Recording window | Intervention re-<br>quired | Events<br>(fospropofol/propofol) |
| --- | --- | --- | --- | --- |
| Xiong 2026 | no operational threshold reported | during the study | Not reported | 2/31 vs 1/31 |
| Bai 2025 | non-invasive SBP <80 mmHg or >30% below baseline | intraoperative | ephedrine 6–10 mg at threshold | 2/70 vs 13/66 |
| Zhang 2025 | no threshold reported | before anaesthesia, 3 min after, at dilation, at end | Not reported | 1/46 vs 7/46 |
| Tang 2026 | no threshold reported | before induction, 1 and 5 min after, at cervical dilation, at end | Not reported | 3/52 vs 2/29 |
| Xu 2025 | MAP fall >20% | before induction to end of procedure, including recovery | ephedrine 6 mg at threshold | 2/40 vs 8/40 |
| Liu 2026 | any of: MAP fall >40% and MAP <70; MAP <60; 30% below baseline; SBP <90 | 1–5 min after induction (primary window); 9 further perioperative time points reported | ephedrine for hypotension | 9/213 vs 32/213 |

Note: this table matches Supplementary Table S3, section B1b. Four studies did not report an operational threshold, and the primary definition window in Liu 2026 (1–5 min after induction) is considerably narrower than in the other studies. Gao 2024 and Gao 2025 used the same definition. Candiotti 2011 also reports a serious adverse event count under a definition requiring a systolic blood pressure of 90 mmHg or less plus medical intervention (1/38 vs 1/22); because that definition differs it is not included in the event counts here and is reported with the intervention-requiring data instead.

Hypotension should be interpreted first by setting rather than by the overall mean. During procedural sedation fospropofol disodium reduced the risk of hypotension (5 studies; OR 0.24, 0.13–0.42; *I*^2^ = 0%), whereas during continuous ICU sedation there was no difference (5 studies; OR 0.90, 0.53–1.55; *I*^2^ = 2%); the between-setting interaction was *Q_bet_* = 11.25, *df* = 1, *P <* 0.001 (Figure 3; the reported thresholds, windows and intervention requirements are given in Table 3). The overall pooled estimate was OR 0.51 (95% CI 0.28–0.93; *I*^2^ = 51%, *τ* ^2^ = 0.44; Figure 3), with a risk difference of *−*7.2% (95% CI *−*12.3% to *−*2.1%) corresponding to an NNT of about 14, and a Peto OR of 0.46 (0.33–0.65). This overall estimate depends on the method used: REML plus modified Hartung-Knapp gave 0.51 (0.25–1.03), with a confidence interval crossing 1, and the 95% prediction interval was 0.09–2.72. We therefore do not use the overall estimate as the basis for a claim that the drug lowers blood pressure; the clinical interpretation rests on the setting-specific results, and neither estimate should be extrapolated to mechanically ventilated ICU patients.

**Figure 3:**
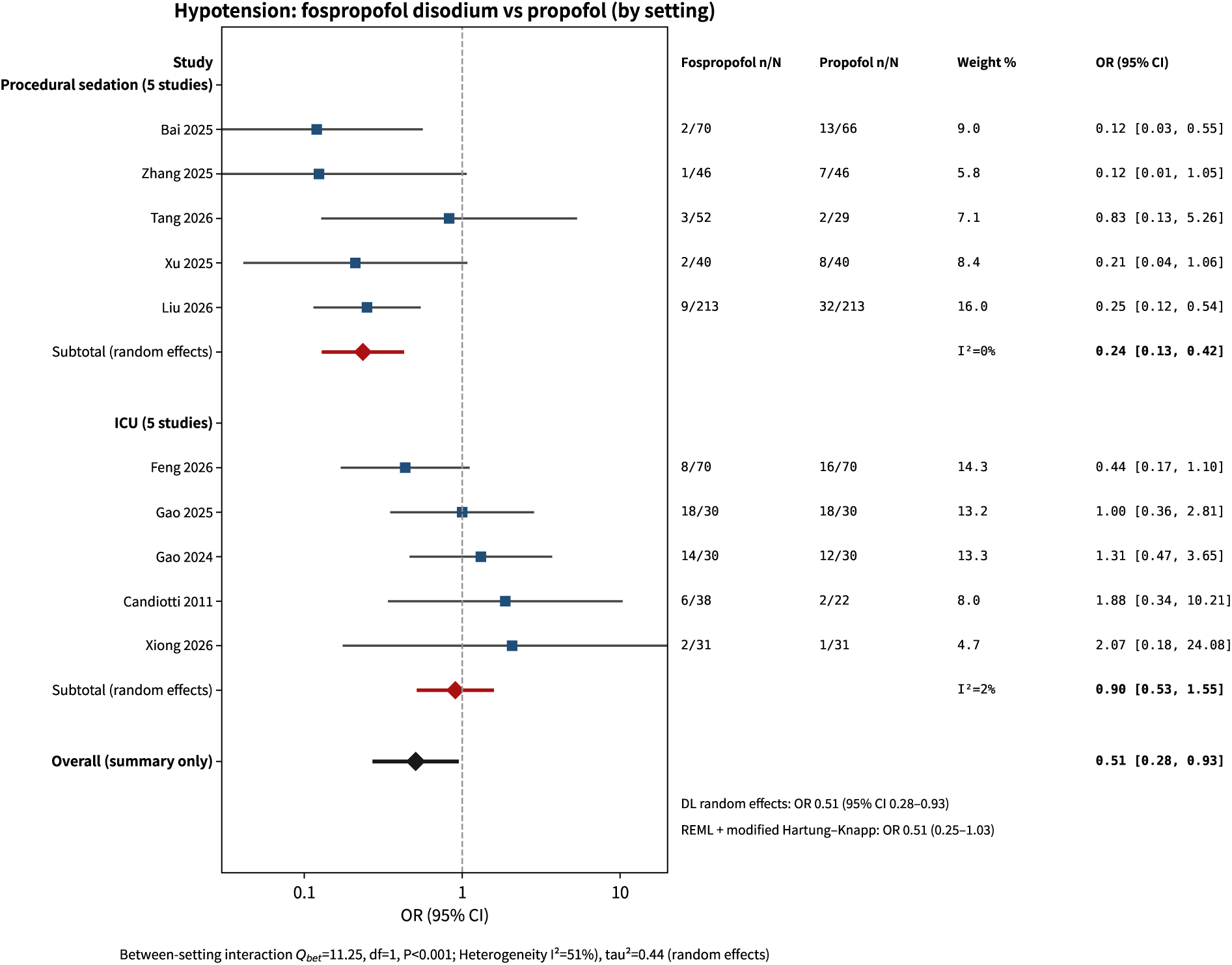
Setting-stratified forest plot for hypotension (random-effects model). Studies are grouped into procedural sedation and ICU strata, with events/number analysed, random-effects weight, and the study-specific OR with 95% CI, followed by a stratum subtotal and a bottom overall row (summary only). The overall row also shows the REML plus modified Hartung-Knapp interval (0.25–1.03, crossing 1); the between-setting interaction is *Q_bet_*= 11.25, *df* = 1, *P <* 0.001.

#### Hypotension requiring intervention or a vasopressor

Only 2 studies reported such counts, and their definitions differ, so they were not pooled: Fan 2024 reported that 36/126 (28.6%) versus 11/34 (32.4%) of patients needed additional ephedrine (5–10 mg), and Candiotti 2011 reported hypotension requiring medical intervention in 1/38 versus 1/22. A further 2 ICU studies reported vasopressor use without event counts: Gao 2024 reported a norepinephrine infusion lasting 24.00 [5.75, 37.00] h versus 24.75 [2.25, 39.88] h (*P* = 0.911) at a dose of 0.20 *±* 0.05 versus 0.15 *±* 0.04 *µ*g/kg/min (*P* = 0.356), and Gao 2025 reported no difference in the duration or dose of vasopressors. These data suggest that where the need for intervention was reported, the two groups were similar, and this should be taken into account when the difference in hypotension incidence is interpreted (Supplementary Table S3).

### 3.4 Injection pain

Eight studies reported injection pain. The overall incidence was lower with fospropofol disodium than with propofol (3.9% versus 27.4%). Pooling with the registered random-effects model gave OR 0.09 (95% CI 0.03–0.21; *I*^2^ = 65%, *τ* ^2^ = 0.99; Figure 4), with a risk difference of *−*26.9% (*−*37.0% to *−*16.7%) and an NNT of about 4 (3 to 6). Three studies reported no injection pain events in the fospropofol disodium arm (Bai 2025, Zhang 2025 and Tang 2026), and the pooled estimate is sensitive to how zero cells are handled: the Peto method gave 0.12 (0.09–0.17), and restricting the analysis to the five studies with observed events (Zhan 2025, Xu 2025, Liu 2026, Fan 2024 and Candiotti 2011) gave 0.13 (0.05–0.35) with *I*^2^ = 74%. The crude pooled OR, which requires no continuity correction, was 0.11, close to the random-effects estimate. Once the full text of Candiotti 2011 was obtained, its Table 3 row for infusion-site pain (2/38 versus 3/22) was included as the same construct. Injection pain with propofol is caused by the lipid emulsion irritating the vein wall, so infusion-site pain is the same clinical phenomenon; because that study used a continuous ICU infusion, the original article recorded it as infusion-site pain, and this review treats it as an equivalent endpoint for injection pain. The difference in source labelling is noted in Supplementary Table S3. All three approaches point in the same direction and suggest a stable benefit for injection pain, although the magnitude cannot be estimated precisely when zero events are present.

**Figure 4:**
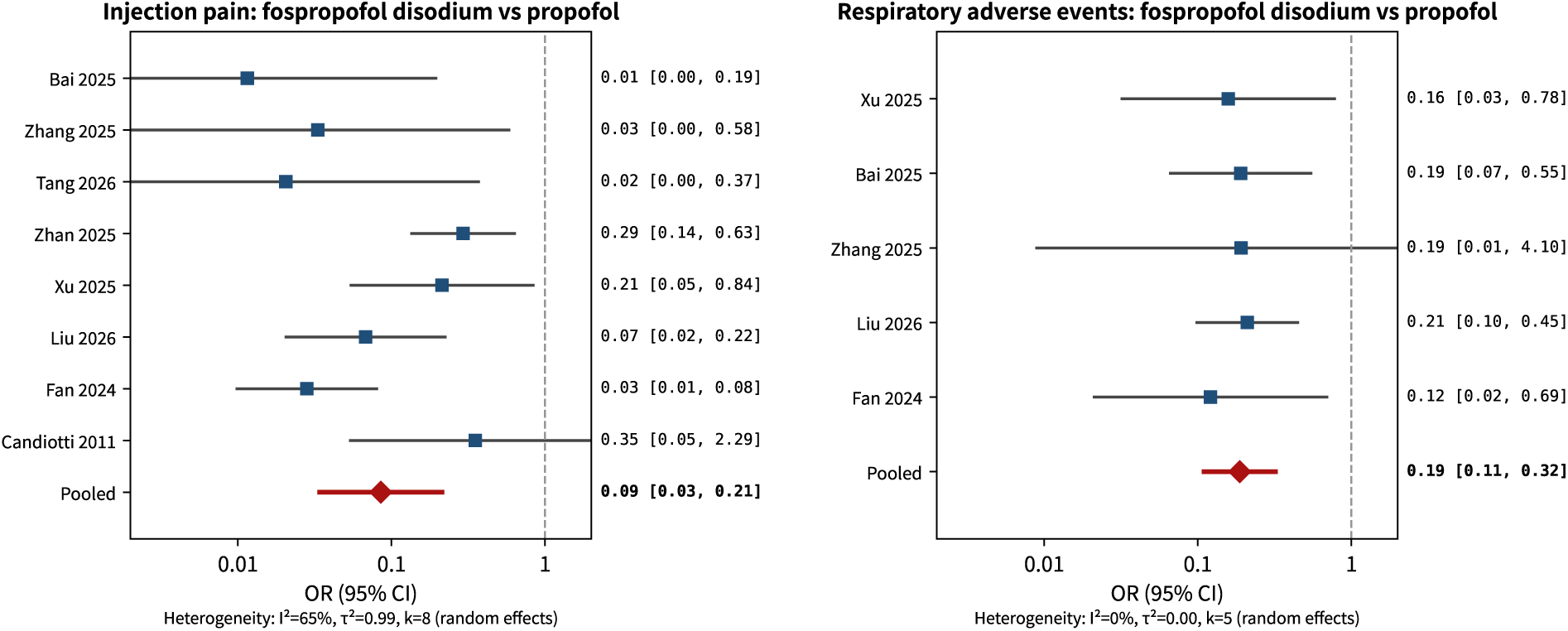
Forest plots for injection pain and respiratory adverse events (random-effects model). Left: injection pain (8 studies); right: respiratory adverse events (5 studies). The area of each square does not represent weight.

### 3.5 Bradycardia

Nine studies reported bradycardia (55 events in total); the counts for Gao 2024 and Candiotti 2011 were added in this version once their full texts were obtained. With the pre-specified randomeffects model, the risk of bradycardia was directionally lower with fospropofol disodium but did not reach statistical significance (OR 0.45, 95% CI 0.17–1.18; *I*^2^ = 44%, *τ* ^2^ = 0.89; RD *−*3.8%, *−*8.3% to +0.8%; Supplementary Material S6). The Peto method gave a smaller point estimate that was statistically significant (0.41, 0.24–0.72). The difference between the two methods has a statistical basis: Peto is a fixed-effect method and is biased towards the null when the arms are very unbalanced, whereas this outcome still shows moderate heterogeneity (*I*^2^ = 44%, *τ* ^2^ = 0.89) and the random-effects model widens the pooled confidence interval accordingly. Leave-one-out analysis located the source of heterogeneity: excluding Bai 2025 reduced *I*^2^ from 44% to 11% and changed the pooled OR from 0.45 to 0.70, the largest change of any study, while excluding Liu 2026 reduced *I*^2^ to 12% and changed the OR to 0.29. Heterogeneity therefore comes mainly from Bai 2025 rather than Liu 2026. The control-group incidence of bradycardia in Bai 2025 was 25.8% (17/66), far above the 0% to 17.4% in the other eight studies (most below 10%), and the reliability of that value remains to be confirmed against the original article: whether the threshold and recording time for bradycardia in that study match the other studies could not be confirmed here. The pooled result for this outcome therefore largely reflects a single study and interpreted with particular caution. Among the nine studies, only Bai 2025 (70 versus 66) had a slight imbalance between arms; the other studies had equal or nearly equal arm sizes. The two added studies also introduce a comparability limitation: Gao 2024 stated an explicit threshold (heart rate more than 20% below baseline or below 50 beats/min), whereas the Candiotti 2011 count comes from the treatment-emergent adverse event row of Table 3 with no stated heart-rate threshold, which contributes to the heterogeneity of the outcome definition and is flagged in the footnote to Table 2 and in Supplementary Table S3. Because this outcome is not sparse (55 events), sparse-event methods do not apply and the random-effects estimate is preferred. The conclusion is that there is no significant difference and that the evidence is inconsistent, which does not support a claim of benefit for bradycardia.

### 3.6 Respiratory adverse events (respiratory depression/hypoxaemia)

Five studies reported respiratory depression/hypoxaemia (Xu 2025, Zhang 2025 and Fan 2024 reported “respiratory depression”, while Bai 2025 and Liu 2026 reported hypoxaemia; these were harmonised as events related to a fall in SpO_2_ and then included). This outcome is a composite respiratory adverse-event outcome defined by the authors rather than a single endpoint identical across trials. Pooling with a random-effects model gave OR 0.19 (95% CI 0.11–0.32; *I*^2^ = 0%; Figure 4), a Peto OR of 0.22 (0.14–0.35), and RD *−*12.4% (*−*18.3% to *−*6.4%) with an NNT of about 8. Tang 2026 reported a composite of “respiratory depression/cough” (1/52 versus 0/29) that could not be separated and was not pooled. When analysed by the original definitions, the hypoxaemia studies (Bai 2025 and Liu 2026) gave a pooled OR of 0.20 (0.11–0.38) and the respiratory depression studies (Xu 2025, Zhang 2025 and Fan 2024) gave 0.15 (0.05–0.44); the directions agree, but each layer contains only two or three studies, so an influence of definition differences on the effect size cannot be excluded. Because mechanically ventilated ICU patients are already receiving ventilatory support, respiratory depression is difficult to observe in that setting, and this outcome comes mainly from endoscopy and hysteroscopy settings.

### 3.7 Hypertriglyceridaemia

Four studies reported hypertriglyceridaemia. Fospropofol disodium reduced the risk (OR 0.29, 95% CI 0.13–0.65; *I*^2^ = 0%; RD *−*8.3%, *−*16.4% to *−*0.2%; Supplementary Material S6), consistent with the absence of a lipid emulsion carrier. Because the upper limit of the 95% CI for RD is close to 0, the NNT of about 12 has an unstable interval and should not be used alone as a benefit measure. All four studies (Feng 2026, Gao 2025, Gao 2024 and Xiong 2026) were conducted in ICU settings with prolonged sedation (at least 24 h), and no short procedural study reported this outcome. This distribution has a biological rationale: plasma triglyceride levels change only after a lipid load lasting hours, and such change has little clinical meaning during endoscopy or hysteroscopy lasting a few tens of minutes. The conclusion for this outcome therefore applies to prolonged ICU sedation and not to short procedures.

### 3.8 Pruritus/paraesthesia

Five studies reported pruritus/paraesthesia; both arms of Gao 2024 had no events and the study was not pooled, so the pooled analysis is based on the remaining four studies. Pooling with a random-effects model showed a clear increase with fospropofol disodium (OR 32.56, 95% CI 12.89–82.26; *I*^2^ = 0%; Figure 5), with RD +28.5% (+11.2% to +45.7%) and an NNH of about 4. The direction was consistent across studies: Tang 2026 (20/52 versus 0/29), Zhan 2025 (51/90 versus 3/90), Liu 2026 (30/213 versus 1/213) and Fan 2024 (17/126 versus 0/34). The pooled value is sensitive to how zero cells are handled, and here the Peto estimate is clearly lower: Peto gave 8.71 (5.77–13.13), below the pooled value and below the corrected OR of every individual study (11.0 to 37.9), and below the crude pooled OR that requires no correction (29.4, based on 24.5% versus 1.1%). The reason is that Peto assumes an effect close to the null and similar arm sizes, whereas all study-specific ORs here exceed 11 and the arms in Fan 2024 (126 versus 34) and Tang 2026 (52 versus 29) are very unbalanced. We therefore report the random-effects estimate as the primary result, consistent with the registered protocol, and reports the Peto estimate alongside as a sensitivity analysis. Both methods agree in direction, but the magnitude is imprecise and is best interpreted together with the risk difference (+28.5%): the main cost of fospropofol disodium is transient, self-limiting pruritus or abnormal sensation after administration, with an absolute increase of about 28 percentage points.

**Figure 5:**
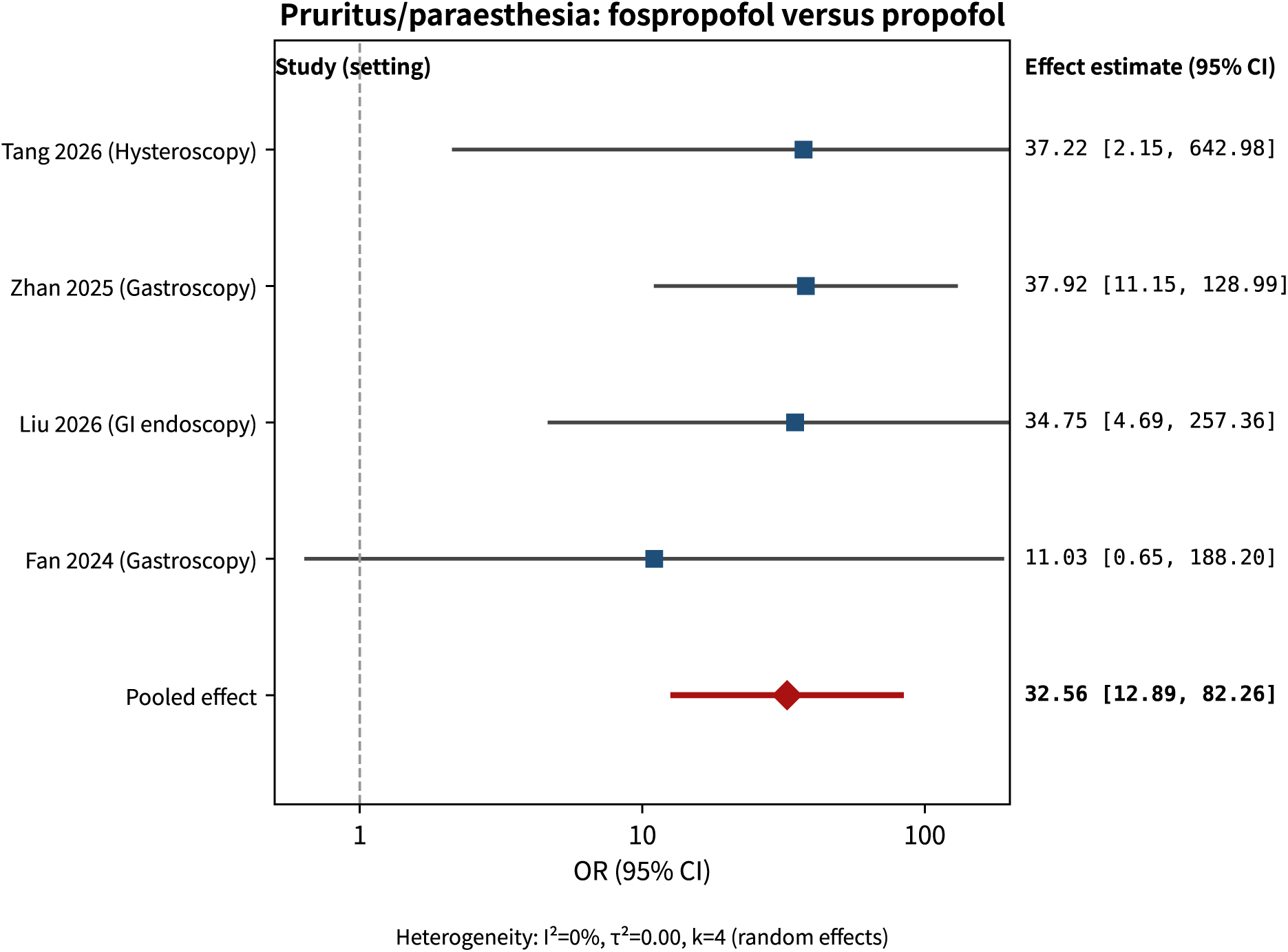
Forest plot for pruritus/paraesthesia (random-effects model, primary analysis). The area of each square does not represent weight.

### 3.9 Sedation efficacy, recovery and extubation

Sedation efficacy was not pooled quantitatively. Definitions and measurement approaches differed across studies (proportion of time at target, proportion attaining a RASS target, success rate of sedation and analgesia, and number of supplementary doses), so pooling would have required additional assumptions. The comparison is therefore narrative: Gao 2025 reported target attainment in 96.78% versus 98.43% of patients, Gao 2024 reported medians of 83.33% in both groups, and Xiong 2026 reported no difference in the success rate of sedation and analgesia. These data suggest similar sedation efficacy, but because no non-inferiority margin was set, no pooling was performed and no GRADE rating was applied, this review cannot conclude that the two drugs are equivalent. Successful extubation did not differ significantly (2 studies; OR 1.08, 95% CI 0.51–2.29; RD +1.7%, 95% CI *−*15.0% to +18.5%; *I*^2^ = 0%; Supplementary Material S6), but with only two studies and few events this cannot establish equivalence either.

#### Continuous outcomes

Three studies provided continuous data that could be pooled (Supplementary Material S6). Onset or loss-of-consciousness time was reported by only two studies with extreme heterogeneity (*I*^2^ = 99.6%) and was therefore not pooled; instead, Zhan 2025 reported 3.14 *±* 0.66 versus 0.92 *±* 0.20 min (MD +2.22 min, 95% CI +2.08 to +2.36) and Bai 2025 reported 1.46 *±* 0.23 versus 0.54 *±* 0.16 min (MD +0.92 min, 95% CI +0.85 to +0.98). Both show slower onset with fospropofol disodium, but the magnitudes differ by about 1.3 min. Recovery time (3 studies; MD *−*0.33 min, *−*1.46 to 0.80; *I*^2^ = 92%) and discharge or room-exit time (3 studies; MD *−*0.67 min, *−*1.59 to 0.25; *I*^2^ = 85%) did not differ significantly between groups, but heterogeneity was high and the confidence intervals were wide, so a non-significant result does not establish equivalence and the evidence is insufficient to demonstrate it. The onset evidence comes only from two procedural sedation studies that used intravenous bolus dosing (Zhan 2025 and Bai 2025) and reflects the time to loss of consciousness after a bolus. In goal-directed continuous ICU sedation the situation may be the opposite: Feng 2026 reported that the target attainment time was shorter with fospropofol disodium than with propofol (1.74 versus 2.59 min, reported *P* = 0.02; the reported dispersion and test statistic in that article are not fully self-consistent, so only the point estimate and the authors’ *P* value are reported). These two studies measured different processes (onset after a loading bolus versus titration to target during continuous infusion), so “slower onset” should not be regarded as a general feature of fospropofol disodium across all sedation settings. The continuous outcomes rest on few studies with high heterogeneity, and their estimates are indicative only.

### 3.10 Body movement and cough during the procedure

Three studies reported body movement during the procedure and two reported cough. Neither outcome was pooled quantitatively, because the direction of the effect differed between studies. For movement, Zhan 2025 reported substantially fewer events with fospropofol disodium (10/90 versus 56/90) and Tang 2026 also reported fewer (13/52 versus 13/29), whereas Liu 2026 reported more with fospropofol disodium (67/213 versus 26/213, counting any movement grade, that is, a movement score below 10); pooling the three gives OR 0.48 (0.04–5.44) with *I*^2^ = 97%, which cannot be interpreted. For cough, Zhan 2025 reported fewer events with fospropofol disodium (5/90 versus 15/90) whereas Liu 2026 reported more (2/213 versus 0/213). This inconsistency most likely reflects differences in threshold and assessment: Liu 2026 used a five-level movement score and counted occasional slight movement as an event, whereas Zhan 2025 and Tang 2026 counted clinically evident movement. We therefore list the raw counts only (Table 2) and does not pool them or claim a reduction in movement or cough; the heterogeneity also indicates that the claim of fewer procedural adverse events applies to injection pain and respiratory adverse events and should not be extended to movement.

### 3.11 Patient-important outcomes

Because most safety endpoints are surrogate outcomes, we also extracted patient-important outcomes. These outcomes were reported very infrequently (death and delirium in one study each; ICU length of stay and duration of mechanical ventilation in two studies each) and their definitions and measurement times differed between studies. They were therefore not pooled quantitatively, and this review draws no directional or equivalence conclusion from them; the available data are recorded item by item below:

- **Delirium:** Feng 2026 reported similar incidences of delirium in the fospropofol disodium and propofol groups (7/70 versus 9/70).
- **28-day all-cause mortality:** only Gao 2025 provided data (13/30 versus 12/30, *P >* 0.999). That study had only 60 participants and 25 deaths in total, so its power to test mortality is very low and it can support neither a non-inferiority nor a superiority conclusion.
- **ICU length of stay:** Feng 2026 reported 10.57 *±* 2.69 versus 10.69 *±* 2.79 days, and Gao 2025 reported medians of 5.50 (4.00–11.50) versus 10.00 (5.75–17.75) days, with no statistically significant difference.
- **Duration of mechanical ventilation or ventilator-free time:** Feng 2026 reported 8.6 *±* 2.09 versus 8.71 *±* 2.44 days, and Xiong 2026 reported 7-day invasive ventilator-free time of 61.49 *±* 5.35 versus 61.28 *±* 5.27 h; the groups were similar.
- **28-day non-ICU hospital stay (days):** Xiong 2026 reported 2.18 *±* 0.35 versus 2.07 *±* 0.32 days, with similar values in both groups.
- **Successful extubation:** no statistically significant difference, but the evidence is insufficient to establish equivalence (see above).

Overall, the 12 available studies cannot support any definitive conclusion on death, delirium or ventilator-free days: these outcomes were reported infrequently, and each was provided by only one or two small studies. The absence of patient-important outcomes is the main knowledge gap in this review and the priority for future research.

### 3.12 Sensitivity analyses

Leave-one-out analysis showed that the pooled OR for hypotension ranged from 0.44 to 0.58, always favouring fospropofol disodium; the confidence interval crossed 1 after excluding Feng 2026, Bai 2025, Zhang 2025, Xu 2025 or Liu 2026, and remained significant after excluding each of the other five studies (Supplementary Material S6). The overall estimate is therefore not driven by any single study, but its statistical significance depends on pooling all 10 studies.

1. **Exclusion of the preprint.** After excluding Liu 2026, the pooled OR for hypotension became 0.58 (0.31–1.11) with a confidence interval crossing 1, so the statistical significance of the overall hypotension estimate partly depends on this preprint. Within procedural sedation, however, the estimate remained OR 0.22 (0.09–0.52) with the direction and significance unchanged. The direction and significance of every other outcome were unchanged: injection pain (Peto) 0.11 (0.08– 0.17); bradycardia OR 0.29 (0.12–0.70); respiratory adverse events (Peto) 0.19 (0.10–0.36); and pruritus/paraesthesia (Peto) 9.34 (5.68–15.34).
2. **The two sensitivity analyses for injection pain differ and should be reported separately.** First, restricting to studies with observed events (excluding the three studies with zero events in the fospropofol disodium arm, *k* = 5) gave a pooled OR of 0.13 (0.05–0.35) with *I*^2^ = 74% and *τ* ^2^ = 0.97. Second, excluding the preprint (*k* = 7) gave a pooled OR of 0.09 (0.03–0.26) with *I*^2^ = 69% and *τ* ^2^ = 1.28. The two analyses use different study sets, estimators and results. The results for injection pain under the four analysis settings are listed side by side in the summary table in Appendix 6 (including the primary analysis and each exclusion scheme) so that they can be checked line by line; no two rows should be treated as interchangeable.
3. **Handling of dose arms in multi-arm trials.** Tang 2026 was a three-arm trial (fospropofol disodium 12.5 mg/kg, *n* = 28; 15 mg/kg, *n* = 24; propofol, *n* = 29), and the primary analysis combined the two dose arms. Including only the 12.5 mg/kg or only the 15 mg/kg arm gave essentially unchanged results (hypotension OR 0.39 [0.22–0.67] and 0.37 [0.22–0.63]; injection pain Peto OR 0.13 and 0.13; pruritus/paraesthesia Peto OR 9.18 and 9.24). Fan 2024 was a five-arm trial (fospropofol disodium 8.0, 10.0, 12.5 and 15.6 mg/kg versus propofol 2.0 mg/kg, *n* = 34), and the primary analysis combined the four dose arms (*n* = 126); restricting to therapeutic dose arms (excluding the 8.0 mg/kg arm, which the authors judged to be below the appropriate dose) gave similarly stable results (injection pain Peto OR 0.13 [0.09–0.18]; pruritus/paraesthesia 8.83 [5.85–13.33]; respiratory depression 0.23 [0.15–0.35]). The handling adopted here should be stated explicitly: the primary analysis includes every randomised dose arm so as to remain consistent with the original article and with randomisation, rather than excluding an already randomised arm after the fact; however, *clinical interpretation should rest on the therapeutic-dose arms*, since 8.0 mg/kg was judged by the authors themselves to be below the appropriate dose and is not a recommended regimen. Including it does not affect the pooled results (the two conventions give almost identical values for every outcome; see Supplementary Material S7).
4. **Robustness to the preprint and to overlapping cohorts (exploratory analyses).** The hypotension conclusion partly depends on the preprint (Liu 2026) and on two cohorts that may overlap (Gao 2024 and Gao 2025).

- *Perturbation of the preprint event counts:* after reverse-direction perturbation of the two arms of Liu 2026, the pooled OR for hypotension remained between 0.51 and 0.59, with confidence intervals that did not cross 1 in any perturbation scenario; complete exclusion of the study gave 0.58 (0.31–1.11) with a confidence interval crossing 1. Results are given in Appendix 6.
- *Overlapping cohorts:* Gao 2024 and Gao 2025 came from the same team and centre (Department of Critical Care Medicine, Union Hospital, Wuhan). The author lists are essentially identical and both had 60 participants (30 versus 30), but the sedation depth targets differed (Gao 2024, light-to-moderate sedation at RASS *−*3 to 0; Gao 2025, deep sedation at RASS *−*4 to *−*5), Gao 2025 was registered (NCT05870514) and cites Gao 2024 as that team’s earlier work. We therefore judged them to be separate, sequentially conducted studies rather than duplicate publication of one cohort, although partial patient overlap cannot be fully excluded without individual-level information. Sensitivity analysis showed that for hypertriglyceridaemia, excluding Gao 2025 gave OR 0.26 (0.08–0.81), excluding Gao 2024 gave 0.31 (0.12–0.82), and excluding both gave 0.27 (0.04–1.67); for hypotension, excluding Gao 2025 gave OR 0.46 (0.24–0.88) and excluding Gao 2024 gave OR 0.44 (0.24–0.80). The conclusions for hypertriglyceridaemia and hypotension did not change when either study was removed.
5. **Stratification by blinding (exploratory analysis).** This analysis directly tests the most fundamental threat to validity in this review. Only Liu 2026 and Fan 2024 used a double-blind design among the 12 pooled studies (Table 4). For injection pain the direction was opposite to the expectation that open-label studies overestimate benefit (double-blind, 2 studies, OR 0.04, 0.02–0.10; other studies, 6 studies, OR 0.13, 0.05–0.36), but the difference was not statistically significant (*Q_bet_* = 3.05, *df* = 1, *P* = 0.081); this test has limited power with *k* = 2 and *k* = 6 strata and can only be regarded as descriptive. Respiratory adverse events (double-blind 0.19 versus other 0.18, *P* = 0.910) and pruritus/paraesthesia (23.73 versus 37.81, *P* = 0.645) pointed in the same direction in both strata, and the differences were not significant. For hypotension and bradycardia the blinded stratum contained only Liu 2026, and the interaction test is not valid when a stratum contains one study, so these are described only: in the single double-blind study, the direction for bradycardia reversed (8/213 versus 5/213, OR 1.62, 0.52–5.05), whereas the pooled OR for the other eight, non-double-blind studies was 0.29 (0.12–0.70), with a confidence interval that excludes the null. In other words, almost all of the apparent bradycardia benefit comes from non-double-blind studies, while the only double-blind study points the other way. The interaction test remains invalid because one stratum contains a single study, so this cannot be attributed to open-label bias; it does show, however, that the evidence for a bradycardia benefit is unstable and should not be presented as an advantage of fospropofol disodium.
6. **Sensitivity analysis of data-extraction conventions.** The impact of the four extraction conventions used in this review (treating infusion-site pain in Candiotti 2011 as injection pain; the possible cohort overlap between Gao 2024 and Gao 2025; and pooling the dose arms of Fan 2024 and of Tang 2026) is quantified in Supplementary Material S7. The main points are as follows. Injection pain was OR 0.07 (0.03–0.19) when Candiotti 2011 was excluded, versus 0.09 in the primary analysis. Retaining only the therapeutic dose arms of Fan 2024 gave 0.09 for injection pain, 0.19 for respiratory adverse events and 32.87 for pruritus/paraesthesia, versus 0.09, 0.19 and 32.56 in the primary analysis. Analysing the arms of Tang 2026 separately gave 0.49–0.51 for hypotension, 0.09 for injection pain and 32.45–32.68 for pruritus/paraesthesia. Excluding either or both of the Gao studies gave 0.37–0.46 for hypotension, 0.36–0.45 for bradycardia and 0.26–0.31 for hypertriglyceridaemia. None of the four conventions changed the direction or statistical significance of any outcome. The one result that warrants attention is hypertriglyceridaemia, whose confidence interval crosses the null when both Gao studies are excluded (0.04–1.67); this is consistent with its very low certainty and its status as a hypothesis-generating finding only, and confirms that it cannot support a definitive conclusion.
7. **Estimator sensitivity analysis (exploratory analysis).** To examine how the pooling method affects the conclusions, every outcome was recalculated with four methods (DerSimonian-Laird as the primary analysis, REML plus modified Hartung-Knapp, Mantel-Haenszel and Peto, with 95% prediction intervals), and the results are given in Appendix 6. Apart from hypertriglyceridaemia, the direction and significance of every outcome were consistent across the four methods. Hypertriglyceridaemia was the only outcome whose conclusion changed: DerSimonian-Laird gave OR 0.29 (0.13–0.65) whereas REML plus modified Hartung-Knapp gave 0.29 (0.08–1.08) with a confidence interval crossing 1. *τ* ^2^ was 0 under both DerSimonian-Laird and REML, so the two methods give the same point estimate; the interval differs because with *k* = 4 the modified Hartung-Knapp method uses the *t*_(3)_ quantile and truncates the standard error at the fixed-effect level, which widens the interval relative to DerSimonian-Laird. The conclusion for this outcome is therefore sensitive to the interval estimation method rather than to the point estimate or heterogeneity, and it is not treated as a definitive finding (GRADE certainty is very low). In addition, the 95% prediction intervals for hypotension and injection pain crossed 1 (0.09–2.72 and 0.01–1.25 respectively), indicating that extrapolation across settings warrants caution.

**Table 4:** Subgroup analysis by blinding status (exploratory analysis).

| Outcome | Stratum | Studies, n | Pooled OR (95% CI) | Interaction P |
| --- | --- | --- | --- | --- |
| Hypotension | Double-blind | 1 | 0.25 (0.12, 0.54) | Not testable<br>(single study) |
|  | Not double-blind or not described | 9 | 0.58 (0.31, 1.11) |  |
| Injection pain | Double-blind | 2 | 0.04 (0.02, 0.10) | 0.081 |
|  | Not double-blind or not described | 6 | 0.13 (0.05, 0.36) |  |
| Bradycardia | Double-blind | 1 | 1.62 (0.52, 5.05) | Not testable<br>(single study) |
|  | Not double-blind or not described | 8 | 0.29 (0.12, 0.70) |  |
| Respiratory depression/hypoxaemia | Double-blind | 2 | 0.19 (0.10, 0.38) | 0.910 |
|  | Not double-blind or not described | 3 | 0.18 (0.08, 0.42) |  |
| Pruritus/paraesthesia | Double-blind | 2 | 23.73 (4.62, 121.82) | 0.645 |
|  | Not double-blind or not described | 2 | 37.81 (12.28, 116.44) |  |

Note: The interaction test uses the between-group heterogeneity $Q_{bet}$ with the same model as the primary analysis. "Not testable" means that a stratum contains a single study, so the within-stratum "pooled" estimate is that study itself and the interaction test is not methodologically valid.

### 3.13 Trial sequential analysis

The exploratory trial sequential analysis (TSA) for hypotension, including the heterogeneity adjustment and the per-setting analyses, is reported in Supplementary Material S5. In the pooled analysis the cumulative information reached only 26% of the heterogeneity-adjusted requirement and did not cross the monitoring boundary; in the procedural sedation stratum (*I*^2^ = 0%) the cumulative Z value (*−*4.93) did cross the stratum boundary (3.37), whereas the ICU stratum (*Z* = *−*0.37) was far from crossing. The per-setting TSAs were not adjusted for multiplicity across strata; even if the two-sided *α* is split by Bonferroni (*α* = 0.025) or into thirds (*α* = 0.0167), the procedural-stratum boundaries are 3.85 and 4.11 respectively, both below the cumulative Z value of *−*4.93, so the result is robust to *α* adjustment. The result also depends on the pre-specified relative risk reduction (RRR): it still crosses when the RRR is reduced from 25% to 18% (boundary 4.76) but no longer crosses at 16% or below; together with the fact that the analysis does not account for the *α* spent on the interaction test, this means the finding is supportive only (sensitivity analysis in Table S5-3). Because this outcome shows a significant interaction between settings (*P <* 0.001), a single TSA of the pooled effect has limited value, so the analysis is reported as exploratory only and is not used to support or refute the main conclusions; its inputs, boundaries and limitations are given in Supplementary Material S5.

### 3.14 Publication bias

Hypotension included 10 studies, which meets the threshold of at least 10 studies specified in the registered protocol, so the funnel plot and the Harbord test were performed (using Harbord as the primary test is a deviation from the protocol, listed in section 2.1): the funnel plot showed no obvious asymmetry and the Harbord test gave an intercept of 1.55 (*t* = 1.19, *df* = 8, *P* = 0.27), detecting no small-study effect. For comparability with earlier literature and with the original protocol, Egger’s test (intercept 0.43, *P* = 0.76) is also reported; both tests and their implementation are given in Supplementary Material S6-E. Three caveats apply. First, *k* = 10 only just reaches the conventional minimum and the power of these tests remains low. Second, *I*^2^ = 51% for this outcome indicates substantial heterogeneity, whereas all funnel-plot tests assume that studies share a single true effect, and heterogeneity alone can produce asymmetry. Third, these tests detect *small-study effects*, which may reflect publication bias, differences in populations, differences in methodological quality, or other causes. The result therefore neither supports nor excludes publication bias and must not be reported as excluding it; it is presented for reference only. Injection pain reached eight studies, but three of them had zero events in the fospropofol disodium arm, and funnel plots are unreliable with such data, so no publication bias test was performed for that outcome.

### 3.15 Subgroup and cumulative analyses

By setting, the effect of fospropofol disodium on hypotension was clearly larger during procedural sedation (OR 0.24, 0.13–0.42; *I*^2^ = 0%) than during ICU sedation (OR 0.90, 0.53–1.55; *I*^2^ = 2%), with an interaction *P <* 0.001 (Figure 3). All injection pain data came from procedural sedation, so a between-setting comparison was not possible. For bradycardia, the setting stratification showed no significant interaction (ICU, 5 studies, OR 0.68, 0.21–2.19; procedural sedation, 4 studies, OR 0.28, 0.05–1.64; *P* = 0.412). In a cumulative meta-analysis ordered by study sample size from smallest to largest (the horizontal axis is cumulative sample size), the cumulative confidence interval crossed 1 for the first nine studies, and the cumulative OR first excluded 1 (0.51, 0.28–0.93) only when the final and largest study, Liu 2026 (*n* = 426), was added (Supplementary Material S6). That order is determined by sample size and was not chosen by effect size, but pooled estimates in a cumulative analysis nevertheless depend on the order of inclusion, and the order differs from the chronological ordering used in the trial sequential analysis, so this is presented as a trend only.

### 3.16 Benefit and harm reported side by side

Expressed as risk differences, the NNT/NNH values were: injection pain NNT about 4, respiratory depression about 8, hypotension about 14 and hypertriglyceridaemia about 12; pruritus/paraesthesia NNH about 4 (Supplementary Material S6). These events are not equivalent in clinical severity, duration or the need for intervention, so the numerical values of NNT and NNH alone cannot be used to derive net clinical benefit or to rank the drugs. We report them side by side only: fospropofol disodium may reduce injection pain and respiratory events while clearly increasing pruritus or abnormal sensation; the final benefit-harm judgement should take account of the specific setting, event severity and patient preferences.

### 3.17 Certainty of evidence

Certainty of evidence for each outcome is shown in Table 5 and Supplementary Material S6: low for hypotension, injection pain, respiratory depression/hypoxaemia, pruritus/paraesthesia and successful extubation, moderate for bradycardia, and very low for hypertriglyceridaemia. Downgrading was judged separately for each outcome. Inconsistency was applied to hypotension (setting interaction *P <* 0.001), injection pain (*I*^2^ = 65%); bradycardia was not downgraded for inconsistency (*I*^2^ = 44%, with 8 of 9 studies pointing in the same direction and the confidence interval of the only double-blind study including the null). Imprecision was applied to hypotension (the REML plus modified Hartung-Knapp interval 0.25–1.03 crosses 1 and the 95% prediction interval is 0.09–2.72), bradycardia (wide CI crossing the null), hypertriglyceridaemia (only 4 studies with few events and an interval width that depends on the interval estimation method), pruritus/paraesthesia (only 4 studies, with the OR between 8.7 and 46.6 depending on how zero cells are handled) and successful extubation (only 2 studies). Indirectness was applied to respiratory depression/hypoxaemia (observable only during procedural settings) and to hypertriglyceridaemia (prolonged ICU sedation only, with inconsistent thresholds of 1.7–2.3 mmol/L and measurement times). Risk of bias was applied to subjective outcomes such as injection pain and pruritus/paraesthesia, to the limitations in randomisation and data completeness in the respiratory depression studies, and to hypertriglyceridaemia, in which all 4 studies were open-label and Gao 2024 and Gao 2025 came from the same centre and may overlap. Hypotension, bradycardia, respiratory depression/hypoxaemia and successful extubation are objectively measured and were not downgraded for the open-label design alone.

**Table 5:**
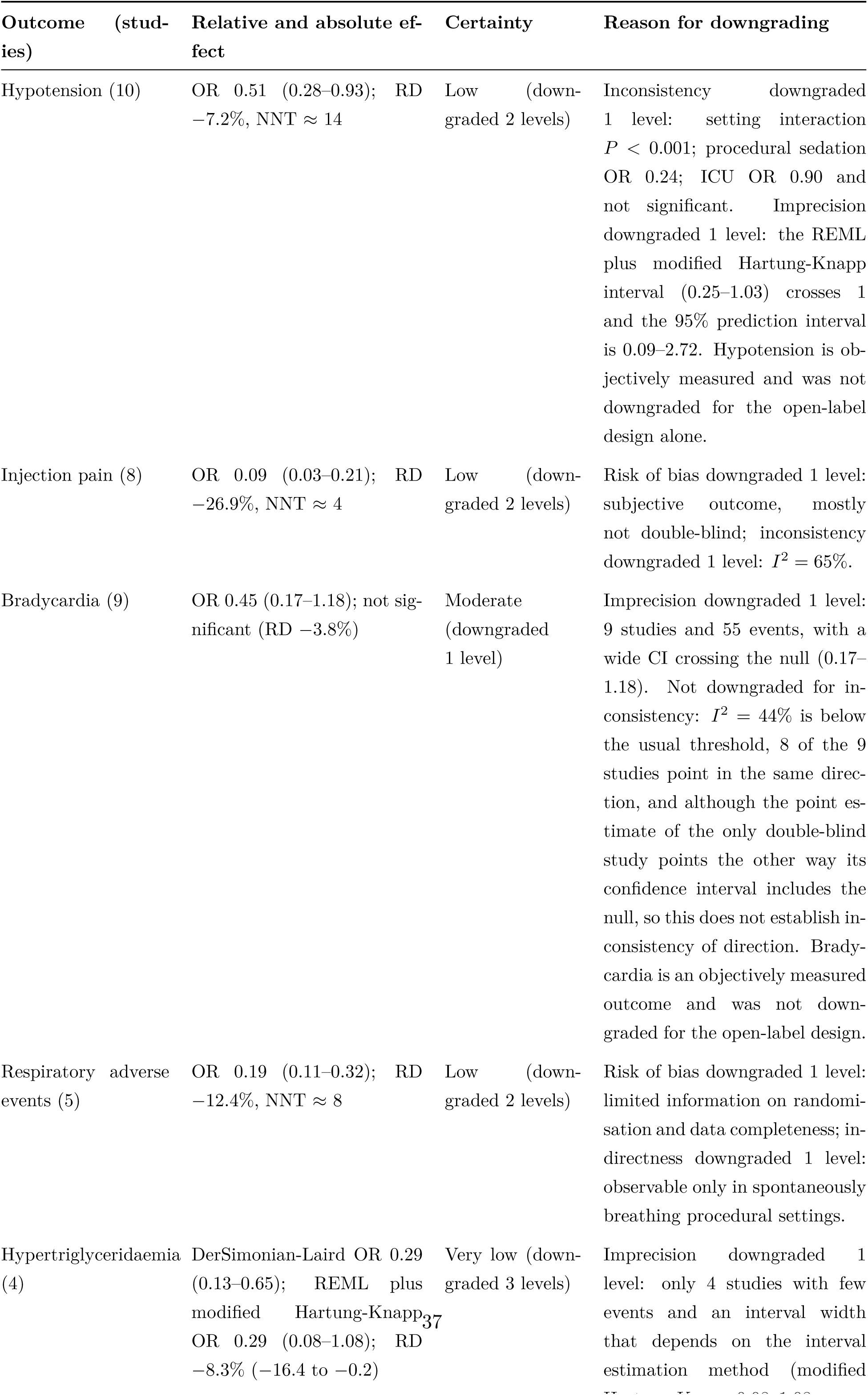
GRADE summary of findings for the main outcomes.

Certainty of evidence and robustness of the conclusion answer different questions. Low certainty mainly reflects the small number of original studies, their small samples and limited methodological reporting. Most sensitivity analyses supported the direction of the effects on injection pain, respiratory adverse events and pruritus/paraesthesia; the point estimate for hypotension was consistent in direction across analyses, but its statistical significance was sensitive to the preprint and to the setting composition. Hypertriglyceridaemia is a clear exception: the confidence interval crossed the null under the modified Hartung-Knapp method and after excluding both Gao 2024 and Gao 2025, and the hypotension TSA no longer crossed its boundary after excluding Liu 2026. We therefore do not describe all outcomes as robust and explicitly downgrades hypertriglyceridaemia and the TSA conclusion to uncertain or exploratory.

## 4 Discussion

### 4.1 Main findings

Within the literature retrieved here, this is, to our knowledge, the first published systematic review that specifically synthesises head-to-head randomised evidence on fospropofol disodium versus propofol for sedation outside the operating room (ICU and procedural sedation), and it does not overlap with the published induction meta-analysis. Pooled results from 12 RCTs showed that injection pain and respiratory adverse events favoured fospropofol disodium (OR 0.09 and 0.19). For hypertriglyceridaemia the direction was the same under the DerSimonian-Laird model (OR 0.29), but the REML plus modified Hartung-Knapp confidence interval crossed 1 and GRADE certainty was very low, so this is treated as a hypothesis only. The effect on hypotension was concentrated in procedural sedation (OR 0.24) and was absent during continuous ICU sedation (OR 0.90), with an interaction *P <* 0.001; the overall pooled estimate (OR 0.51) crossed 1 under REML plus modified Hartung-Knapp and is therefore not used as a basis for conclusions. Bradycardia did not differ significantly. Successful extubation did not differ significantly, but the evidence is insufficient to conclude equivalence. Onset was slower with fospropofol disodium during bolus-dosed procedural sedation, and pruritus and abnormal sensations increased clearly (OR 32.56; RD +28.5%). Overall certainty ranged from very low to moderate.

### 4.2 Consistency with previous syntheses and with mechanism

These results are broadly consistent with the pharmacology of fospropofol disodium. The drug is water-soluble and is slowly released by alkaline phosphatase, so plasma concentrations rise gradually, injection pain and circulatory and respiratory depression are reduced, and the lipid load from a lipid emulsion is avoided. The findings of the previous induction meta-analysis (less injection pain, slower onset, more pruritus and numbness) are reproduced here. Its suggestion of less bradycardia was not reproduced: the effect was not significant under random effects, and the direction reversed in the only double-blind study (8/213 versus 5/213, OR 1.62). This indicates that the bradycardia outcome is sensitive to blinding and to the choice of statistical method, and it should not be presented as an advantage of fospropofol disodium.

### 4.3 Clinical implications

The populations in which these findings might apply are inferred from pharmacology. We did not include trials that specifically enrolled older patients, patients with cardiorespiratory impairment, patients with dyslipidaemia or patients who cannot tolerate lipid emulsion, so applicability to those groups still requires dedicated studies. During procedural sedation, less injection pain and fewer respiratory events, together with a clear reduction in hypotension, may improve patient experience and procedural safety. The slower onset should be taken into account, and pre-dosing or supplementary doses may be needed. Pruritus and abnormal sensations are common but are usually transient and resolve without intervention.

### 4.4 Setting heterogeneity and the limits of extrapolation

The most important heterogeneity observed here relates to setting: the benefit for hypotension was clear during procedural sedation (OR 0.24) and absent during continuous ICU sedation (OR 0.90), and the interaction test supports a difference between the two (*P <* 0.001). This difference has a clinical explanation. ICU patients often have septic shock, receive vasoactive drugs, and are sedated more deeply and for longer, so a difference between two drugs in haemodynamic effect is harder to detect against that background. Procedural sedation is shorter and baseline haemodynamics are relatively stable, so the smoother plasma concentration profile produced by a prodrug is more likely to translate into an observable reduction in hypotension. Pooling the two settings would overestimate the ICU benefit and underestimate the benefit during procedures. We therefore distinguish the two settings in its conclusions and does not claim consistency across settings.

### 4.5 Potential costs: delayed onset and pruritus/paraesthesia

Two main disadvantages of fospropofol disodium need to be weighed in clinical decisions. First, as a prodrug it requires hydrolysis by alkaline phosphatase, so onset or loss of consciousness after an intravenous bolus during procedural sedation is slower: Zhan 2025 and Bai 2025 reported MD +2.22 min and +0.92 min respectively, and because the two differ widely no pooling was performed. In outpatient procedures that require rapid onset or fast turnover, pre-dosing or supplementary doses may be needed, whereas no such disadvantage was observed during titrated continuous infusion in the ICU (target attainment was in fact faster with fospropofol disodium in Feng 2026). Recovery and discharge times did not differ significantly, but heterogeneity was high, so equivalence to propofol cannot be concluded. Second, pruritus and abnormal sensations are the most common characteristic adverse effects after administration, and their mechanism is not yet clear; current hypotheses involve stimulation of sensory nerve endings by the phosphate group in the molecule and by the metabolite formate [21]. These reactions may be reduced by diluting the drug or slowing the injection, but the reported rate was clearly higher in the included studies (OR 32.56; RD +28.5%). We did not extract data on their duration, severity or need for intervention, so any statement that they are transient or self-limiting remains a mechanistic inference rather than a finding of this review. Some early studies used fentanyl pretreatment to reduce the reaction, but a recent randomised trial showed that premixed lidocaine did not effectively relieve fospropofol-related paraesthesia [22], so preventive measures remain debated and warrant prospective evaluation.

### 4.6 Metabolic safety of formaldehyde and formate

When alkaline phosphatase hydrolyses fospropofol disodium to release propofol, formaldehyde and phosphate are also produced, and formaldehyde is rapidly metabolised to formate by formaldehyde dehydrogenase [17]. In vitro and clinical pharmacology studies indicate that plasma formate concentrations after fospropofol disodium are within the endogenous range and well below the toxic concentrations seen in methanol poisoning, and no case of formate toxicity attributable to this drug has been reported [17]. In theory, however, prolonged high-dose infusion may still carry a risk of formate accumulation, and the withdrawal of the drug from the US market in 2012 has been associated with concerns about formaldehyde accumulation and safety in the outpatient setting [21]. Among the included studies, Gao 2024 reported no significant difference in plasma formate concentrations between groups at any time point, while the others did not systematically report blood gas variables (pH, bicarbonate, lactate) or formate concentrations. Evidence on formate safety during prolonged high-dose use, particularly continuous ICU sedation, therefore remains insufficient and remains a major limitation of this review and a priority for future research.

### 4.7 Relationship to newer intravenous sedatives

Other intravenous sedatives commonly used outside the operating room include remimazolam, dexmedetomidine, ciprofol and etomidate. We did not construct a network meta-analysis (NMA) including these drugs. First, direct comparative evidence is sparse and the network lacks reliable connecting edges. Within the literature retrieved here, no head-to-head randomised trial of fospropofol disodium against remimazolam, dexmedetomidine, ciprofol or etomidate was identified. Because the search strategy required propofol to be present, so trials using only midazolam or another non-propofol drug as the comparator cannot be identified completely; Feng 2026, for example, includes a midazolam arm, which shows that such studies exist. We can therefore state only that the present search did not identify sufficient direct evidence to build a network, and cannot claim that such head-to-head trials do not exist. Second, transitivity would be difficult to establish even if a network were built. The present data show a significant interaction between settings (the interaction for hypotension, *P <* 0.001), which means that even within the single comparison of fospropofol disodium versus propofol, ICU and procedural settings cannot be treated as interchangeable; adding differences in concomitant drugs (fentanyl, alfentanil, sufentanil, remifentanil), sedation depth targets (RASS and MOAA/S systems differ) and outcome definitions makes transitivity across comparisons even harder to justify. Building separate networks for each setting is feasible in principle, but at this level of sparsity it would produce indirect estimates with very low power. Third, forcing the data into one network would contradict the central finding of this review, namely that the settings differ fundamentally while assuming they are exchangeable. Several network meta-analyses have evaluated drug choice in similar settings, such as a network meta-analysis of 60 RCTs, 7071 patients and 32 regimens comparing propofol plus opioid with other strategies for gastrointestinal endoscopy sedation [24], and a Bayesian network meta-analysis of bronchoscopy sedation. None of these networks included fospropofol disodium. We therefore limit the scope to the direct comparison question of replacing propofol, which complements rather than duplicates those network analyses.

### 4.8 Implications for guidelines and practice

Current ICU sedation guidelines recommend propofol or dexmedetomidine rather than benzodiazepines for sedation of mechanically ventilated adults [6, 7], but none of them addresses differences between propofol formulations. The available evidence suggests that during procedural sedation fospropofol disodium may reduce injection pain, hypotension and respiratory adverse events, whereas during continuous ICU sedation the evidence for a hypotension benefit is insufficient and the evidence on bradycardia is inconsistent, so it does not justify changing first-line practice. Applicability to patients with dyslipidaemia or intolerance of lipid emulsion is an inference from mechanism: this review did not specifically include such patients, and a triglyceride surrogate endpoint should not be equated with a clinical benefit. Because the approved indication for fospropofol disodium in China is adult general anaesthesia induction only, every sedation use discussed here should be regarded as investigational and does not constitute a routine clinical recommendation.

### 4.9 Strengths

This synthesis brings together the sedation evidence for fospropofol disodium; within the literature retrieved here, no previous systematic review has covered this scope. Both Chineseand Englishlanguage literature were included and analysed by ICU and procedural setting. Heterogeneity, leave-one-out analysis, exclusion of the preprint and trial sequential analysis are all reported, and certainty of evidence was assessed with GRADE. All outcomes were pooled with a random-effects model as specified in the registered protocol, Peto estimates and risk differences are reported alongside for outcomes with a high proportion of zero cells, and the influence of zero-cell handling is stated explicitly. We also stratified by blinding status and performed sensitivity analyses for a potentially overlapping cohort from one team and for the preprint event counts. Most sensitivity analyses did not change the direction of the effects on hypotension, injection pain, respiratory depression or pruritus/paraesthesia, but hypertriglyceridaemia was sensitive to the estimator and to cohort exclusion, and the hypotension TSA depended on the preprint; not every conclusion can therefore be described as robust.

### 4.10 Limitations

Several limitations should be noted, and they fall into three areas.

#### Evidence base and applicability

One included study (Liu 2026) is a preprint that has not been peer reviewed; its hypotension and hypoxaemia event counts appear directly in the preprint tables and both the perturbation and exclusion analyses showed that the conclusions were stable, but it remains non-peer-reviewed evidence. Most outcomes are surrogates, and patient-important outcomes (death, delirium, ICU length of stay) were reported very infrequently, with each outcome provided by only one or two small studies, which cannot support any definitive conclusion. Eleven of the 12 studies came from China and only 2 reported double blinding; injection pain and pruritus/paraesthesia are subjective outcomes that are readily influenced by patient reporting and caregiver behaviour, so the generalisability of these effect sizes to non-Chinese practice settings remains uncertain. Gao 2024 and Gao 2025 came from the same team and centre, and although the sedation depth targets and registration information differ, partial patient overlap cannot be excluded completely. Search limitations include the following: Embase, Cochrane CENTRAL and Web of Science were searched on 21 September 2026 and screened record by record, and identified no additional eligible published RCT; however, because many registered trials have no results yet, this review cannot claim that unpublished literature has been exhausted. Future updates should continue to track registered trials that have been completed but not published and should re-check VIP coverage.

## Methods

The analysis is based on aggregate data and did not perform an individual participant data (IPD) meta-analysis, so individual-level subgroup analysis was not possible. Only 2 of the 12 pooled studies used double blinding, whereas the key outcomes are subjective or readily influenced by caregiving behaviour, so open-label bias is the most fundamental threat to validity. After stratification by blinding, the benefit of fospropofol disodium for injection pain was larger in the double-blind studies and the directions for respiratory depression and pruritus/paraesthesia were consistent, but the interaction tests for all three were not significant (*P* = 0.081, 0.910 and 0.645 respectively) and each stratum contained few studies with limited power, so these can only be regarded as descriptive. The blinded strata for hypotension and bradycardia each contained a single study, so bias could not be tested. In addition, the TSA covered hypotension only, and its *α* spending was not taken into account in the GRADE downgrade; onset time was not pooled, and the pooled recovery and discharge estimates rest on only three studies each with *I*^2^ *≥* 85%, so they are indicative only. In addition, although the pooled hypotension definition was standardised as “any event meeting the original threshold”, four of the 10 studies (Candiotti 2011, Xiong 2026, Zhang 2025 and Tang 2026) did not report an operational threshold, so their event counts had to follow the adverse-event counts reported in the original article. For Candiotti 2011 specifically, this review used the TEAE count (6/38 vs 2/22); the study-specific OR under that definition (1.88) is higher than the OR based on its separately reported serious adverse events requiring medical intervention (1/38 vs 1/22, OR 0.57). The standardised definition therefore sacrifices the estimate favourable to fospropofol disodium for this study rather than selecting an estimate by the direction of the result; nevertheless, the absence of thresholds still limits definitional consistency for this outcome.

### Extrapolation

We used a broad operational definition of sedation outside the operating room that includes ICU sedation, which does not correspond exactly to the narrow NORA concept; given the significant interaction between settings, the pooled estimates themselves should be interpreted with caution. The conclusion for hypertriglyceridaemia rests on four ICU studies and does not apply to short procedures.

#### 4.11 Implications for future research

The main implication for future work is a clearer specification of the trials that are now needed. Future studies should use patient-important outcomes as the primary endpoint, apply standardised sedation assessment (RASS; MOAA/S, the modified observer’s assessment of alertness/sedation) and lipid monitoring (triglycerides, formate), and prospectively evaluate whether opioid or midazolam pretreatment relieves pruritus/paraesthesia; multicentre collaboration should be encouraged to raise the level of evidence. Methodologically, the definitions and measurement times for hypertriglyceridaemia and bradycardia should be harmonised. The evidence on bradycardia is driven by a single study and only one blinded study is available, so future trials should prioritise adequate blinding and high-quality outcome measurement. Once sufficient head-to-head trials have accumulated between fospropofol disodium and newer intravenous sedatives such as remimazolam, dexmedetomidine, ciprofol and etomidate, a multi-node network meta-analysis could compare these drugs with propofol for efficacy and safety within one evidence network and inform drug selection across clinical settings.

## 5 Conclusions

During procedural sedation, fospropofol disodium may reduce injection pain and respiratory adverse events and may reduce hypotension compared with propofol; during continuous ICU sedation there was no difference in hypotension (OR 0.90) and no data were available for injection pain or respiratory adverse events, so the safety conclusions for ICU sedation remain limited. The overall pooled estimate (OR 0.51) crossed 1 under REML plus modified Hartung-Knapp and is not used as a basis for conclusions. Hypertriglyceridaemia was reduced only under the DerSimonian-Laird model, the REML plus modified Hartung-Knapp confidence interval crossed 1 and GRADE certainty was very low, so this cannot be treated as a definitive finding. Bradycardia did not differ significantly, and the direction reversed in the only double-blind study. After stratification by blinding, the injection pain benefit was larger in the double-blind studies and the directions for respiratory depression and pruritus/paraesthesia were consistent between strata, but the interaction tests were not significant (*P* = 0.081, 0.910 and 0.645) and statistical power was limited. Sedation efficacy was not pooled and successful extubation did not differ significantly, so equivalence between the two drugs cannot be asserted. The costs are slower onset after bolus dosing and more pruritus or abnormal sensation (about 1 in 4 patients), whereas no delayed onset was observed during continuous ICU infusion. Because certainty of evidence ranges from very low to moderate and the sedation use is off-label and investigational in China, the available evidence is insufficient for a routine clinical recommendation and supports further evaluation in randomised trials.

## Supporting information

Attachments to reproduce calculated data in this review

## Declarations

### Author contributions (CRediT)

Guiping Xu: clinical interpretation, supervision, guarantor; Yutong Shi: methodology, searching, screening, extraction, analysis, writing; Yang Wang: conceptualisation, screening, extraction, risk-of-bias assessment; Alimujiang Simayi and Li Qu: clinical interpretation, revision.

### Data availability

The analysis scripts (`analyze_en.py, analyze.py, analyze_v2.py, estimator_check.py`), the extracted data (JSON and CSV files in the data/ directory), the cell-by-cell source-of-truth audit tables (`source_of_truth_audit.csv/json`), the study-level RoB table, the outcome-specific RoB 2 tables for the four key outcomes (Supplementary Table S4), the two-reviewer screening table and the search records are provided with the manuscript as supplementary material; the protocol is available in PROSPERO (CRD420261507586).

## Appendix 1. Full search strategies

1. Europe PMC (REST API, default full-text fields, no field restriction): `fospropofol AND propofol AND (sedation OR endoscopy OR colonoscopy OR gastroscopy OR bronchoscopy OR hysteroscopy OR “intensive care” OR “mechanical ventilation” OR “critically ill” OR ICU)` 144 records (searched 21 September 2026).
2. PubMed (NCBI E-utilities, esearch): (fospropofol[tiab] OR “fospropofol disodium”[tiab] OR Lusedra[tiab] OR Aquavan[tiab] OR phosphopropofol[tiab]) AND (propofol[tiab] OR “2,6-diisopropylphenol”[tiab]) AND (sedation[tiab] OR sedative[tiab] OR endoscopy[tiab] OR colonoscopy[tiab] OR gastroscopy[tiab OR bronchoscopy[tiab] OR hysteroscopy[tiab] OR “intensive care”[tiab] OR “mechanical ventilation”[tiab] OR “critically ill”[tiab] OR ICU[tiab]) 55 records (searched 21 September 2026).
3. CNKI (professional search): `SU=(’fospropofol’+’fospropofol sodium’) AND SU=(’propofol’) AND SU=(’sedation’+’endoscopy’+’g ventilation’+’procedural sedation’)` 27 records (searched 21 September 2026).
4. Wanfang Data (advanced search): Subject:(“fospropofol” OR “fospropofol sodium”) AND Subject:(“propofol”) AND Subject:(“sedation” OR “endoscopy” OR “gastroscopy” OR “hysteroscopy” OR “bronchoscopy” OR “ICU” OR “mechanical ventilation” OR “procedural sedation”) 43 records (searched 21 September 2026).
5. VIP (Jinglun platform, accessed through the XJMU Library database proxy). The platform does not provide a shareable search URL; the following rule was submitted through its search interface ([+] indicates OR, [*] indicates AND, and TKS is the title, keyword and abstract subject field): (TKS=@k0[+]TKS=@k1)[*]TKS=@k2[*](TKS=@k3[+]TKS=@k4[+]TKS=@k5[+]TKS=@k6[+]TKS=@k7[+]TKS=@k8[+] Parameters: @k0 fospropofol, @k1 fospropofol sodium, @k2 propofol, @k3 sedation, @k4 endoscopy, @k5 gastroscopy, @k6 hysteroscopy, @k7 bronchoscopy, @k8 ICU, @k9 mechanical ventilation, @k10 procedural sedation. Stepwise narrowing on the same platform confirmed the operators: fospropofol alone 167 records, fospropofol OR fospropofol sodium 280 records, and the complete expression 69 records. 69 records were retrieved (searched 21 September 2026).
6. Embase (Ovid Embase; fields: title, abstract, author keywords): (fospropofol:ab,ti,kw OR “fospropofol disodium”:ab,ti,kw OR aquavan:ab,ti,kw OR lusedra:ab,ti OR “gpi 15715”:ab,ti,kw OR “gpi-15715”:ab,ti,kw) AND (sedation:ab,ti,kw OR endoscopy:ab,ti,kw OR gastroscopy:ab,ti,kw OR colonoscopy:ab,ti,kw OR bronchoscopy:ab,ti,kw OR hysteroscopy:ab,t OR “intensive care”:ab,ti,kw OR “mechanical ventilation”:ab,ti,kw OR “critically ill”:ab,ti,k 208 records, 200 after normalised title deduplication (searched 21 September 2026; no language or year restriction).
7. Cochrane CENTRAL (Cochrane Library; searched by default in title, abstract and keywords): `fospropofol OR “fospropofol disodium” OR Aquavan OR Lusedra OR “GPI-15715”` Results were limited to Trials / Cochrane Central Register of Controlled Trials; 146 records (searched 21 September 2026).
8. Web of Science Core Collection (field: Topic): `TS=(fospropofol OR “fospropofol disodium” OR Aquavan OR Lusedra OR “GPI 15715” OR “GPI-15715”` 135 records (searched 21 September 2026; no language or document type restriction; exported as tab-delimited text and normalised to WOS_20260921_clean.csv). (8b) Broadened re-search of the Chinese databases. CNKI was searched by drug term as a subject term (69 records, 68 unique) and the records were then checked against the setting terms one by one; Wanfang was re-searched with the original advanced strategy (44 records, 42 unique). Both re-searches targeted the same databases on the same search day, and the differences in counts reflect the breadth of the strategy and the databases’ live indexing. The re-searched records entered the merged deduplicated set of 531 records and were screened.
9. Registries and supplementary sources. All were searched on 21 September 2026 (except PROSPERO; see below). PROSPERO does not provide a stable, reproducible bulk-search export, so only the specific registration records directly related to this question were checked (CRD42024618153, CRD42023477740; query terms fospropofol, ciprofol and the Chinese drug name); that source did not contribute to systematic screening and its records did not enter the deduplication or screening pool. ClinicalTrials.gov was searched with the term fospropofol (33 records); WHO ICTRP with fospropofol (140 records); and SinoMed with “fospropofol” (35 records). The CNKI and Wanfang conference proceedings search terms are given in the main text and supplementary material. Registry records do not enter the quantitative synthesis unless they are published and meet the pre-specified PICO criteria.
10. Deduplication rules. Records were first merged by exact DOI or PMID match. When neither was available, titles were normalised by removing leading and trailing numbering, whitespace, full-width and half-width punctuation differences, case differences and hyphens before comparison. For the same record, the version with the most complete metadata was retained, and all source database labels were kept in the source_db field. One point about how that field should be counted: source_db records whether a given unique record is present in a database, not that database’s raw hit count. The labels therefore total 838 (unique record × source database) rather than 859. The difference of 21 is made up of duplicate titles within a single database (Embase 6, VIP 8, Web of Science 3, Europe PMC 2, Cochrane CENTRAL 2; PubMed, CNKI and Wanfang had none). The 859 source records therefore reduce to 531 unique records after within-database and cross-database deduplication of normalised titles, and this chain can be checked record by record against the eight database export files and deduplicated_20260921.csv supplied with this submission. The first-step search of the five core databases returned 327 retrieved records and 214 after deduplication; the broadened re-search combined with the extended standard databases gave 859 source records and 531 after deduplication. The two sets of counts correspond to search strategies of different breadth (see item 8b) and are not interchangeable.

### Search counts and deduplication

We report two sets of search counts, corresponding to the narrower first-step search and the broadened unified re-search. First step: the five core databases reported 338 records in total (Europe PMC 144, PubMed 55, CNKI 27, Wanfang 43, VIP 69), of which 327 were retrieved; after title-based deduplication (normalised comparison) 214 remained. Of the 327 retrieved records, 113 were duplicates: 20 were duplicate titles within the same database and 93 were cross-database duplicates (cross-database overlap involved 82 records, comprising Europe PMC*↔*PubMed 52, CNKI*↔*VIP 14, CNKI*↔*Wanfang*↔*VIP 11, and Wanfang*↔*VIP 5). Of the 214 records, 200 were excluded at title and abstract screening, 14 full texts were assessed, 2 were excluded at the full-text stage, and 12 studies were included.

### Two limitations merit emphasis

(i) Eleven Wanfang records could not be retrieved (only 32 of 43), so they did not enter the original deduplication and screening pool. To avoid missing studies, a broad Wanfang search for “fospropofol” was then run (614 records, about 98 relevant, screened record by record) and identified no additional eligible study (see the supplementary Wanfang search record), but this does not prove that the complete content of those 11 records was covered by the supplementary search. (ii) CNKI, Wanfang and VIP do not provide stable snapshots of search results or an exportable search history, and their hit counts change with database updates, synonym list adjustments and ranking algorithm changes, so the Chinese database counts cannot be reproduced exactly by another person at a different time. This limitation arises from the platform, not from the completeness of the search strategy. To allow record-by-record checking, the supplementary material provides the search date, hit count and exported full records for all five databases, together with the deduplication mapping. Counts for the English-language databases are fully reproducible.

## Appendix 2. Definition of the review scope

The review included only sedation outside the operating room, defined operationally as sedation given for purposes other than general anaesthesia induction or maintenance, covering continuous sedation during ICU mechanical ventilation and procedural sedation. Trials of general anaesthesia induction or maintenance were explicitly excluded to avoid overlap with the 2026 induction metaanalysis [23]. Chinese- and English-language studies were included together and analysed by setting. ICU sedation is not classified as out-of-operating-room anaesthesia (NORA) in the narrow sense under some classification systems, and we used the broader definition; because the two settings showed a significant interaction for hypotension (*P <* 0.001), the conclusions for each are stated separately in the main text.

## Appendix 3. PRISMA 2020 checklist

| No. | Item | Reporting in this review |
| --- | --- | --- |
| 1 | Title | Identified as a systematic review and meta-analysis. |
| 2 | Abstract | Structured abstract (background, methods, results, conclusions). |
| 3 | Rationale | Introduction (importance of sedation, formulation limitations of propofol, pharmacology of fospropofol disodium and the evidence gap). |
| 4 | Objectives | Primary and secondary objectives and expectations stated explicitly. |
| 5 | Eligibility criteria | PICOS (Methods 2.3). |
| 6 | Information sources | Europe PMC, PubMed, Embase, Cochrane CENTRAL, Web of Science Core Collection, CNKI, Wanfang, VIP, ClinicalTrials.gov, WHO ICTRP, SinoMed, CNKI conference proceedings and Wanfang conference proceedings; all searched on 21 September 2026. |
| 7 | Search strategy | Appendix 1 (full strategies for every database; counts for the English-language databases are reproducible and the reproducibility limitation of the Chinese databases is described in Appendix 1). |
| 8 | Selection process | Two reviewers screened independently with third-party adjudication; screening records are provided as supplementary material. |
| 9 | Data collection process | Two reviewers extracted data independently into a standardised form; the extraction form is provided as supplementary material. |
| 10 | Data items | Outcome definitions, event counts and numbers analysed (Table 2). |
| 11 | Risk of bias | RoB 2 across five domains; all 12 studies were assessed by two reviewers from full-text sources (five English-language studies: Feng 2026, Gao 2025, Liu 2026, Gao 2024 and Candiotti 2011; seven Chinese-language studies). The scoring table is provided as supplementary material. |
| 12 | Effect measures | OR for all binary outcomes (random effects); Peto OR reported for outcomes with a high proportion of zero cells as a robustness analysis; the primary outcome is also reported as RD with NNT/NNH. |
| 13 | Synthesis methods | DerSimonian-Laird random effects (all outcomes) with Peto as sensitivity analysis; $I^2$, $\tau^2$, subgroup interaction, sensitivity analysis and TSA. |
| 14 | Reporting bias | The registered protocol set a threshold of at least 10 studies; hypotension included 10 and therefore reached the threshold. The Harbord test, which is appropriate for binary outcomes, is the formal test (intercept 1.55, $P = 0.27$; the change is a protocol deviation, listed in section 2.1), with the originally specified Egger's test (intercept 0.43, $P = 0.76$) also reported for comparison; no test was performed for injection pain because of the large number of zero events. |
| 15 | Certainty of evidence | GRADE (Table 4, with downgrading reasons given for each outcome). |
| 16 | Study selection | PRISMA flow diagram (Figure 2). |
| 17 | Study characteristics | Table 1 (including a blinding column). |
| 18 | Risk of bias in studies | Figure 4. |
| 19 | Results of individual studies | Table 2 and forest plots (Figures 5 to 10 and 11). |
| 20 | Results of syntheses | Pooled estimates, heterogeneity, subgroup interaction, sensitivity analyses and TSA for each outcome. |
| 21 | Reporting biases | Hypotension: the protocol threshold was met (10 studies), and a funnel plot and the Harbord test (intercept 1.55, $P = 0.27$; the change is a protocol deviation, listed in section 2.1) were performed, with the originally specified Egger's test (intercept 0.43, $P = 0.76$) reported alongside; power is still limited at $k = 10$. |
| 22 | Certainty of evidence | Table 4. |
| 23 | Discussion | Main findings, mechanism, limits of extrapolation by setting, implications for guidelines and limitations. |
| 24 | Registration and protocol | Prospectively registered (CRD420261507586, effective 16 September 2026); all searches were run on 21 September 2026; data extraction, risk-of-bias assessment and statistical analysis were undertaken after registration; see Methods 2.1. |
| 25 | Support | No external funding. |
| 26 | Competing interests | None. |
| 27 | Availability of data and code | Analysis scripts, extracted data, screening table and RoB tables are provided as supplementary material; the protocol is available in PROSPERO (CRD420261507586). |

## Appendix 4. Main types of excluded studies

Two specific studies were excluded at the full-text stage. (i) Hu 2026 (Journal of Practical Medicine; painless ultrasound-guided transbronchial needle aspiration) [30]. The study gave fospropofol disodium 12.5 mg/kg or propofol 2 mg/kg at induction of anaesthesia, then injected rocuronium and inserted a laryngeal mask for mechanical ventilation; the English title also states “during anaesthesia induction” and the keywords include “general anaesthesia”, so it is a general anaesthesia induction and maintenance trial and was excluded under the pre-specified criteria. (ii) Zhang Yi 2026 (Chinese Journal of Clinical Pharmacology and Therapeutics) [36]. This was a dose-response study in which all three sex subgroups received fospropofol disodium without a propofol comparator, so it did not meet the requirement for a propofol comparator and was excluded. The supplementary Wanfang search also identified a 2025 master’s thesis by Li Xiaoning on painless fibreoptic bronchoscopy in older patients, which explored the dose-response of fospropofol disodium combined with alfentanil without a propofol comparator and was excluded before full-text assessment. All of these decisions were made independently by two reviewers during screening or full-text assessment and confirmed by adjudication.

Appendix 4. Main types of excluded studies
| Type | Reason for exclusion |
| --- | --- |
| General anaesthesia induction or maintenance trials | Belong to the existing induction meta-analysis; excluded to avoid overlap |
| Studies without a propofol comparator | Compared only with midazolam, etomidate or placebo |
| Dose-finding and sequential-dose studies | No propofol-comparator event counts (in some cases) |
| Non-randomised studies and reviews | Observational studies, narrative reviews and conference abstracts |

### Registered trial excluded after assessment at the search stage

NCT01401049 (”Preventing Propofol Injection Pain: Prospective Randomized Trial Comparing Propofol Versus Fospropofol”; USA; randomised, parallel, single-blind; 116 participants actually enrolled; primary outcome the incidence and intensity of injection pain). This trial compared fospropofol disodium with propofol head to head, but the comparator arm was a mixture of propofol and lidocaine, and the registry record notes that the original files and patient data were lost because of a hurricane, so no analysable values were provided for the primary outcome or adverse events. It could not enter the quantitative synthesis. No corresponding published article was identified; even if the original data became available, the propofol-plus-lidocaine comparator would have to be handled separately from a plain propofol comparator.

## Appendix 5. Data sources and verification procedure

### Verification of event counts (all 12 studies)

Every event count in the included studies was checked against the full-text original publication: Feng 2026, Gao 2025 and Liu 2026 from full-text PDF or PMC full text; Gao 2024 and Candiotti 2011 from the publisher’s full-text PDF (the hypotension counts were taken from the treatment-emergent adverse event rows of Table 3); and the seven Chinese studies from full-text PDFs (Xiong 2026, Bai 2025, Zhan 2025, Xu 2025, Tang 2026, Zhang 2025 and Fan 2024). Liu 2026 is a five-centre RCT preprint (Research Square, not peer reviewed), and its hypotension and hypoxaemia event counts appear directly in the preprint tables (Table 3 “Post-dose 5-min hypotension” and Table 2 “Hypoxemia”, both reported as counts with percentages). We also checked the counts against the percentages: PIH 9/213 and 32/213 correspond to 4.22% and 15.02% (9*/*213 = 4.23%, 32*/*213 = 15.02%), and hypoxaemia 9/213 and 37/213 correspond to 4.23% and 17.37% (9*/*213 = 4.23%, 37*/*213 = 17.37%), so the counts and percentages agree. The data are direct counts, but the source is a preprint that has not been peer reviewed; the sensitivity and exclusion analyses are reported in section 3.12 and Appendix 6.

### Independent two-reviewer extraction and checking

Event counts were extracted independently by two reviewers according to the pre-specified outcome definitions and compared cell by cell, with disagreements resolved by discussion and adjudication against the original articles. Screening agreement was 98.6% (211/214), with three disagreements resolved by discussion; risk-of-bias disagreements were resolved to the more conservative judgement. One of the three disagreements concerned eligibility at the full-text stage (the general anaesthesia induction trial described in Appendix 4), and the adjudication accepted the case for exclusion. Three further decisions made during checking were: (1) the “hypoxaemia” outcome in Bai 2025 (5/70 versus 19/66) was included as a respiratory outcome of similar construct; (2) the composite “respiratory depression/cough” outcome in Tang 2026 (1/52 versus 0/29) could not be separated and was not pooled; and (3) both arms of Gao 2024 had zero pruritus/paraesthesia events, so the study was not pooled.

### Recomputation of pooled estimates (methodological note)

All pooled effect sizes were recalculated by a second script (analyze_v2.py) with independently re-entered data, and the results agreed with the primary English script (analyze_en.py). Both scripts use the same estimation formulas (the DerSimonian-Laird and Peto implementations share the same source), so this recomputation verifies the consistency of data entry and computation rather than providing methodological independence. Methodological robustness is addressed by the leave-one-out analysis, the parallel reporting of estimators and corrections, and the sensitivity analyses including exclusion of the preprint. The reasons for the difference between Peto and random-effects estimates for outcomes with zero cells are described in Appendix 6.

### Search counts (21 September 2026)

The narrower first-step search covered the five core databases: Europe PMC 144, PubMed 55, CNKI 27, Wanfang 43 and VIP 69 (338 in total). Of these, 327 records were retrieved (32 of the 43 Wanfang records), 214 remained after deduplication, two reviewers excluded 200 records at independent screening, 14 full texts were assessed, 2 were excluded at the full-text stage and 12 were included. The extended search gave 859 merged source records and 531 after deduplication, and no additional study was included. All search strategies, exported records and the deduplication mapping are provided in the “search records” package of the supplementary material.

## Appendix 6. Zero-event handling and sensitivity analyses for sparse outcomes

Injection pain, respiratory depression/hypoxaemia and pruritus/paraesthesia contained zero cells. The primary analysis used a DerSimonian-Laird random-effects model for all outcomes and applied a 0.5 continuity correction only to studies whose 2 *×* 2 table genuinely contained a zero cell, consistent with the registered protocol (a study with one event in an arm but no zero cell was not corrected; see section 2.6); as a sensitivity analysis, the table also reports the Peto estimate, which requires no continuity correction.

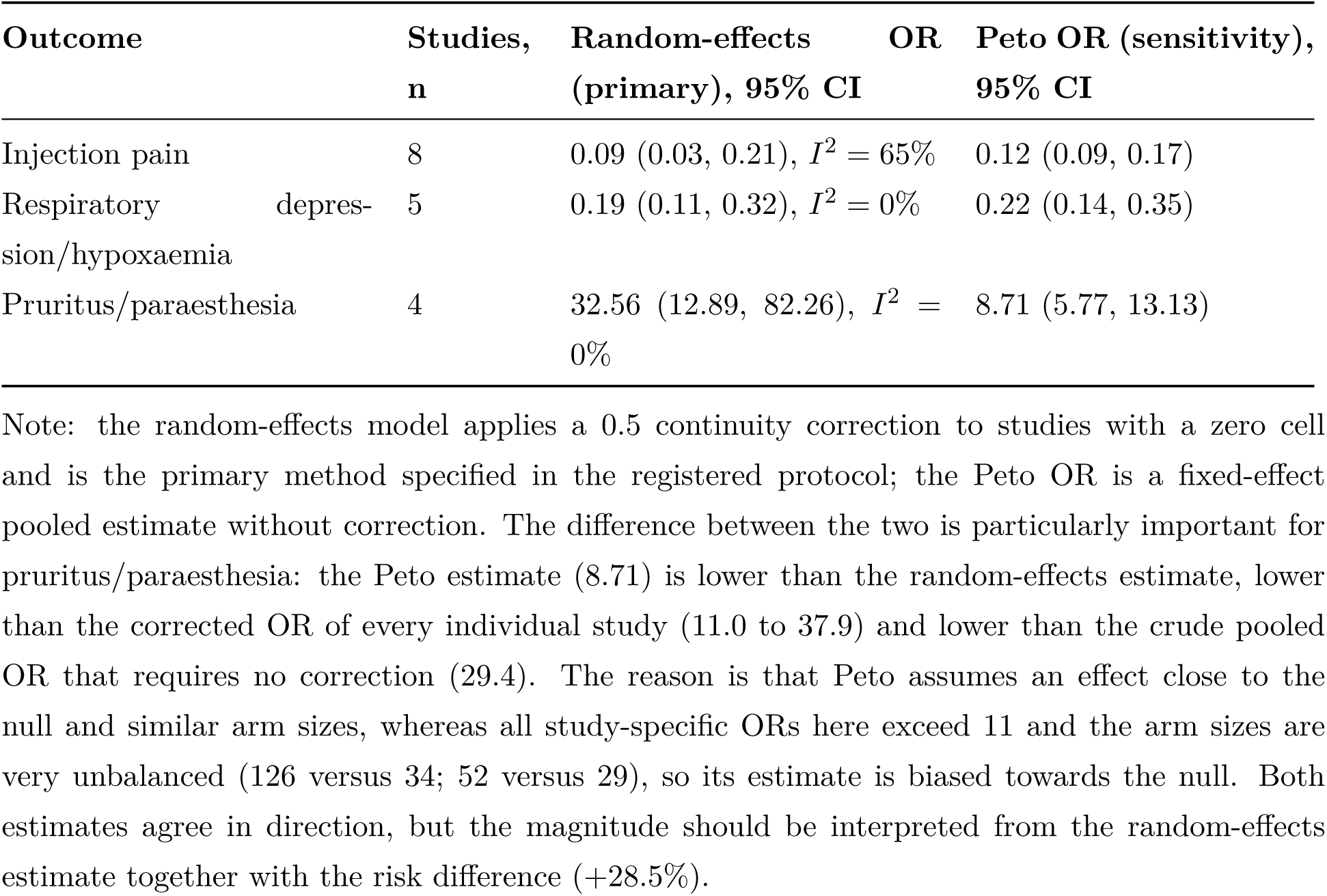

### The two sensitivity analyses for injection pain (reported separately)

**The two sensitivity analyses for injection pain (reported separately).**
| Analysis | Studies,<br>n | Pooled OR (95% CI) | Heterogeneity |
| --- | --- | --- | --- |
| Primary analysis (random effects) | 8 | 0.09 (0.03, 0.21) | $I^2 = 65\%$, $\tau^2 = 0.99$ |
| Studies with observed events only (random effects) | 5 | 0.13 (0.05, 0.35) | $I^2 = 74\%$, $\tau^2 = 0.97$ |
| Excluding the Liu 2026 preprint (random effects) | 7 | 0.09 (0.03, 0.26) | $I^2 = 69\%$, $\tau^2 = 1.28$ |
| Excluding the Liu 2026 preprint (Peto) | 7 | 0.11 (0.08, 0.17) | not applicable |

Note: these analyses use different study sets and estimate different parameters and are not interchangeable.

#### Estimator sensitivity analysis

The table lists the results for every outcome under four pooling methods. The primary analysis is the DerSimonian-Laird random-effects model (consistent with the registered protocol, with a 0.5 continuity correction for studies with zero cells); results are also given for REML estimation of *τ* ^2^ with modified Hartung-Knapp confidence intervals (more robust when *k* is small; the fixed-effect variance is retained when the Hartung-Knapp standard error falls below it), the Mantel-Haenszel method (fixed effect, with no continuity correction) and the Peto method, together with 95% prediction intervals based on *τ* ^2^. All values were recomputed independently by scripts/estimator_check.py.

| Outcome | k | DL random ef-<br>fects (primary) | REML + modi-<br>fied HK | MH OR | 95% predic-<br>tion interval |
| --- | --- | --- | --- | --- | --- |
| Hypotension | 10 | 0.51 (0.28, 0.93) | 0.51 (0.25, 1.03) | 0.45 | 0.09–2.72 |
| Injection pain | 8 | 0.09 (0.03, 0.21) | 0.09 (0.03, 0.25) | 0.09 | 0.01–1.25 |
| Bradycardia | 9 | 0.45 (0.17, 1.18) | 0.45 (0.15, 1.39) | 0.41 | 0.04–5.61 |
| Respiratory depres-<br>sion/hypoxaemia | 5 | 0.19 (0.11, 0.32) | 0.19 (0.09, 0.40) | 0.18 | 0.08–0.45 |
| Hypertriglyceridaemia | 4 | 0.29 (0.13, 0.65) | 0.29 (0.08, 1.08) | 0.28 | 0.05–1.71 |
| Pruritus/paraesthesia | 4 | 32.56 (12.89, 82.26) | 32.56 (7.23, 146.63) | 46.64 | 4.26–249.04 |

Note: the Mantel-Haenszel and Peto methods are fixed-effect analyses; the Mantel-Haenszel estimate applies no continuity correction (zero-cell studies contribute 0 to the numerator or denominator); confidence intervals that are not reported here are given in the main text. The Peto result is also shown in the earlier table, where it deviates markedly from the other methods for pruritus/paraesthesia (8.71 versus values above 30); the explanation is given in the note above.

#### Consistency of direction across methods

Apart from hypertriglyceridaemia, the direction and significance of every outcome were consistent across the four methods. Hypertriglyceridaemia was the only outcome whose conclusion changed between estimators: the DerSimonian-Laird estimate was OR 0.29 (0.13–0.65) whereas REML plus modified Hartung-Knapp gave 0.29 (0.08–1.08), the latter crossing 1. *τ* ^2^ was 0 under both DerSimonian-Laird and REML, so the point estimates agree and the interval differs because with *k* = 4 the modified Hartung-Knapp method uses the *t*_(3)_ quantile and truncates the variance at the fixed-effect level; this review therefore rates the outcome as very low certainty and reports only that it may be reduced under the DerSimonian-Laird model. The prediction intervals indicate that extrapolation warrants caution: the 95% prediction intervals for hypotension (0.09–2.72) and injection pain (0.01–1.25) cross 1, which means that in a different setting or population the effect in a future single study could fall in the null or even the opposite range. This agrees with the finding of a significant interaction between settings for hypotension (*P <* 0.001).

The sensitivity analysis of the Liu 2026 preprint event counts is given in Supplementary Material S6: after reverse perturbation of the two arms (9/32 original, 10/29, 12/22, 15/25) the pooled OR remained between 0.51 and 0.59, and excluding the study entirely gave 0.58 (0.31–1.11).

#### Cohort overlap check

Gao 2024 and Gao 2025 came from the same team and centre, with essentially identical author lists and 60 participants each, but different sedation depth targets (RASS *−*3 to 0 versus *−*4 to *−*5) and different registration information, and Gao 2025 cites Gao 2024 as that team’s earlier work. Exclusion analysis showed that the conclusions for hypertriglyceridaemia and hypotension were unchanged when either study was removed (see section 3.12). Without individual-level data, partial patient overlap cannot be excluded completely, and this is stated in the limitations.

## Appendix 7. PRISMA-S checklist for the search

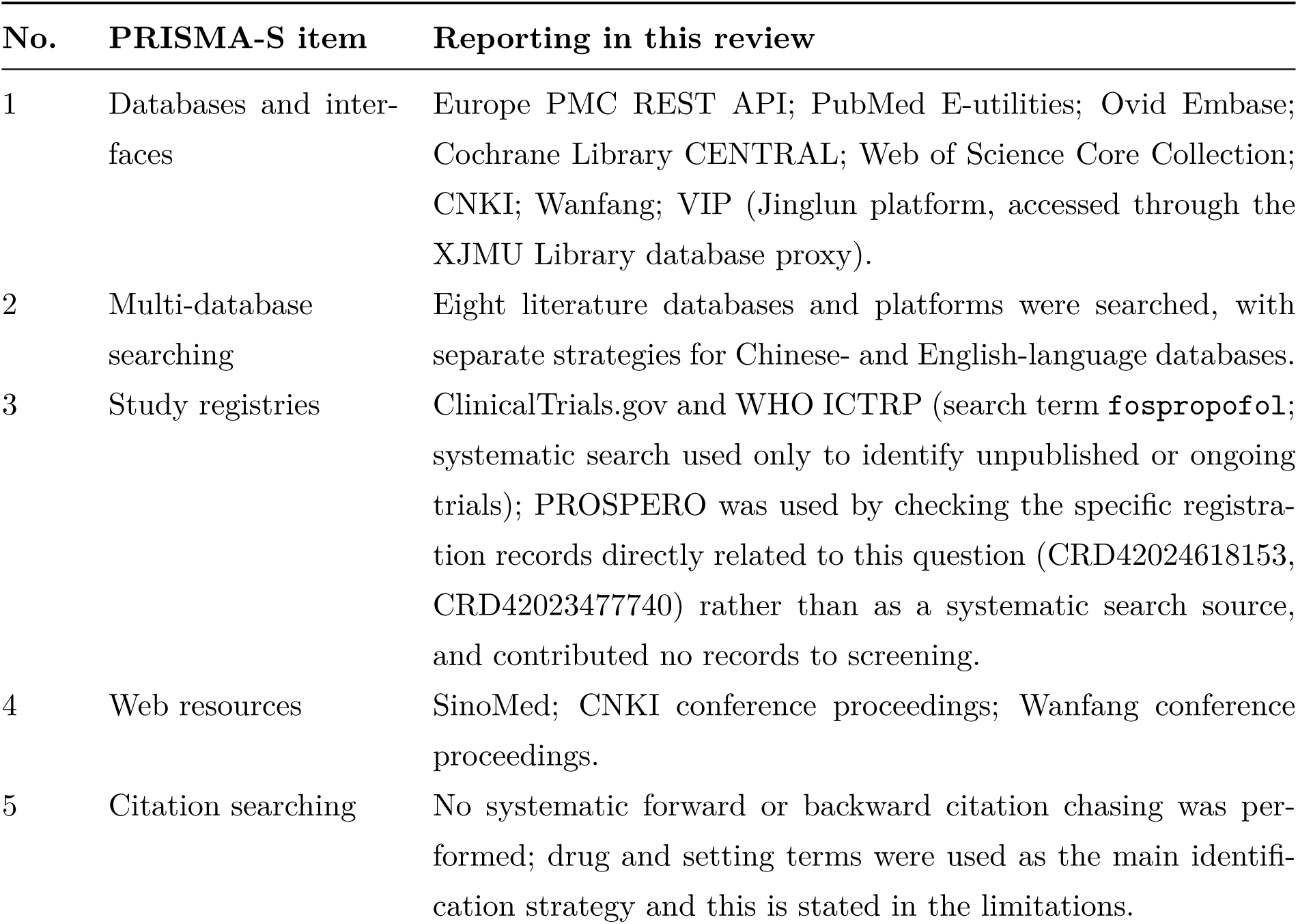
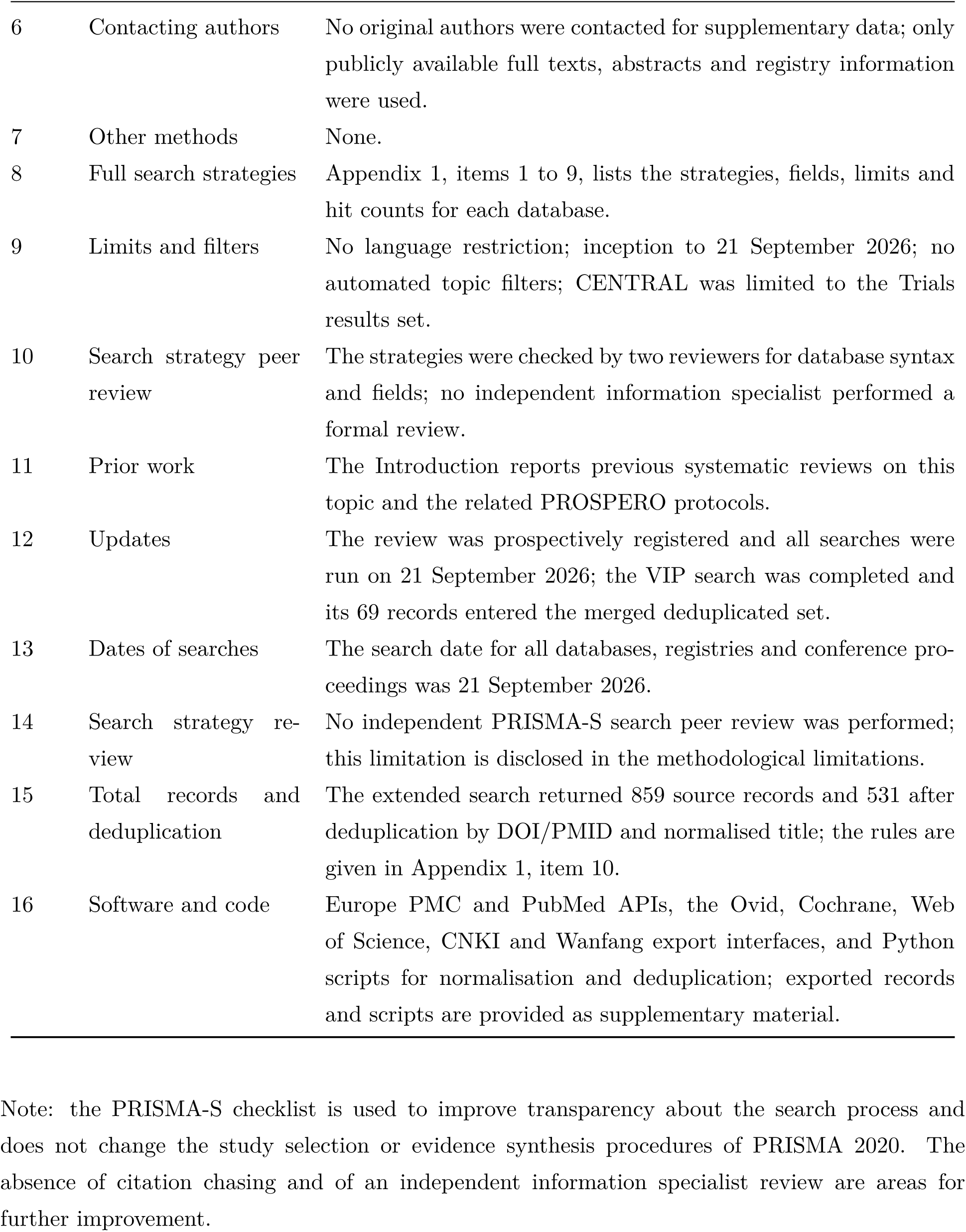

---

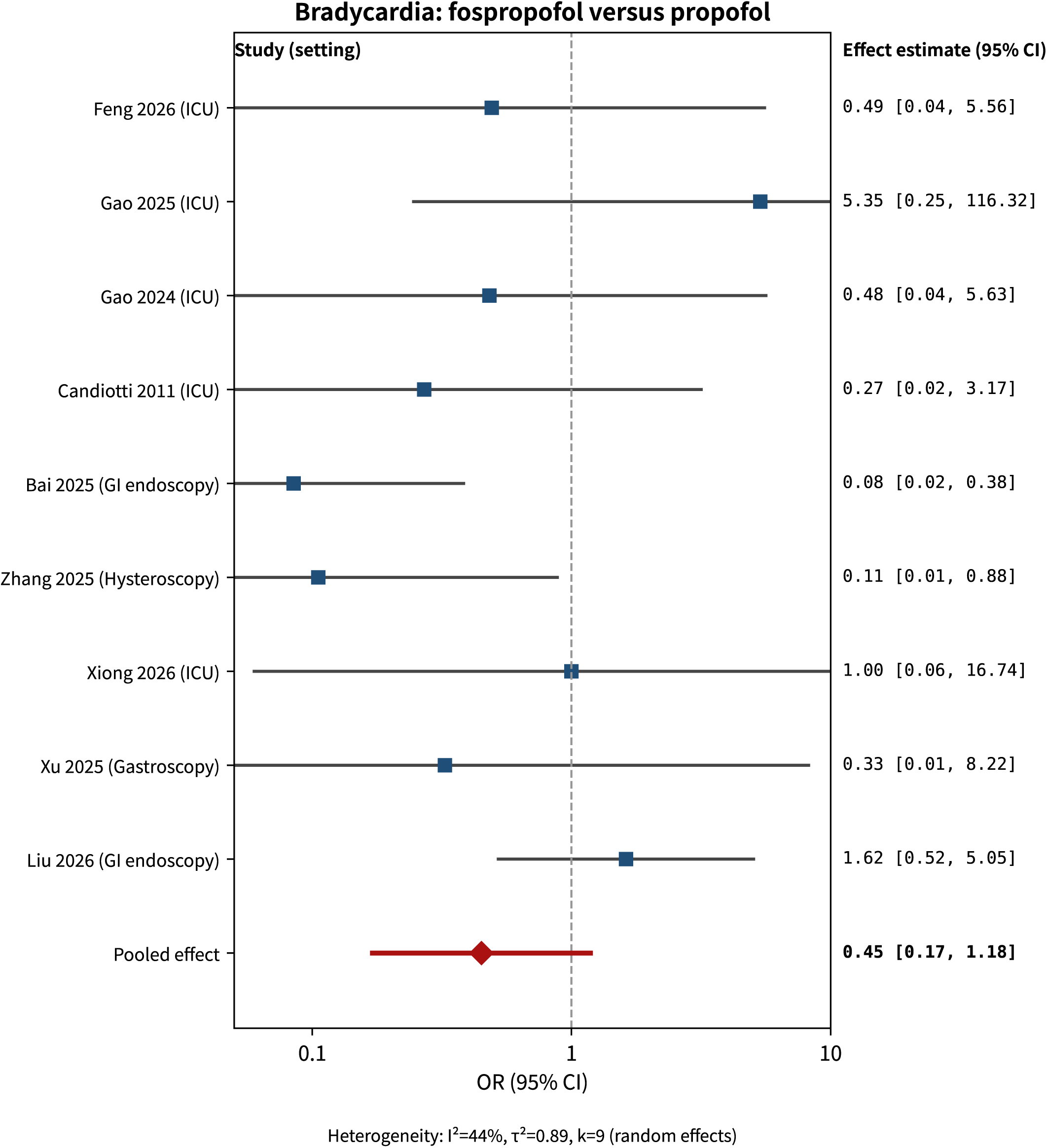

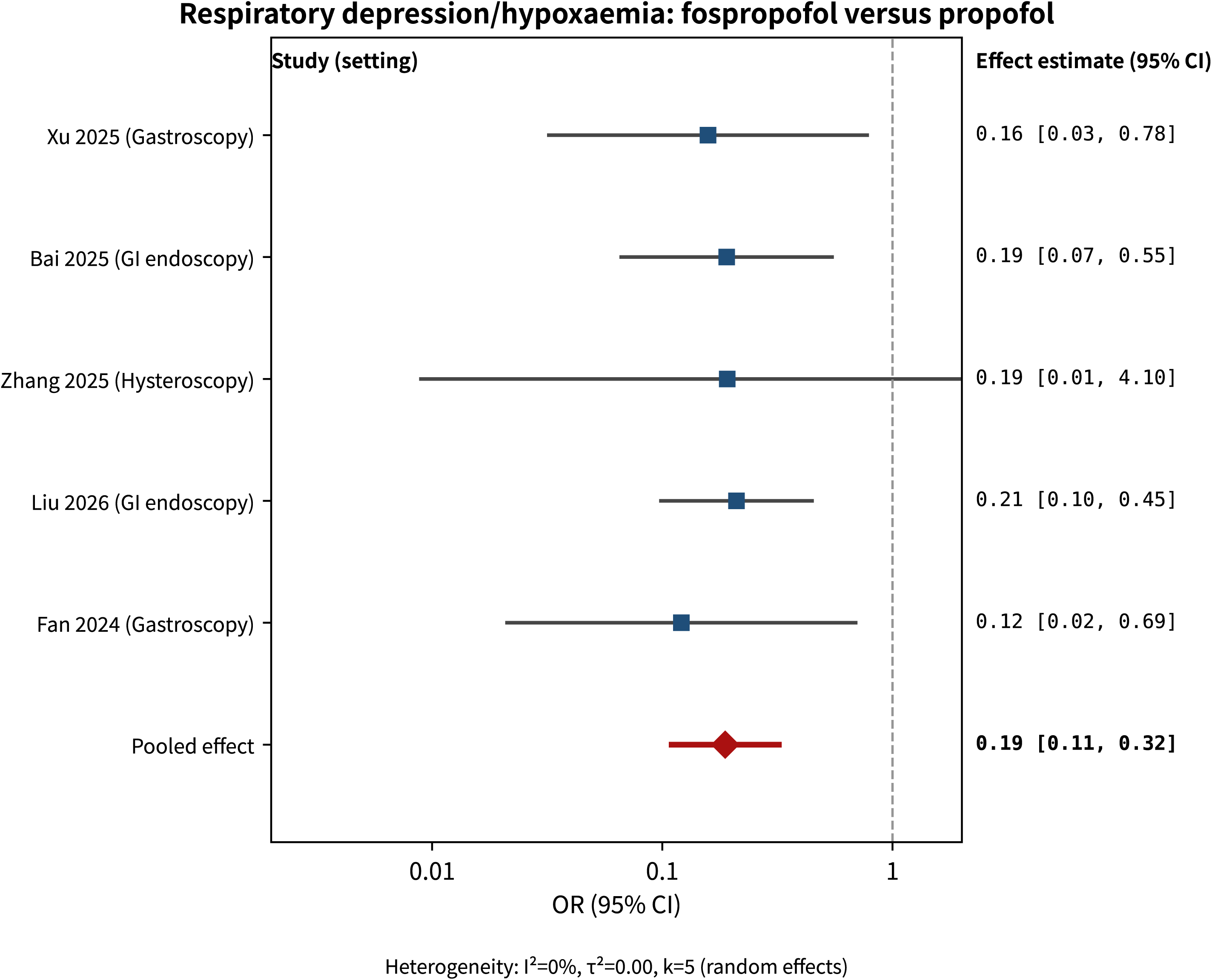

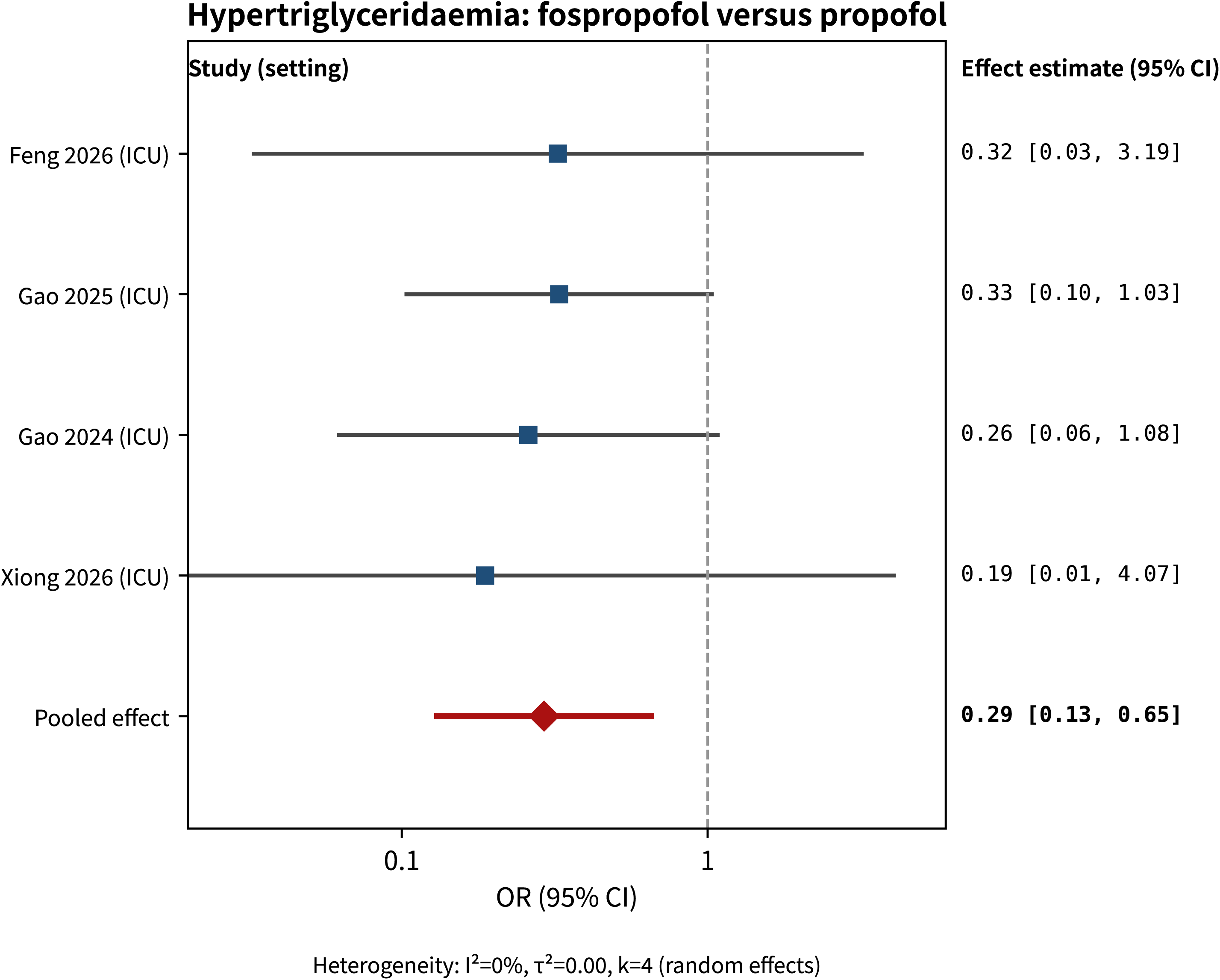

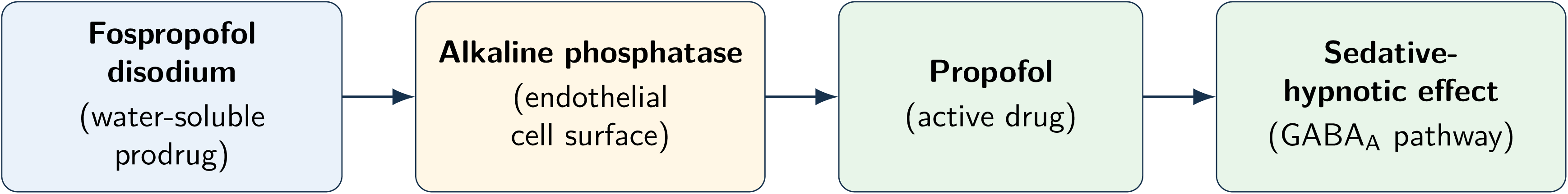

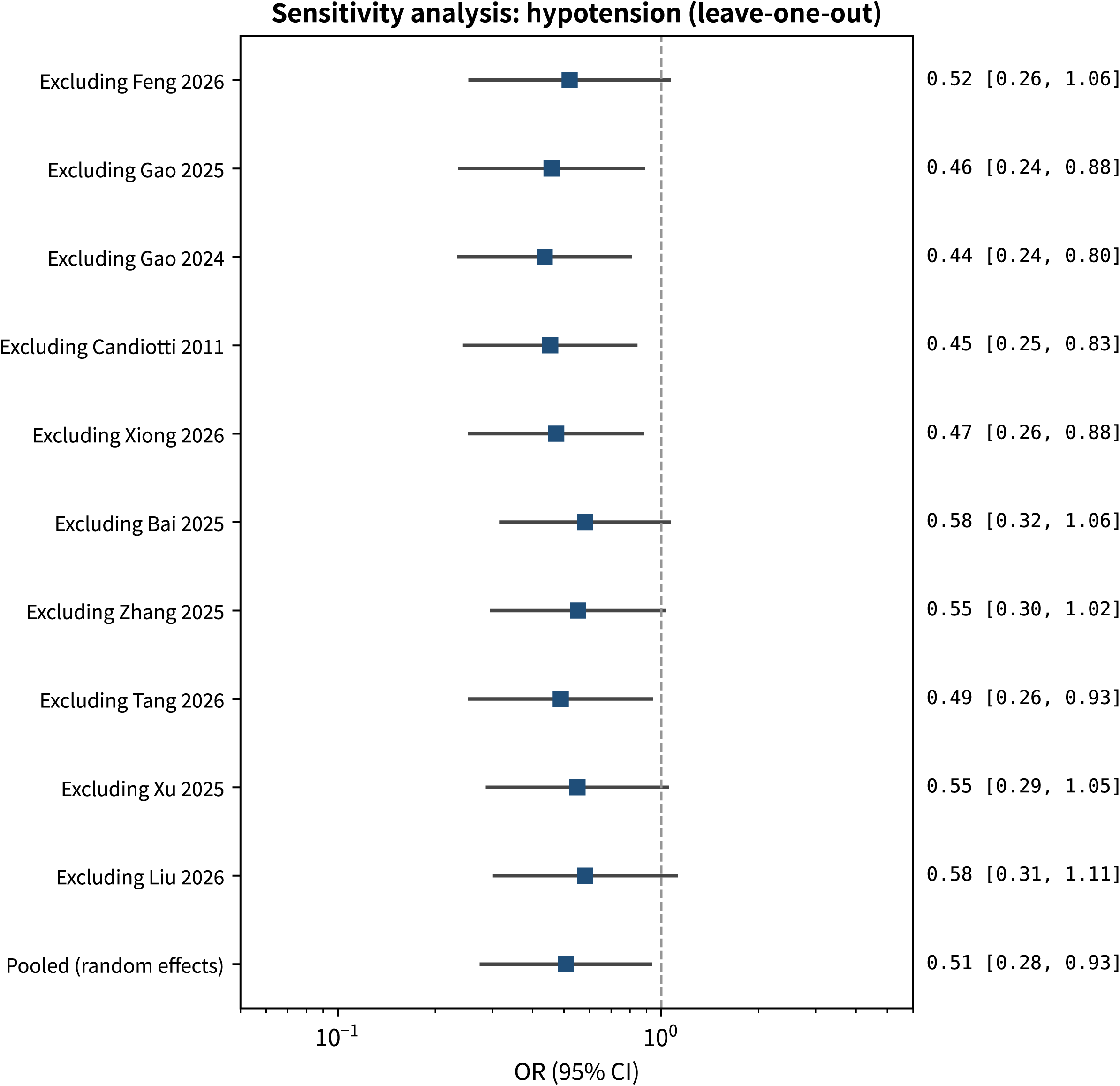

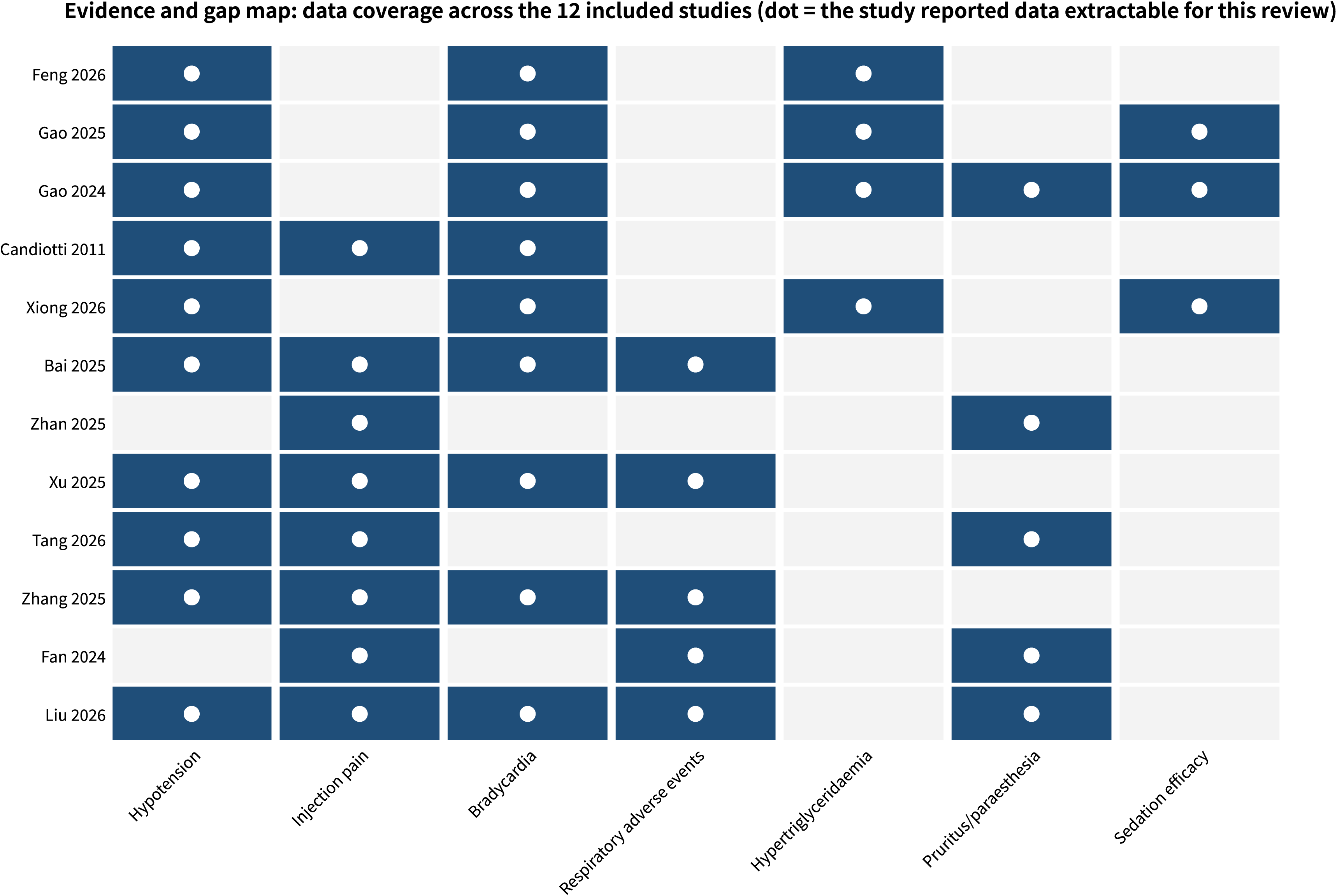

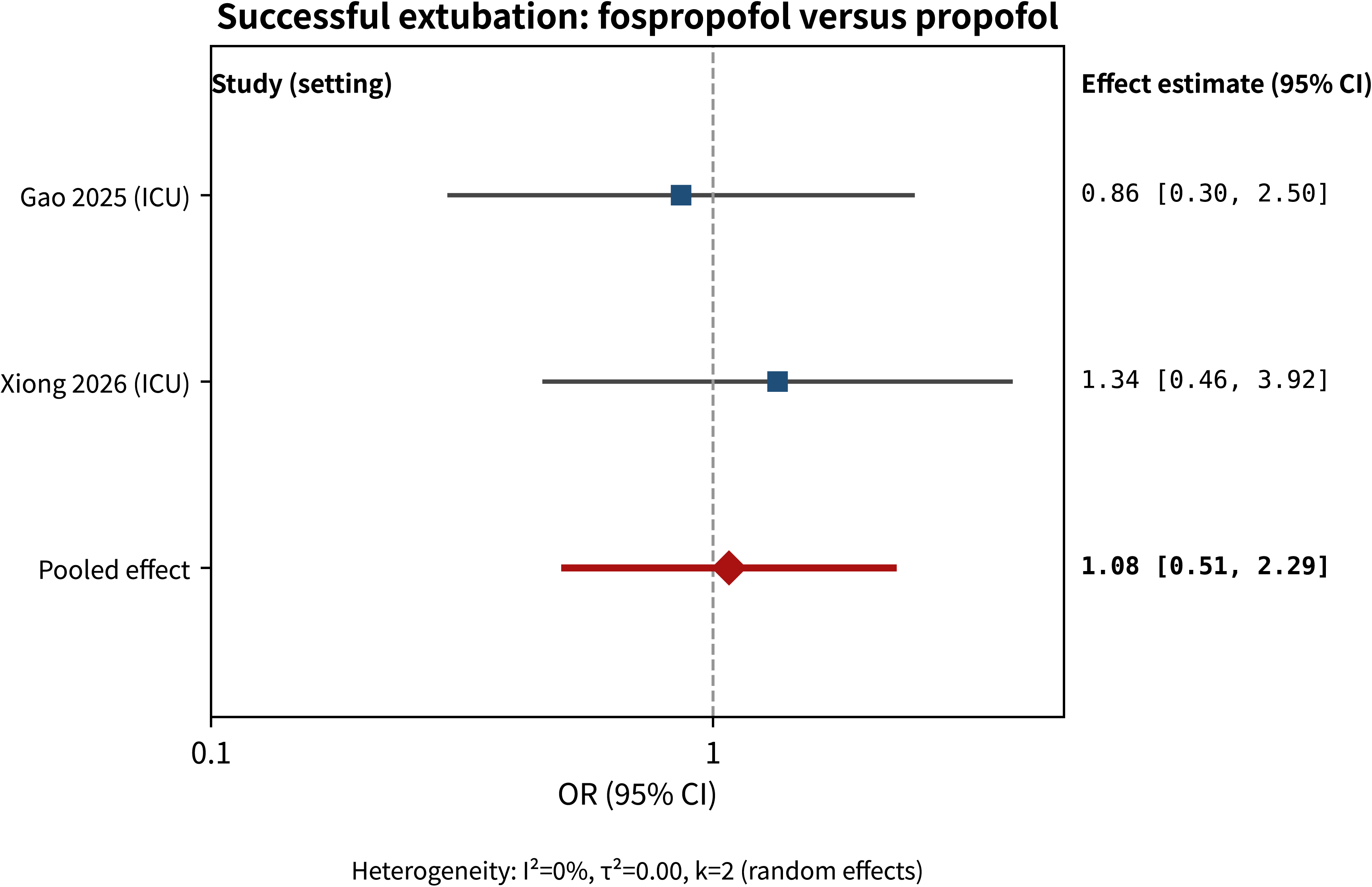

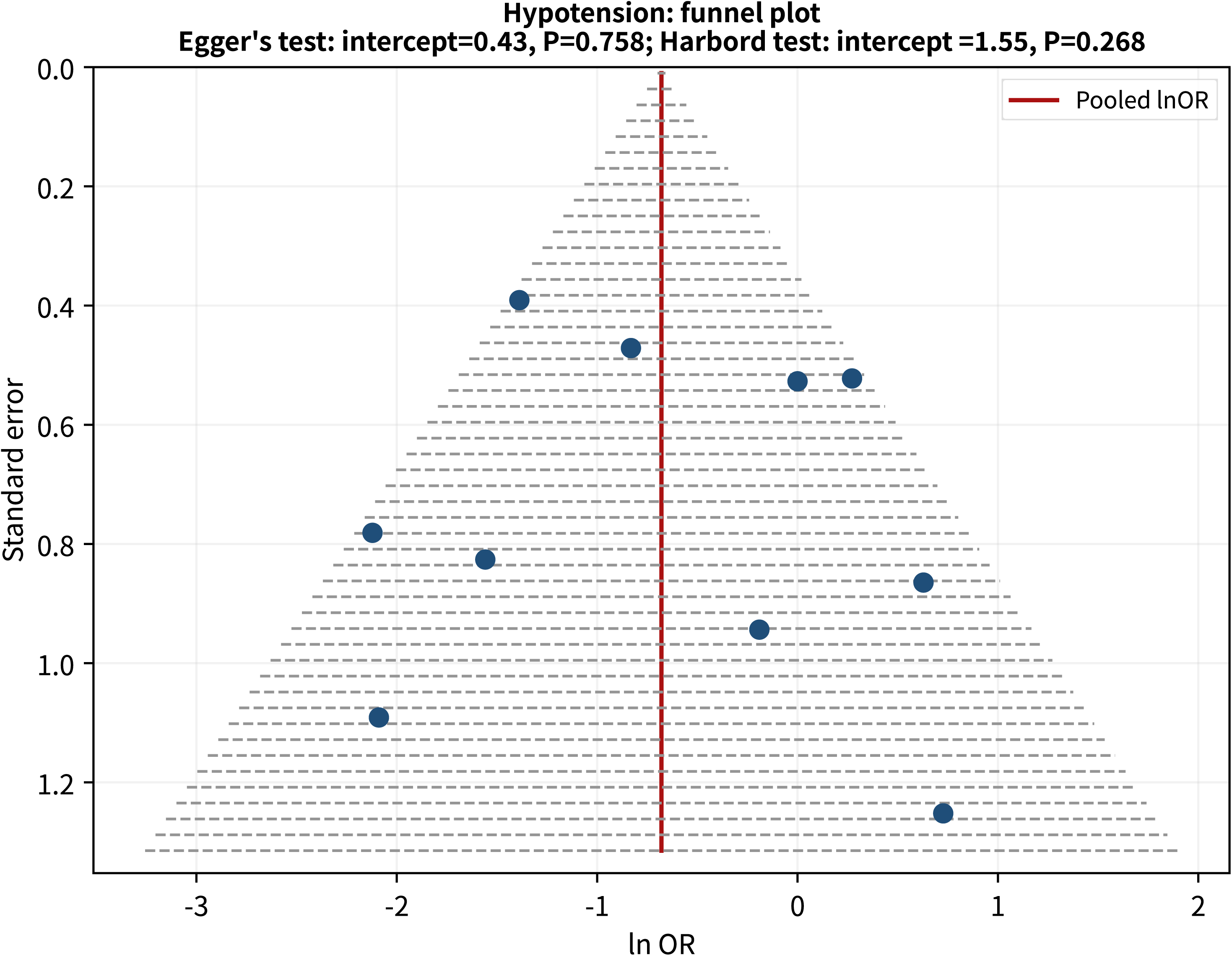

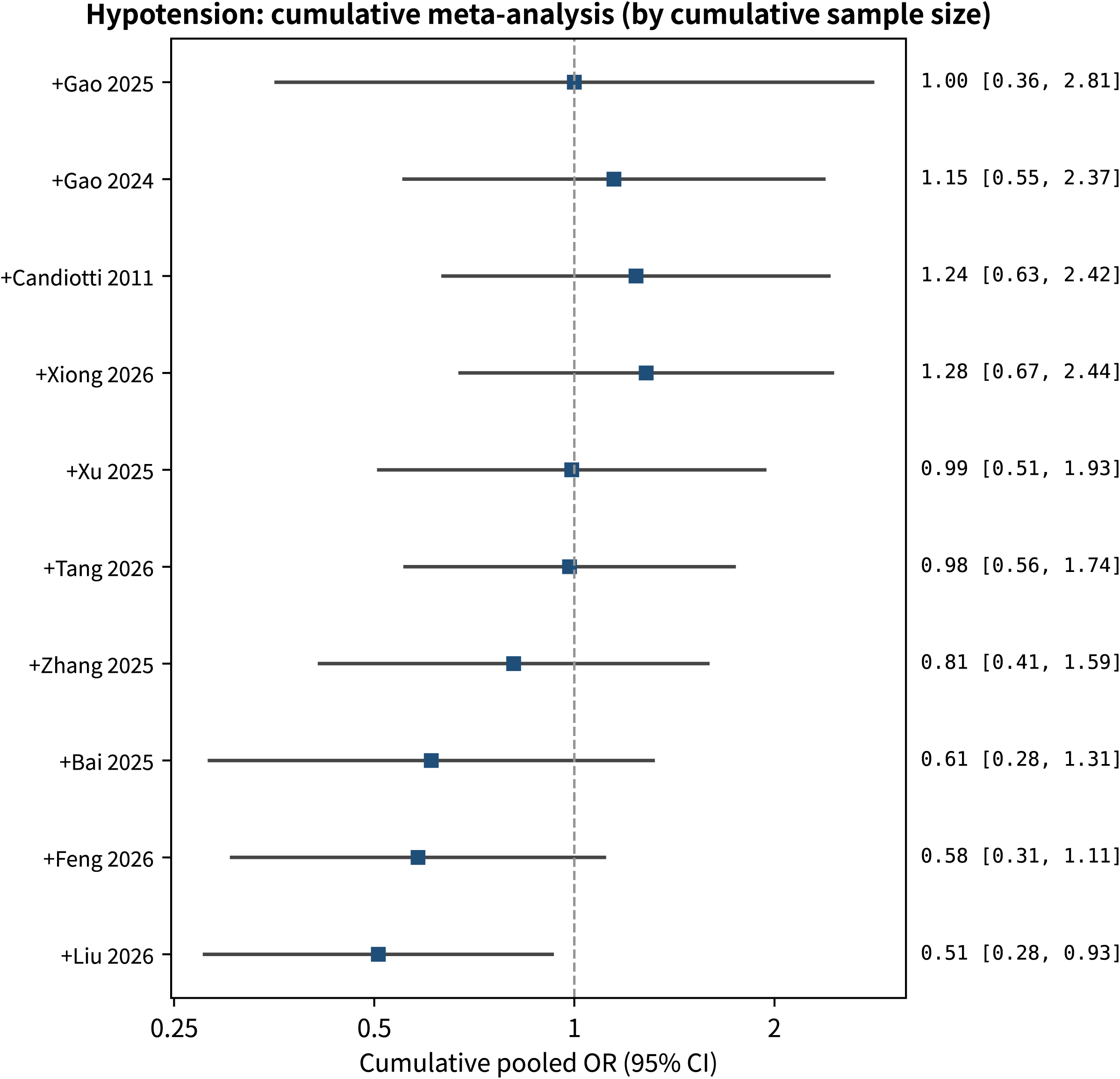

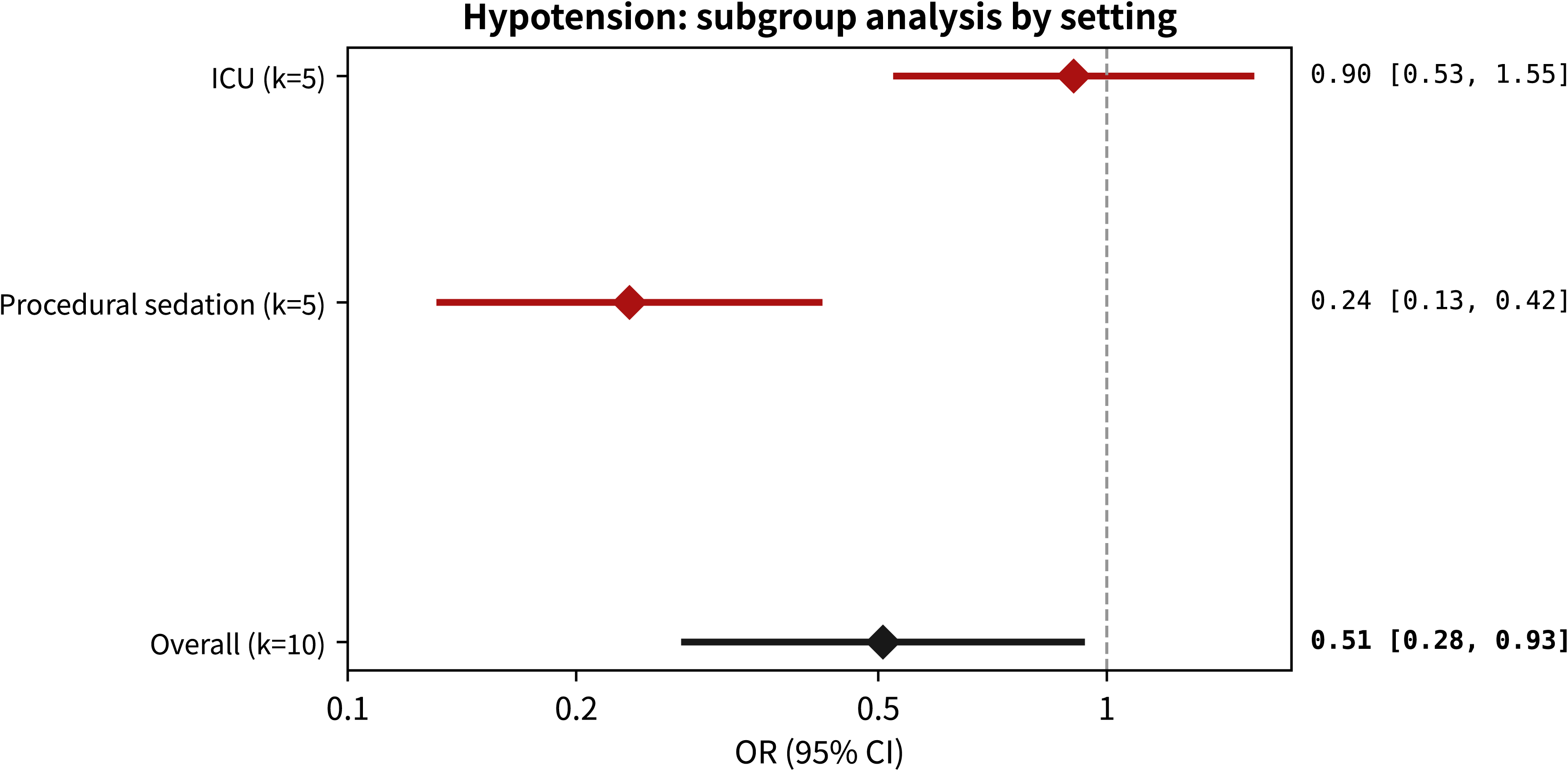

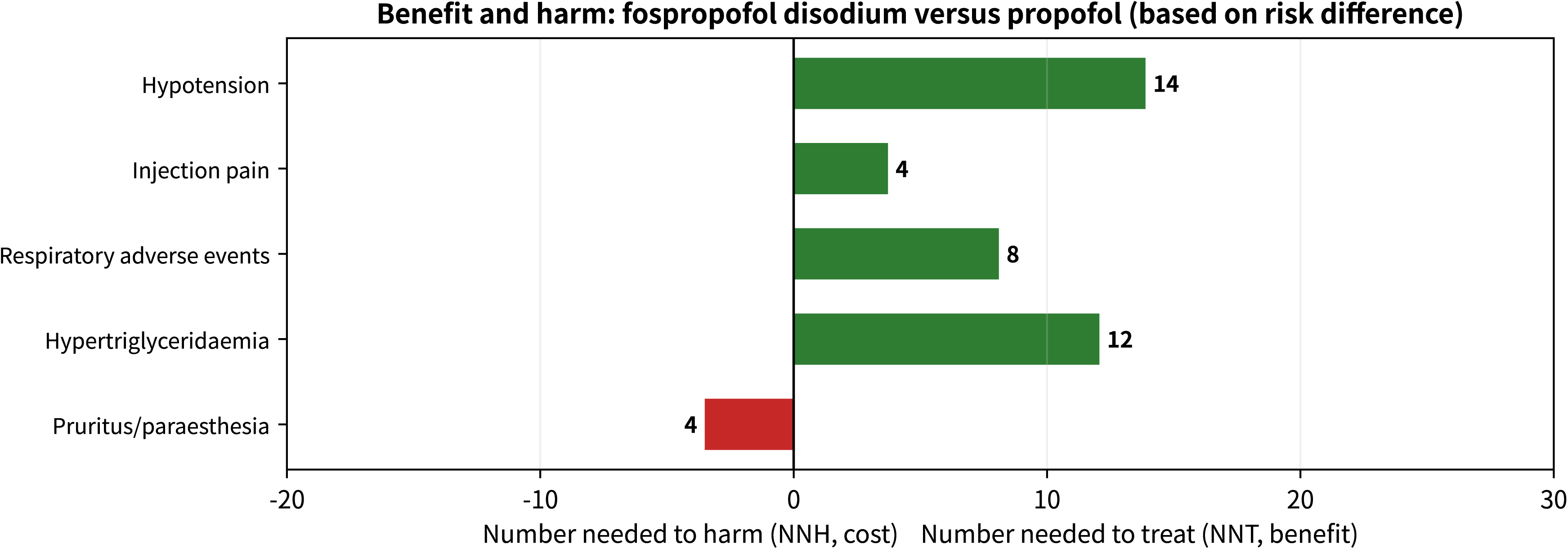

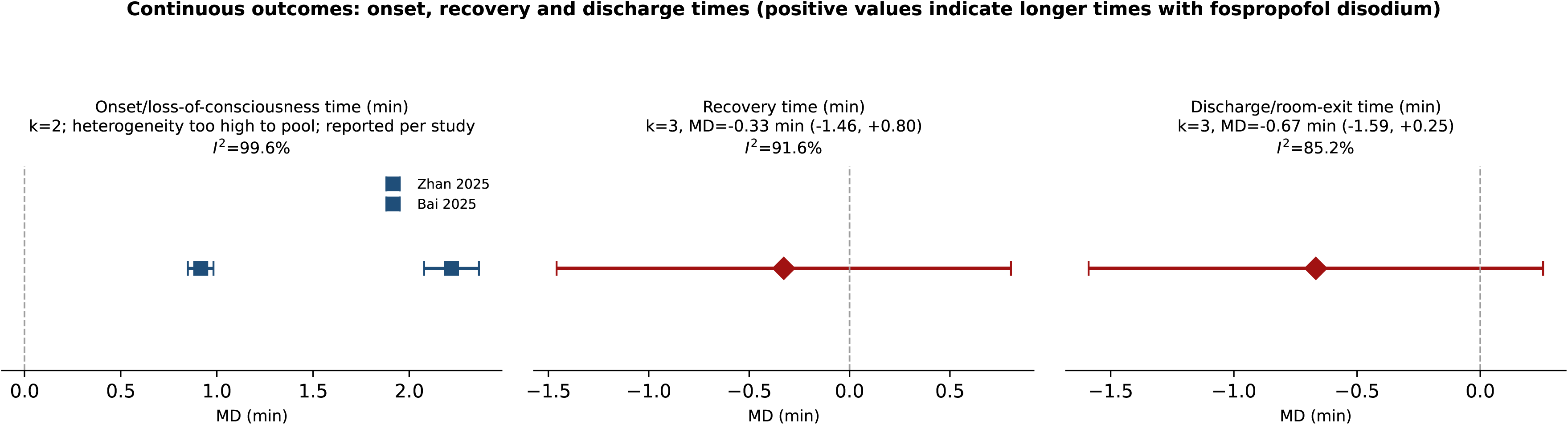

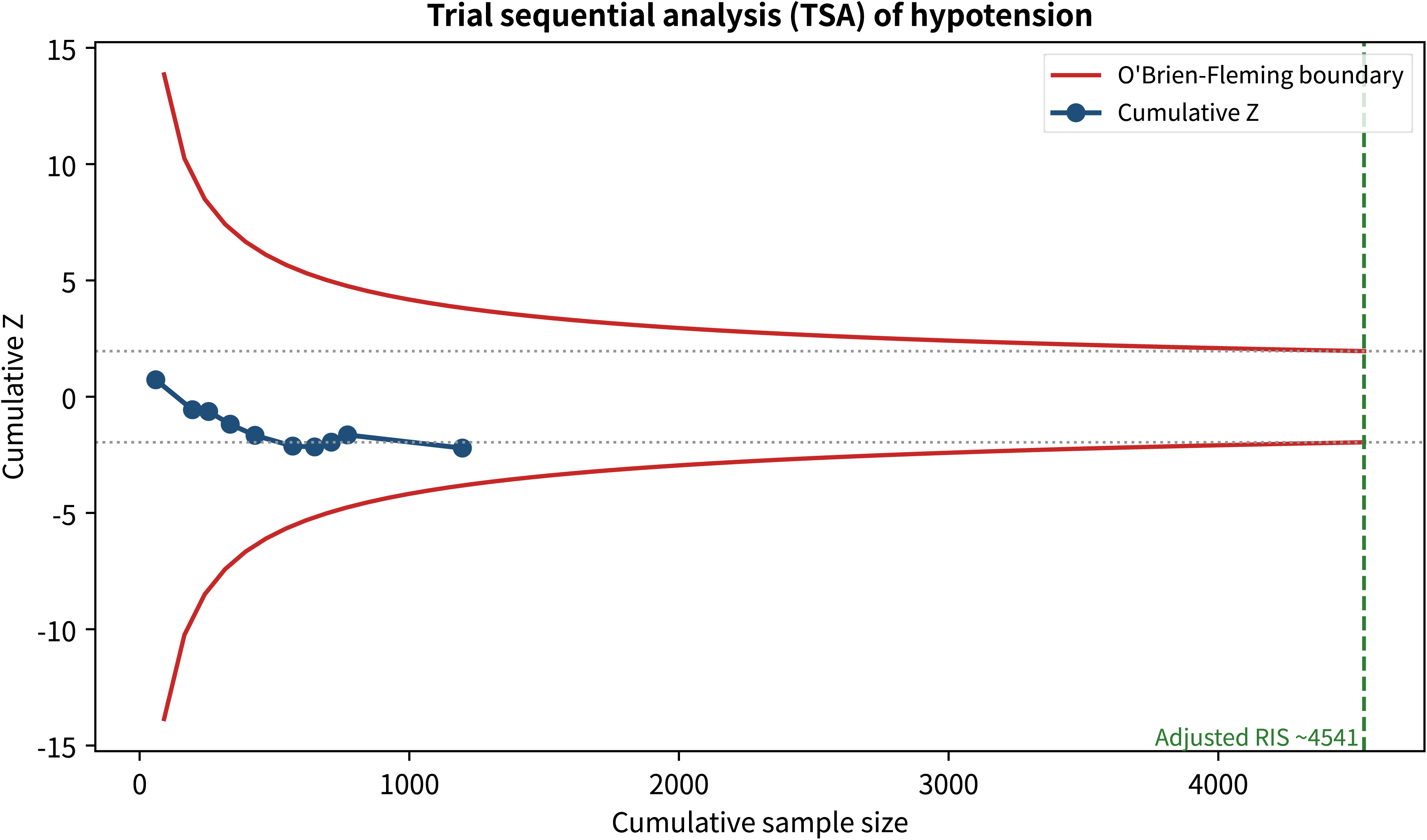

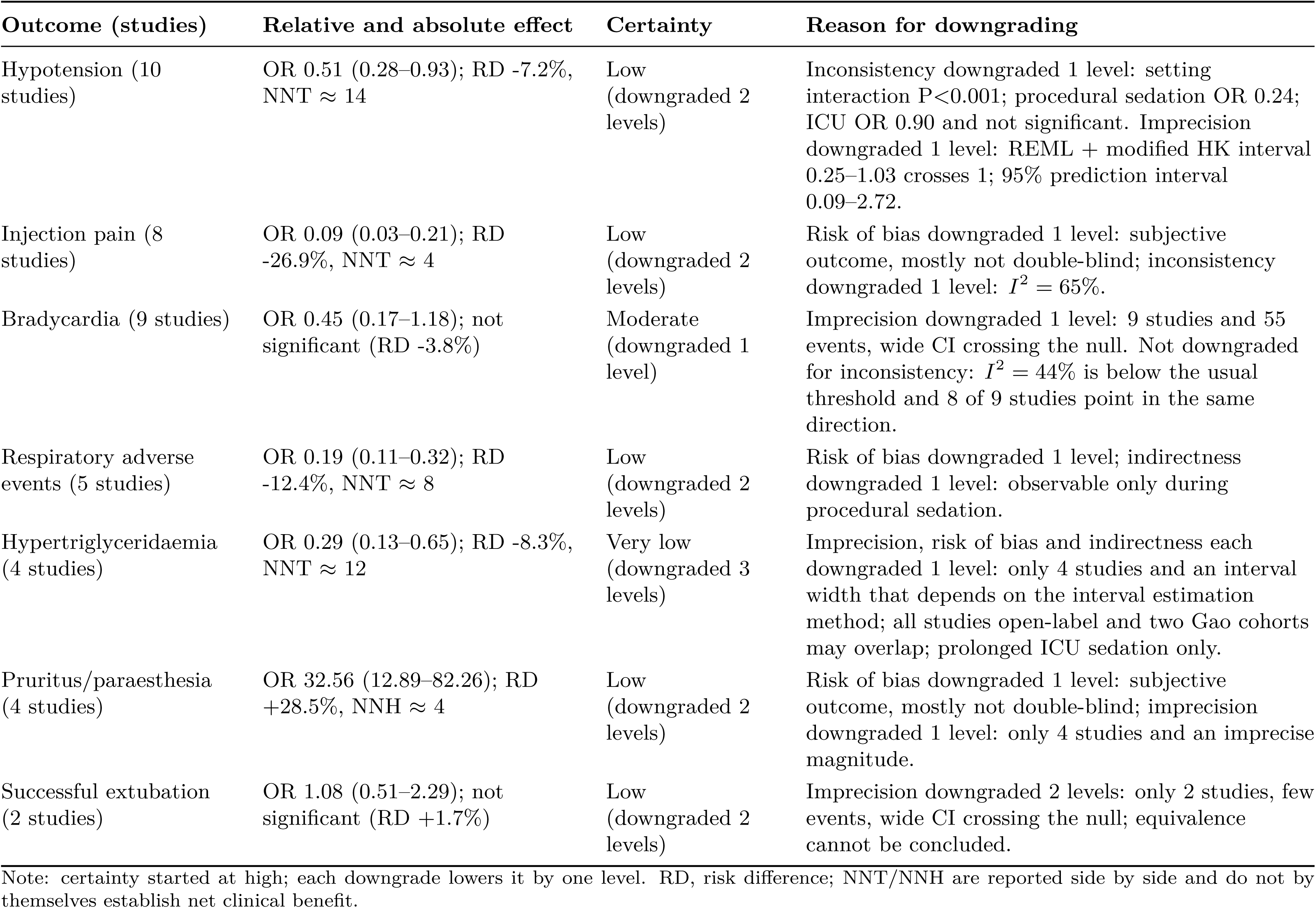

| Outcome (studies) | Relative and absolute effect | Certainty | Reason for downgrading |
| --- | --- | --- | --- |
| Hypotension (10 studies) | OR 0.51 (0.28–0.93); RD -7.2%, NNT ≈ 14 | Low (downgraded 2 levels) | Inconsistency downgraded 1 level: setting interaction $P < 0.001$; procedural sedation OR 0.24; ICU OR 0.90 and not significant. Imprecision downgraded 1 level: REML + modified HK interval 0.25–1.03 crosses 1; 95% prediction interval 0.09–2.72. |
| Injection pain (8 studies) | OR 0.09 (0.03–0.21); RD -26.9%, NNT ≈ 4 | Low (downgraded 2 levels) | Risk of bias downgraded 1 level: subjective outcome, mostly not double-blind; inconsistency downgraded 1 level: $I^2 = 65\%$. |
| Bradycardia (9 studies) | OR 0.45 (0.17–1.18); not significant (RD -3.8%) | Moderate (downgraded 1 level) | Imprecision downgraded 1 level: 9 studies and 55 events, wide CI crossing the null. Not downgraded for inconsistency: $I^2 = 44\%$ is below the usual threshold and 8 of 9 studies point in the same direction. |
| Respiratory adverse events (5 studies) | OR 0.19 (0.11–0.32); RD -12.4%, NNT ≈ 8 | Low (downgraded 2 levels) | Risk of bias downgraded 1 level; indirectness downgraded 1 level: observable only during procedural sedation. |
| Hypertriglyceridaemia (4 studies) | OR 0.29 (0.13–0.65); RD -8.3%, NNT ≈ 12 | Very low (downgraded 3 levels) | Imprecision, risk of bias and indirectness each downgraded 1 level: only 4 studies and an interval width that depends on the interval estimation method; all studies open-label and two Gao cohorts may overlap; prolonged ICU sedation only. |
| Pruritus/paraesthesia (4 studies) | OR 32.56 (12.89–82.26); RD +28.5%, NNH ≈ 4 | Low (downgraded 2 levels) | Risk of bias downgraded 1 level: subjective outcome, mostly not double-blind; imprecision downgraded 1 level: only 4 studies and an imprecise magnitude. |
| Successful extubation (2 studies) | OR 1.08 (0.51–2.29); not significant (RD +1.7%) | Low (downgraded 2 levels) | Imprecision downgraded 2 levels: only 2 studies, few events, wide CI crossing the null; equivalence cannot be concluded. |

Note: certainty started at high; each downgrade lowers it by one level. RD, risk difference; NNT/NNH are reported side by side and do not by themselves establish net clinical benefit.

